# Nutritional deficiency recapitulates intestinal injury associated with environmental enteric dysfunction in patient-derived Organ Chips

**DOI:** 10.1101/2021.10.11.21264722

**Authors:** Amir Bein, Cicely W. Fadel, Ben Swenor, Wuji Cao, Rani K. Powers, Diogo M. Camacho, Arash Naziripour, Andrew Parsons, Nina LoGrande, Sanjay Sharma, Seongmin Kim, Sasan Jalili-Firoozinezhad, Jennifer Grant, David T. Breault, Junaid Iqbal, Asad Ali, Lee A Denson, Sean R. Moore, Rachelle Prantil-Baun, Girija Goyal, Donald E. Ingber

## Abstract

Environmental Enteric Dysfunction (EED) is a chronic inflammatory condition of the intestine characterized by villus blunting, compromised intestinal barrier function, and reduced nutrient absorption. Here, we show that key genotypic and phenotypic features of EED-associated intestinal injury can be reconstituted in a human intestine-on-a-chip (Intestine Chip) microfluidic culture device lined by organoid-derived intestinal epithelial cells from EED patients and cultured in niacinamide- and tryptophan-deficient (-N/-T) medium. Exposure of EED Intestine Chips to -N/-T deficiencies resulted in transcriptional changes similar to those seen in clinical EED patient samples including congruent changes in six of the top ten upregulated genes. Exposure of EED Intestine Chips or chips lined by healthy intestinal epithelium (healthy Intestine Chips) to -N/-T medium resulted in severe villus blunting and barrier dysfunction, as well as impairment of fatty acid uptake and amino acid transport. EED Intestine Chips exhibited reduced secretion of cytokines at baseline, but their production was significantly upregulated compared to healthy Intestine Chips when exposed to -N/-T deficiencies. The human Intestine Chip model of EED-associated intestinal injury may be useful for analyzing the molecular, genetic, and nutritional basis of this disease and can serve as a preclinical model for testing potential EED therapeutics.

## INTRODUCTION

Environmental Enteric Dysfunction (EED) is a pediatric disorder characterized by chronic intestinal inflammation that is associated with malnutrition, stunted growth, cognitive impairment, and attenuated response to oral vaccines ^1-5^. Previously described as “tropical enteropathy” or “environmental enteropathy”, EED has gained renewed interest in recent years due to its devastating effect on millions of children in low-and middle-income countries. The intestine of EED patients commonly exhibits villous atrophy, nutrient malabsorption, barrier dysfunction, and inflammation^6,7^. Currently, there is limited mechanistic understanding of the disease, which has hampered efforts to define biomarkers for diagnosis and therapeutics for effective prevention or treatment ^8^. For example, deficiencies of micronutrients, such as zinc ^9^ and vitamin A ^10^, may contribute to EED pathophysiology as they are associated with abnormal lactulose-mannitol (L:M) ratios, a measure of intestinal permeability. However, attempts at therapeutic intervention with nutritional replenishment have been disappointing likely due to ongoing intestinal dysfunction^8^. Similarly, other dietary interventions, such as administering omega-3 long-chain polyunsaturated fatty acids ^11^, optimizing amino acid profiles ^12^, supplementing with multiple micronutrients^13^ or improving food digestibility through fermentation, hydrolysis, or enzyme supplementation ^12^, also have been tested with limited success.

Beyond its role as a major manifestation of EED, malnutrition is also likely to actively contribute to disease pathophysiology. Murine diets low in the essential amino acid tryptophan lead to decreased antimicrobial peptide secretion and increased susceptibility to chemical-induced intestinal inflammation^14^. Low serum tryptophan levels are also linked to stunting in children suffering from EED ^15,16^. Tryptophan is both a building block for proteins and a precursor for niacin, melatonin, and neurotransmitters, including serotonin and tryptamine^17^. In animal studies, symptoms of tryptophan deficiency, including anorexia and impaired growth, may occur with intakes as little as 25% below the standard requirement, which translates to 2 to 2.5 mg/kg body weight for human infants 6-24 months old^17,18^. Recently, niacin (nicotinic acid) deficiency also has been implicated as a contributor to EED and other inflammatory intestinal conditions as administration of niacin was shown to ameliorate dextran sodium sulfate-induced colitis via prostaglandin D2-mediated D prostanoid receptor 1 activation^19^. In addition, niacin serves as a precursor for coenzymes, such as nicotinamide adenine dinucleotide (NAD) and nicotinamide adenine dinucleotide phosphate (NADP), which are essential for the normal function and survival of living cells. However, a mechanistic role for malnutrition in driving EED pathophysiology in humans remains to be demonstrated.

Studying a multifactorial disease, such as EED, raises substantial methodical and modeling challenges and, at present, there are only a few murine models and no existing human *in vitro* models that can be used to study this disease^20^. Thus, establishing an *in vitro* human EED model would help to elucidate disease pathophysiology and enable the development of new prevention and therapeutic measures. Here, we describe how human organ-on-a-chip (Organ Chip) microfluidic culture technology that faithfully recapitulates the structure and function of many human organs, including intestine^21-30^, can be leveraged to meet this challenge. Our studies using human Intestine Chips lined with organoid-derived primary intestinal epithelium isolated from either healthy or EED patients presented here reveal that both nutritional deficiencies and genetic or epigenetic changes in the intestinal epithelium contribute to the clinically observed EED phenotype. Moreover, by comparing healthy and EED patient-derived Intestine Chips, we were able to study phenotypic responses to nutritional deficiencies, such as villus blunting and barrier dysfunction, which are known to be common to multiple intestinal pathologies (for example, inflammatory bowel diseases, celiac), and which distinguish them from responses due to transcriptomic and cytokine signatures that are unique to EED.

## RESULTS

### Nutritionally deficient EED Chips recapitulate EED patient transcriptional signatures

We previously described a two-channel, microfluidic, human Intestine Chip lined with living human intestinal epithelium isolated from patient-derived organoids that undergoes villus differentiation, accumulates mucus, and exhibits many features of living human intestine when cultured on-chip under continuous flow with peristalsis-like mechanical deformations^27^ (**Fig. 1a**). Additionally, transcriptional analysis demonstrated that when lined by organoid-derived duodenal epithelium, this Intestine Chip more closely mimicked *in vivo* human duodenum than the organoids used to create the chips^27^. To define the contribution of the intestinal epithelium to the EED phenotype, we created Intestine Chips lined with intestinal epithelial cells from organoids derived from surgical biopsies of either healthy or EED patient duodenum (Healthy Chips and EED Chips, respectively). Compared to Healthy Chips, EED Chips showed differential expression of 287 genes (FDR < 0.05 and fold change ≥ 1.5; 86 upregulated, 201 downregulated) (**Fig. 1b**). EED chips showed upregulation of the protective mucin MUC5AC, neuregulin-4 (a known survival factor for colonic epithelium that protects against experimental intestinal injury^31,32^), and the intestinal stem cell marker SMOC2, whereas brush border peptidase MME, oxidative stress and inflammatory response controlling ectoenzyme VNN1, tight junction protein CLDN10, and secreted goblet cell protein CLCA1 were all down-regulated (**Supplementary Table 1**).

**Figure 1.**
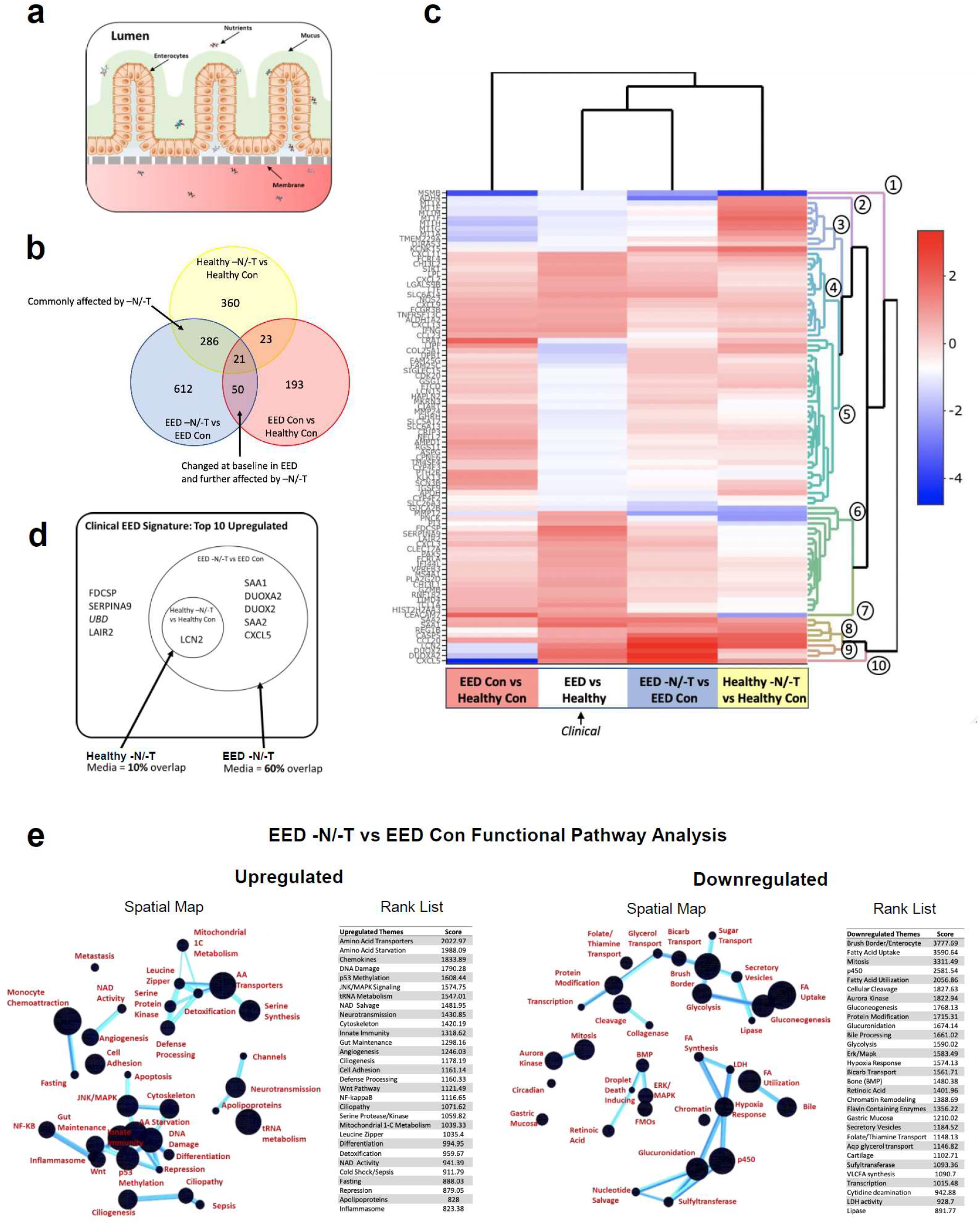
**a)** A schematic representation of Small Intestine chips. **b)** When compared to Healthy Chips, the EED transcriptome consists of 287 differentially expressed genes (red; FDR < 0.05 and fold change ≥ 1.5). Exposure of Healthy and EED Chips to -N/-T media resulted in an increased number of differentially expressed genes with 690 (yellow) and 969 genes (blue) respectively. Of these 307 genes were differentially expressed in both Healthy and EED Chips exposed to nutritional deficiency (yellow and blue). 71 genes were differentially expressed in EED Chips when compared to Healthy Chips in control media that are further affected by the addition of nutritionally deficient media (blue and red). n = 3 chips for each condition. **c)** A comparison of the 50 most up- and 50 most downregulated genes from the clinical EED signature with Healthy or EED Chip gene expression is depicted as a heatmap (red = upregulation, blue = downregulation) showing that EED -N/-T has the closest hierarchical relationship to the clinical EED signature. The comparison for EED -N/-T to Healthy Con is similar and shown in **Supplementary Fig. S9**. Ten dendrogram gene clusters were also defined and the corresponding roots enumerated. n = 3 chips for each condition. **d)** Of the top 9 upregulated genes in the clinical EED signature, 6 were also upregulated when EED chips were exposed to -N/-T media. N = 3 chips for each condition. **e)** Functional pathway analysis was performed using the contextual language processing program COmprehensive Multi-omics Platform for Biological InterpretatiOn (COMPBIO). The spatial map depicts themes related to differentially expressed genes as nodes with interconnections depicted as edges whose thickness relates to the degree of interconnectedness. The themes were also ranked according to a score representing fold enrichment over random clustering. n = 3 chips for each condition.

We compared this differential gene expression profile with a recently derived clinical EED signature, which was obtained by comparing profiles of intestinal tissue samples from EED patients who were refractory to nutritional intervention (Study of Environmental Enteropathy and Malnutrition, SEEM) versus samples from healthy control patients who were investigated for gastrointestinal symptoms but had normal endoscopic and histologic findings (Cincinnati Children’s Hospital Medical Center)^33^. When we compared gene profiles from EED Chips versus Healthy Chips cultured in control medium (i.e., with all nutrients present), the differentially expressed genes had some overlap with the clinical EED signature, including most notably a shared downregulation of metallothioneins (*MT1X, MT1A, MT1F, MT1H, MSMB, MT1M;* gene dendrogram cluster 3) (**Fig. 1c**).

We then carried out the same experiment but perfused both the Healthy and EED Intestine Chips with medium deficient in niacinamide and tryptophan (-N/-T), selected based on past work implicating their role in EED^15,16,19^. When we compared expression profiles from the Healthy Chips exposed to nutritional deficiency (Healthy -N/-T Chip) versus the Healthy Control Chips, we detected differential expression of 690 genes (FDR < 0.05 and fold change ≥ 1.5; 556 upregulated, 124 downregulated) (**Fig. 1b and Supplementary Fig.S1**) including upregulation of the amino acid starvation related transcription factor ATF4, its downstream solute carriers (SLC34A2, SLC7A5, SLC6A9) and the inflammation-associated gene LCN2 (**Supplementary Table 1**). There also appeared to be a trend towards upregulation of several antimicrobial and immune response genes as seen in the clinical EED signature, but these changes did not reach statistical significance. While there was greater overlap between the transcriptome of the Healthy -N/-T Chip with the clinical EED transcriptome than observed with the EED Control Chip compared to Healthy Control Chip, some genes were regulated in an opposing direction,including upregulation of metallothioneins (**Fig. 1c**, gene dendrogram cluster 3).

In contrast, we observed closer unsupervised hierarchical clustering with the clinical EED signature when the EED Chip was exposed to nutritional deficiency (EED -N/-T Chip vs EED Control Chip) (**Fig. 1c**). Culture of the EED Intestine Chips in -N/-T medium yielded differential expression of 969 genes (FDR < 0.05 and fold change ≥ 1.5; 522 upregulated, 447 downregulated) (**Fig. 1b**). This was manifested by upregulation of antimicrobial genes (*SAA1, SAA2, DUOXA2, DUOX2, CXCL5*; gene dendrogram clusters 8, 9 and 10) and downregulation of not only metallothioneins, but also metabolic and digestive genes (*SLC26A3, GUC2AB*; gene dendrogram clusters 5 and 6). This congruence was most striking amongst the top ten upregulated genes of the clinical EED signature, 6 of which (60%) were also upregulated when EED Chips were exposed to nutritionally deficient medium (**Fig. 1d**). Dendrogram cluster 5 was largely incongruent with the Intestine Chip transcriptional signature and included genes predominantly expressed in nerves, muscle and extracellular matrix.

To identify pathways affected by exposure of EED Intestine Chips to -N/-T nutritional deficiency, we used a contextual language-processing program to identify and rank functionally related clusters of genes ^34^. This analysis revealed several pathways that were significantly upregulated when EED Chips were exposed to -N/-T media, including chemokine pathway (score 1833.89, indicates fold enrichment over random association) and pathway associated with amino acid starvation (score 1988.09) (**Fig. 1e**). Within the amino acid starvation pathway, the ATF4 gene is upstream of several other pathways including tRNA metabolism (score 1547.01), DNA damage (score 1790.28), p53 methylation (score 1608.44), and amino acid transporters (score 2022.97). Many of these same pathways were also upregulated in nutritionally deficient Healthy -N/-T Chips. Conversely, nutritional deficiency led to down regulation of pathways related to fatty acid uptake (score 3590.64), brush border structural integrity (score 3777.69), mitosis (score 3311.49), cytochrome p450 (score 2581.54), and fatty acid utilization (score 2056.86) in the EED Chips. These results are consistent with the observation that the intestines of nutritionally deficient EED patients are characterized by having decreased brush border development and impaired cell growth^35-37^.

### Intestinal villus atrophy and barrier compromise

As the transcriptomic analysis revealed downregulation in pathways involved in cell growth and intestinal barrier formation, we carried out differential interference (DIC) and immunofluorescence microscopic analysis which indeed confirmed that both Healthy and EED Intestine Chips showed dramatically reduced growth of villus-like structures when cultured under nutrition deficient (-N/-T) conditions compared to Health and EED Control Chips perfused with complete medium (**Fig. 2a,b**). Quantification of the height of the epithelium revealed that removal of these nutrients resulted in significant villus blunting in both Healthy and EED Chips in response to -N/-T deficiency, as indicated by a 70% and 80% reduction in epithelial height, respectively, when compared to the same chips cultured in complete medium (**Fig. 2c**).

**Figure 2.**
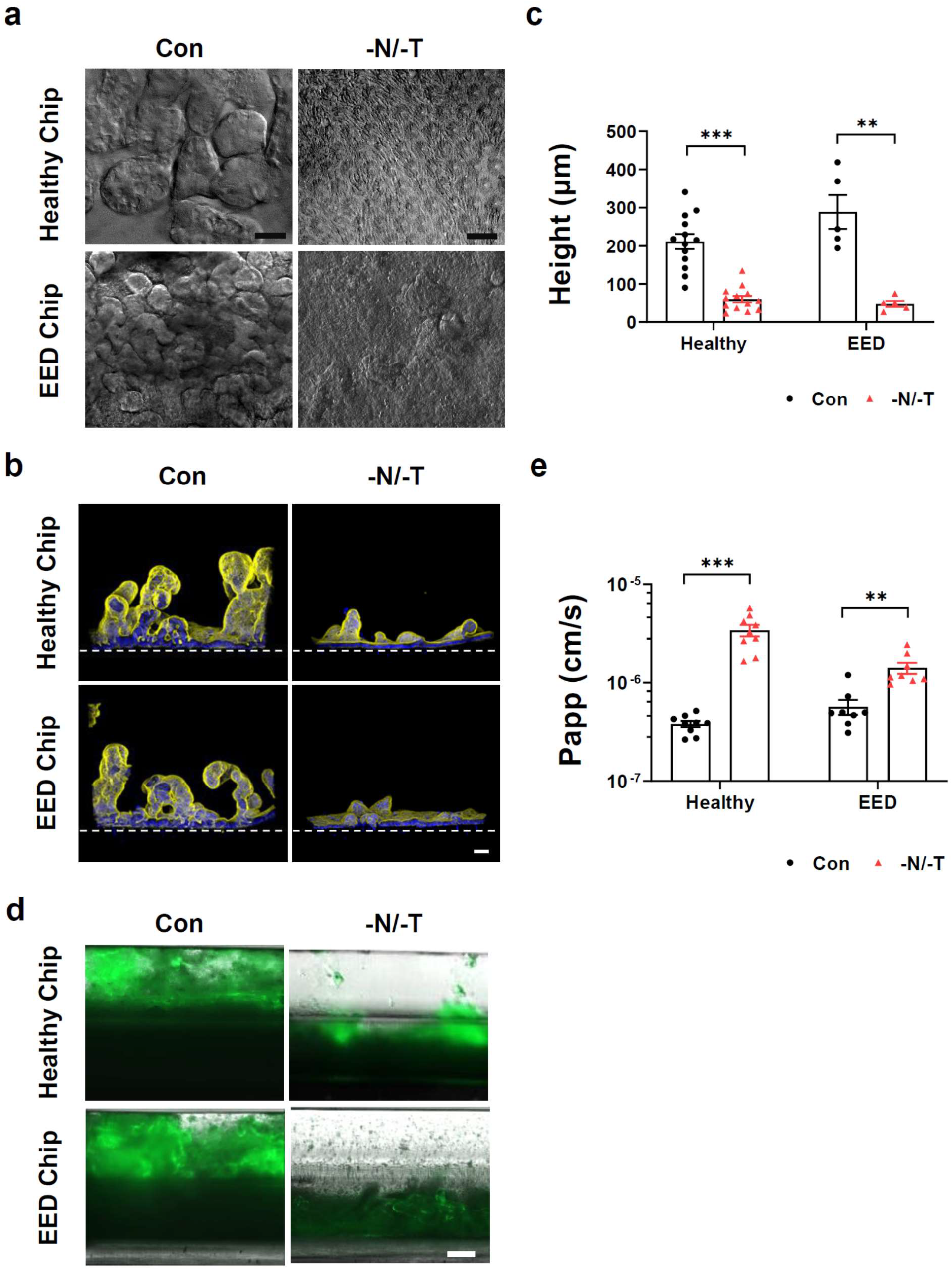
**a)** Differential Interference Contrast (DIC) imaging of Intestine chips, top-down view. Scale bar= 50 µm. Representative images. **b)** Immunofluorescence cross section micrographs showing villus like structures in the Intestine Chips. Yellow-phalloidin, Blue-Hoechst. Scale bar = 50 µm. Representetive image. **c)** Villus like structures height differences between Con and - N/-T, Healthy and EED Intestine Chips. Healthy, p<0.000001, EED, p= 0.000685. Each symbole on the graph represent an average of 4 to 5 measurment points (membrane to top of the villi) measured in two to three cross section images of Intestine Chip, at leaset 2 chips per condition were used. Two different Healthy donors and one EED donor were used. **d)** Immunofluorescence longitudinal section micrographs showing mucus layer in the Intestine Chips, stained with Alexa 488 conjugated lectin. Scale bar= 250 µm. Representative images. **e)** Apparent permeability (*Papp*), differences between Con and -N/-T, Healthy and EED Intestine Chips. Healthy, *p*= 0.000005; EED, p= 0.001382. n=9 for Healthy Intestine Chips and n=8 for EED Intestine Chips.

Another gene cluster identified as being preferentially sensitive to nutritional deficiency includes genes governing brush border structural integrity. These genes included myosin 1a (MYO1A), which links actin to the overlying apical membrane and whose absence results in irregularities of microvilli packing and length ^38^; protocadherin-24 (CDHR2) that forms links between adjacent microvilli and is the target of enterohemorrhagic *Escherichia coli*-mediated brush border damage ^39^; and mucin-like protocadherin (CDHR5) that forms heterophilic complexes with CDHR2 (**Supplementary Fig. S2**). Indeed, scanning electron microscopic (SEM) imaging revealed that culturing healthy intestinal epithelium in -N/-T medium on-chip resulted in severe loss of apical microvilli (as well as links between adjacent microvilli) relative to control enterocytes that had their entire surface covered with tightly packed microvilli (**Supplementary Fig. S3**). Consistent with these findings and the observed down regulation of the MUC5AC gene, live imaging and mucus staining with fluorescent lectin revealed that both the Healthy and EED Chips exhibited a much thinner mucus layer when exposed to nutrient deficient conditions (**Fig. 2d**).

Following these observations of structural changes due to exposure to nutritional deficiency, we next leveraged the advantage of using a two-channel microfluidic Intestine Chip (**Fig. 1a**) (e.g., as opposed to intestinal organoids cultured within a solid ECM gel) to assess the effect of exposure to -N/-T medium on functional differences in intestinal barrier function between Healthy and EED Chips. We compared apparent permeability (*P*_app_) values, which were measured by calculating the transfer of Cascade Blue fluorescent tracer (∼550 Da) from the epithelial lumen in the top channel to the underlying parallel channel below. Both Healthy and EED Control Chips exhibited a tight barrier under baseline conditions (within the 10^-7^ *P*_app_ range) and displayed small, but statistically significant reductions in barrier function when exposed to nutritional deficiency, as indicated by 8.9- and 2.5-fold increases in *P*_app_ for the Healthy and EED Chips, respectively (**Fig. 2e**).

### Reduced nutrient absorption

Our transcriptomic analysis also revealed that nutritional deficiency resulted in down regulation of multiple genes associated with the uptake and processing of important nutritional components, including fatty acids, certain amino acids, and carbohydrates (**Fig. 1d**). This is clinically relevant because reduced absorption of nutrients is another hallmark of EED, and it affects weight and linear growth as well as cognitive development in children^40-43^. For example, expression levels for fatty acid translocase (CD36), microsomal triglyceride transfer protein (MTTP), apolipoprotein B (ApoB), and apolipoprotein C-III (ApoC3) were all lower in nutritionally deficient epithelium (**Fig. 3a**). Similarly, when we used immunofluorescence microscopy to assess expression of ApoB protein, which is a marker of chylomicron and fat metabolism in the intestine ^44^, we found that exposure to -N/-T nutritional deficiency resulted in significant down regulation of ApoB in EED Intestine Chips (**Fig. 3b**). Furthermore, when we quantified cellular uptake of fatty acids using fluorescently-labelled dodecanoic acid, we found that exposure to nutritional deficiency reduced fatty acid uptake by 1.68- and 1.69-fold in Healthy -N/-T Chips and EED -N/-T Chips, respectively, compared to Healthy and EED Control Chips (**Fig. 3c**). These findings are consistent with clinical data that similarly show impaired fatty acid metabolism in children suffering from EED^16^ and suggest that nutritional deficiency alone is sufficient to reduce fatty acid uptake even in healthy intestine.

**Figure 3.**
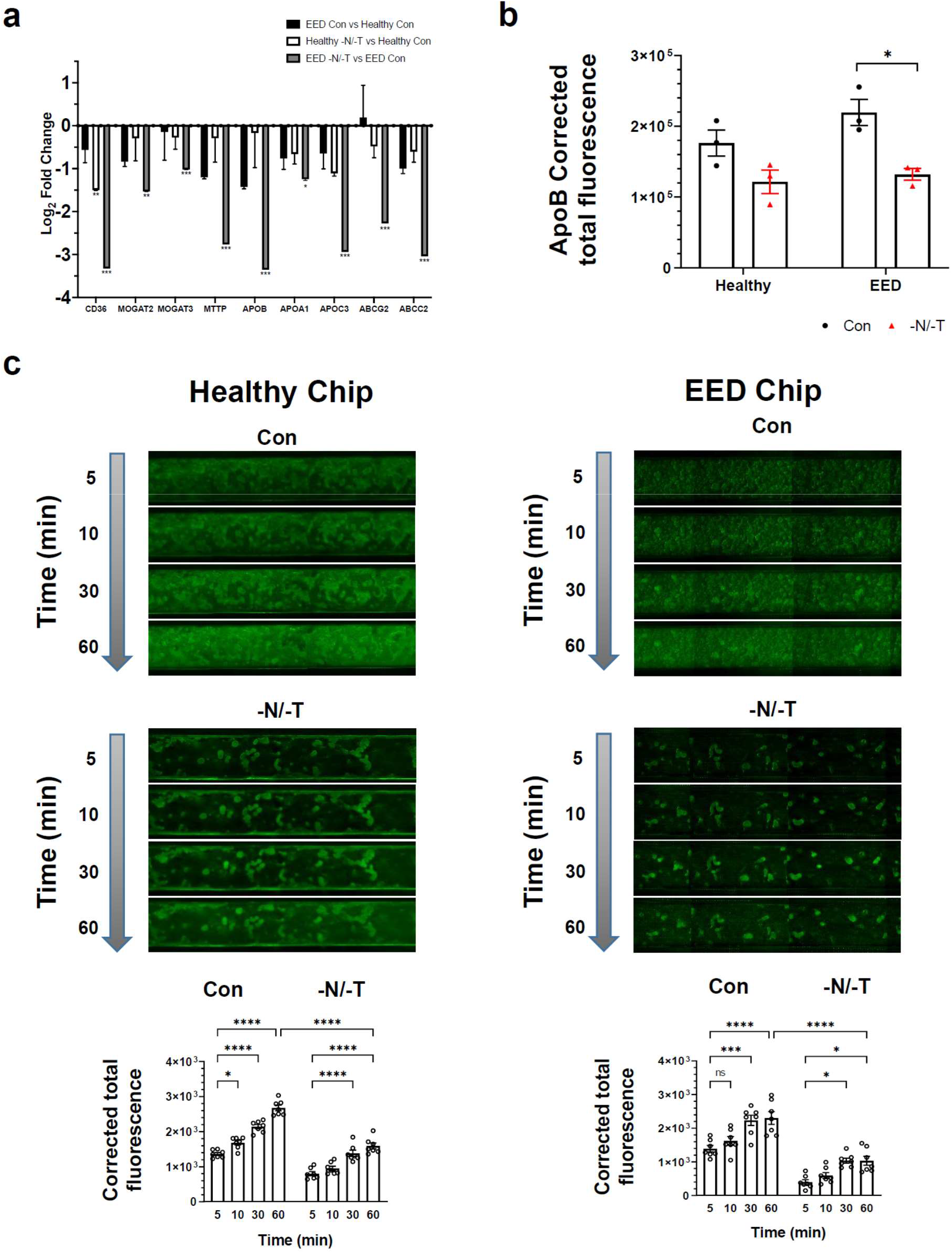
**a)** Transcriptional pathway analysis revealed a strong theme of downregulation for genes related to fatty acid uptake when EED Chips were exposed to -N/-T media. This included a 9.8-fold downregulation of the receptor CD36 (FDR < 0.001), a 10.2-fold downregulation of ApoB (FDR <0.001) and an 8.2-fold downregulation of ABCC2 (FDR <0.001). There was a similar trend towards downregulation when Healthy Chips were exposed to -N/-T media, but most gene changes did not reach statistical significance. n = 3 chips for each condition. **b)** Differences in ApoB corrected total fluorescence expression in Con and -N/-T Healthy and EED Intestine Chips. Healthy, not significant, EED, p= 0.012284. Each symbol on the graph represent an average of corrected total fluorescence expression from 3 different measurment areas of Intestine Chips cross sections, from at least 2 chips per condintion. **c)** Differences in fluorescently-labeled dodecanoic fatty acid uptake by the Intestine Chips at 5, 10, 30 and 60 minute time points. ^*^ *p*=0.0396, ^***^ *p*≤0.0008, ^****^ *p*<0.0001. For each time point, 7 images covering the entire area of a representative chip from each condition were used to calculate the corrected total fluorescence expression.

### Abnormal amino acid uptake and metabolism

Children suffering from EED exhibit impaired development, and protein availability from the diet is a key factor responsible for linear growth; thus, we next explored differences in uptake by Healthy and EED Intestine Chips. In the intestine, dietary protein is broken down into short peptides and free amino acids that are taken up by enterocytes, which serve as building blocks and energy sources for various organs and tissues^40^. Transcriptomic analysis revealed absorption and metabolizing factors that were down regulated in EED Control Chips vs Healthy Control Chips, including the solute carrier family 2 (facilitated glucose transporter) member 2 (SLC2A2) and solute carrier family 2 (facilitated glucose/fructose transporter) member 5 (SLC2A5). Other nutrient transporters (e.g., amino acid absorption and metabolizing factors) that were found to be significantly down regulated in EED Chips in response to -N/-T deficiency (EED -N/-T vs EED Con), include SLC36A1, which encodes the proton-coupled amino acid transporter 1, ANPEP membrane enzyme alanyl aminopeptidase that is responsible for peptide digestion at the brush border, retinol binding protein 2 (RBP2), cytochrome P450, family 27, subfamily A, and polypeptide 1 (CYP27A1) (**Fig. 4a and Supplementary Fig. S4**).

**Figure 4.**
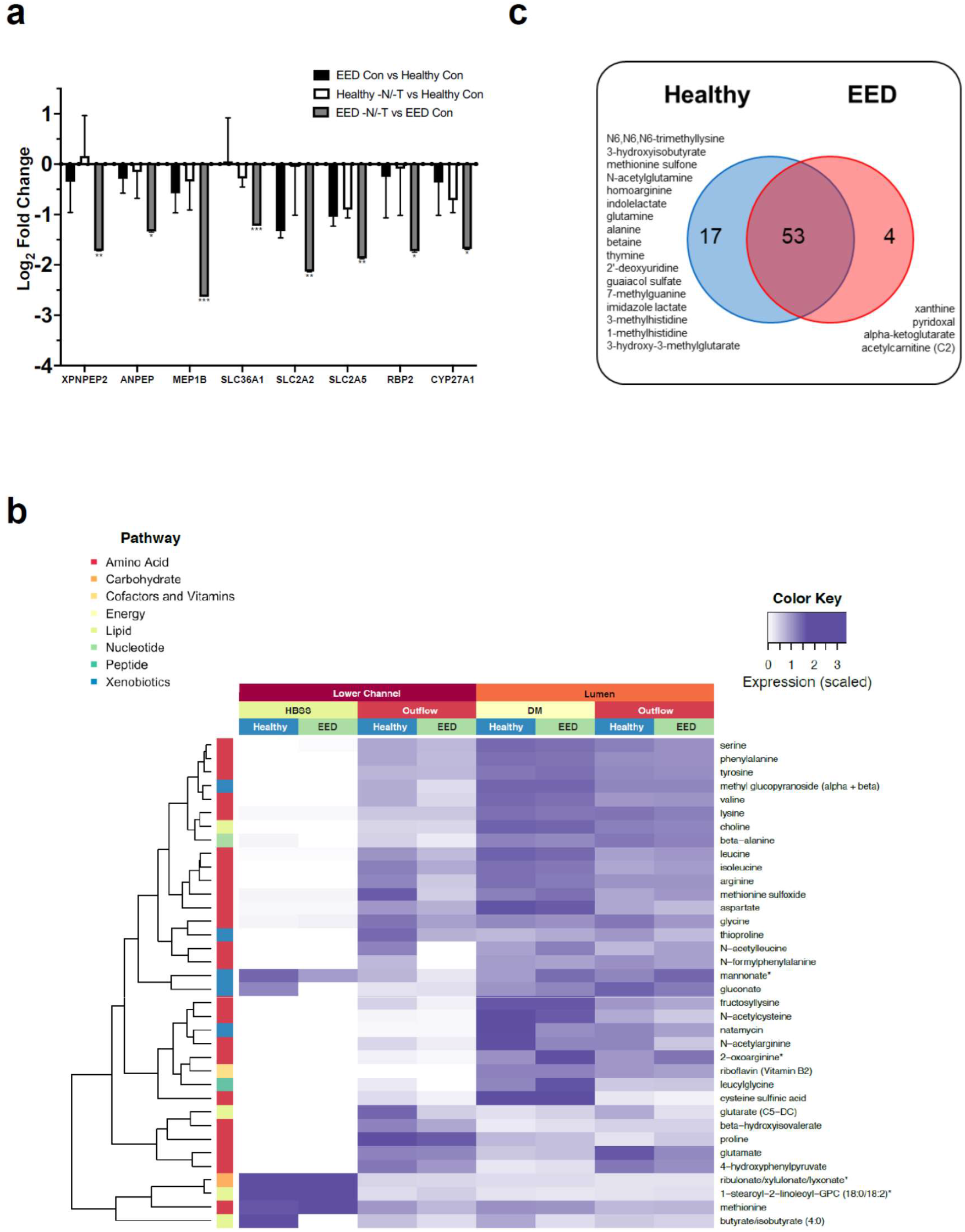
**a)** Amino acid processing and transport was among the strongly downregulated themes when EED Chips were exposed to -N/T media. This included a 6.2-fold downregulation of MEP1B (FDR < 0.001), a 4.4-fold downregulation of SLC2A2 and a 3.3-fold downregulation of XPNPEP2 (FDR < 0.01). There was occasional downregulation of these genes when Healthy Chips were exposed to -N/-T media, but none that achieved statistical significance. n = 3 chips for each condition. **b)** Heatmap showing 36 metabolites up-taken and transferred from the luminal medium to the lower channel of the Intestine Chips at higher abundance in the Healthy vs EED chips. n=3 for each condition. **c)** Venn diagram showing the number of common and unique metabolites secreted by the cells in Intestine Chips.

We then assessed differences in absorption of nutrients between the Healthy and EED Intestine Chips grown in control medium by performing untargeted metabolomic analysis by liquid chromatography tandem mass spectrometry (LC-MS/MS) to analyze epithelial uptake of nutrients and transfer of these molecules from the lumen of the intestinal epithelium in the top channel to the underlying basal channel (**Fig. 1a**). We detected 36 metabolites (out of >500 identified) that exhibited lower transport in EED Intestine Chips compared to Healthy Chips grown in control medium. These included mainly amino acids metabolites, but also metabolites related to nucleotide, cofactors, lipids, carbohydrates and xenobiotic pathways (**Fig. 4b;** an extanded list of metabolites analyzed can be found in **Supplementary Fig. S6**). Interestingly, nine of these metabolites were amino acids previously identified as being reduced in serum of Malawian stunted children^45^. These included essential amino acids (isoleucine, leucine, methionine, phenylalanine, lysine), conditionally essential amino acids (arginine, glycine), and non-essential amino acids (glutamate, serine).

Our LC-MS/MS analysis also revealed 74 metabolites that were secreted by the intestinal cells as they were absent or at extremely low levels (<5%) in the perfusion medium. 17 of these metabolites were unique to the Healthy Control Chips and included products of pathways related to metabolism of amino acids (N6,N6,N6-trimethyllysine, 3-hydroxyisobutyrate, methionine sulfone, N-acetylglutamine, homoarginine, indolelactate, glutamine, alanine, betaine, imidazole lactate, 3-methylhistidine, 1-methylhistidine), nucleotides (thymine, 2’-deoxyuridine, 7-methylguanine), xenobiotic metabolism (guaiacol sulfate), or lipids (3-hydroxy-3-methylglutarate). Interestingly, we also identified 4 metabolites unique to the EED Chips cultured in control medium, including products of purine nucleotide metabolism (xanthine), vitamin B6 metabolism (pyridoxal), citric acid cycle /energy metabolism (alpha-ketoglutarate), and fatty acid metabolism (acetylcarnitine (C2)) (**Fig. 4c**).

To assess if the observed differential uptake and metabolism of molecules in the Intestine Chips were transporter dependent, we quantified uptake of the dipeptide, glycyl-sarcosine (Gly-Sar) by the epithelial cells in the top channel, and its transfer to the lower channel using LC-MS/MS. This analysis revealed reduced uptake and transport of Gly-Sar in nutritionally deficient Healthy Intestine Chips when compared to the same chips cultured in control medium (**Supplementary Fig. S5**). In addition, these studies confirmed that these effects were due to transport through the PEPT1 transporter, and not due to passive inter- or intra-cellular diffusion, as this response could be completely prevented by adding Gly-Gly dipeptide (1 mM), which is a specific inhibitor of this transporter^46^ (**Supplementary Fig. S5**).

### Altered inflammatory mediators

Altered intestinal inflammation is a key component of EED and our transcriptional analysis revealed that genes encoding key inflammatory marker proteins, such as lipocalin 2 (LCN2) and regenerating islet-derived protein 3 alpha (REG3A) were upregulated when EED or Healthy Chips were exposed to nutritional deficiency (**Fig. 1c** and **Supplementary Fig. S7**). Indeed, when we quantified the expression of nine key intestinal cytokines using a multiplexed ELISA assay, we detected higher levels of several cytokines (IL-6, ICAM, VCAM, IL-33, MCP-1, MIP-1 alpha and IL-8) in the epithelial lumen of Healthy -N/-T Intestine Chips compared to the same chips cultured in control medium **(Fig. 5a)**. The antimicrobial peptide Reg3A was also downregulated by nutritional deficiency in these chips. Interestingly, EED Intestine Chips grown in complete medium displayed reduced levels of all of the secreted cytokines analyzed, while their levels were significantly upregulated when these chips were grown in nutritionally deficient medium **(Fig. 5a)**. Similar responses were observed when we analyzed cytokine levels in the lower parenchymal or vascular channel; however, there significantly higher levels of the inflammatory cytokines IL-8, IP-10, and MCP-1 were observed (**Fig. 5a**). Interestingly, we found that Intestine Chips that were not exposed to mechincal peristalsis-like deformation, secreted lower levels of IL-8 and MCP-1 (**Supplementary Fig. S8)**.

**Figure 5.**
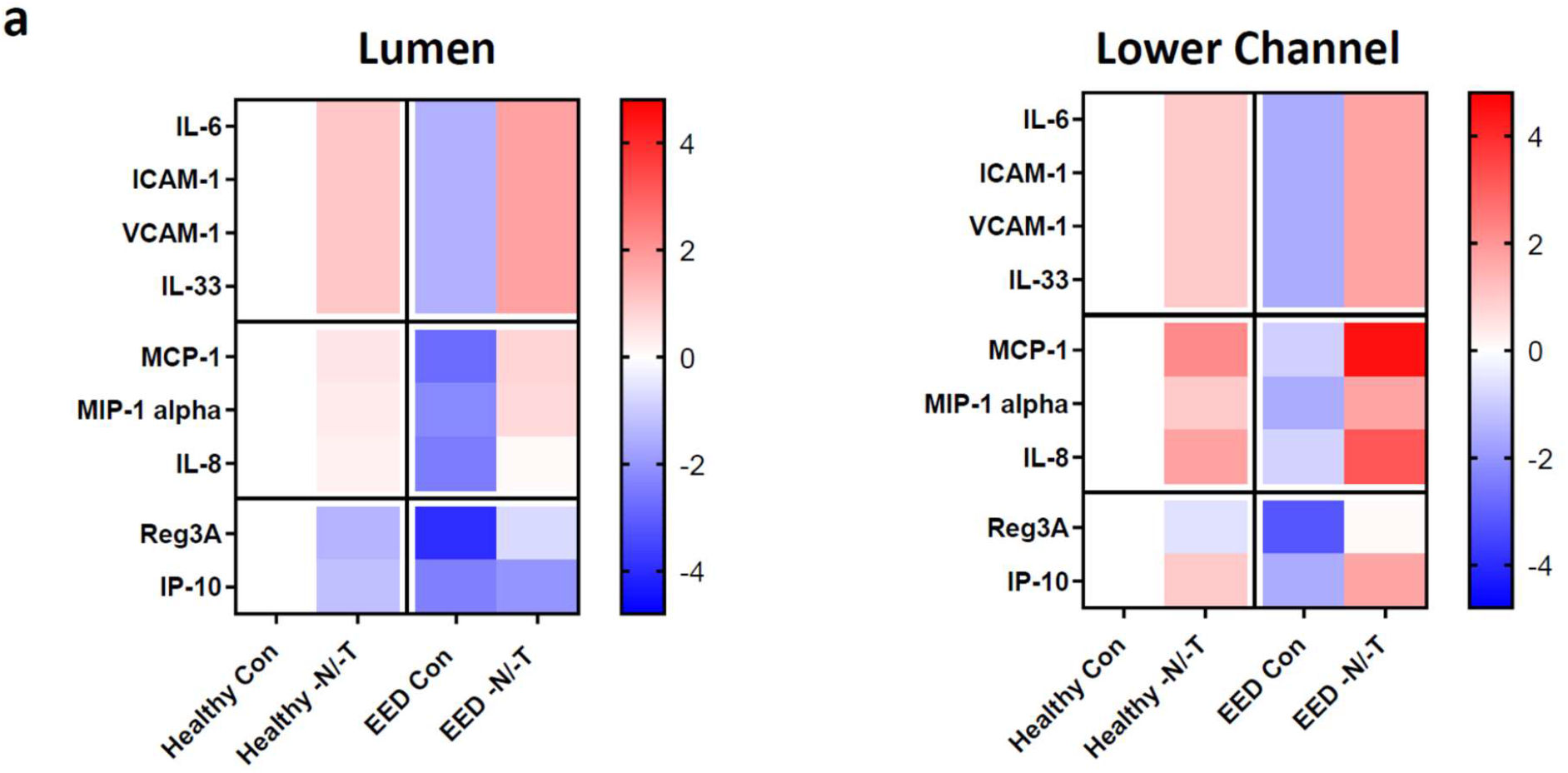
**a)** Heatmaps showing differential expression of 9 cytokines secreted into the lumen or lower channel of the Con or -N/-T, Healthy and EED Intestine Chips and quantified using a bead based multiplexed ELISA. Color coded scale represents Log_2_ fold change in expression. n=3 to 6 for each condition.

## DISCUSSION

Given the importance of studying the pathophysiology and the underlying mechanisms of EED, and the current lack of human *in vitro* models, this study leveraged the Human Intestine Chip technology to create an in vitro EED model using cells obtained from EED and healthy patient intestinal biopsies. The Intestine Chip offers a unique advantage over other advanced in vitro models, such as intestinal organoids, because direct access to the two parallel flow channels of the device enable quantitation of intestinal barrier function as well as transepithelial absorption and transport that are not possible in static 3D enteroid cultures. Using this approach, we explored the effect of nutritional deficiencies on the manifestation of the disease at both the phenotypic and functional levels. Importantly, our results showed that human Intestine Chips lined by intestinal epithelial cells isolated from organoids derived from EED patients mimic key features of the transcriptome signature of EED patients, including upregulation of antimicrobial genes and downregulation of metallothioneins and genes involved in digestion and metabolism, but only when exposed to nutritional deficiency, which we modeled by removing niacin and tryptophan from the medium. In contrast, nutritional deficiency induced similar intestinal villus atrophy, disruption of barrier function, and changes in amino acid and fatty acid absorption in chips lined by cells from both healthy and EED patients. Thus, we were able to attribute these various responses specifically to nutritional deficiency, to genetic or epigenetic changes in the intestinal epithelium, or to a combination of both, distinctions that have not been possible to make in clinical studies.

Malnutrition in EED can be regarded as both a cause and an outcome of the disease. In this study, we examined the effect of dual deficiency of an essential amino acid tryptophan and the vitamin niacin (in the form of niacinamide) because of their reported effect on intestinal development and function, and the suggested correlation between their deficiencies and EED development^45,47^. Remarkably, we observed a 6-fold increase in number of genes differentially expressed when we compared the EED Intestine Chips exposed to nutritional deficiency compared to Healthy Chips grown in complete medium. Moreover, many of the affected pathways were associated with nutritional uptake and cellular energy processing. These results imply that nutritional deficiency itself leads to derangements in nutritional processing that create a positive feedback loop, which further worsens the nutritional deficiency in EED patients.

Villus blunting and barrier dysfunction are hallmarks of EED, but also of other intestinal pathological conditions, such as inflammatory bowel diseases, celiac, diarrhea and small intestine bacterial overgrowth^48,49^. Indeed, we were able to show in this study that these phenotypes can manifest because of nutritional deficiencies regardless of whether the intestinal epithelium was derived from healthy or EED patients. This implies that the villus blunting and barrier dysfunction observed in EED, may be a response to environmental conditions, rather than an inherent genetic or developmental feature of this disease. This finding has high clinical relevance because the intestinal epithelium undergoes continuous shedding and renewal every 3-5 days; hence, the chronic negative effect of nutritional deficiencies might explain a fundamental aspect of the malabsorption and poor response to oral vaccination seen in EED patients. The ability to dissect and assess the factors leading to villus blunting in EED (as well as in other intestinal conditions) in vitro is a unique capability enabled by Intestine Chips that support formation of 3D villus-like structures as well as mucus production and quantification of transepithelial transport, absorption, and secretion.

Fat is a macronutrient responsible for 30 to 40% of total caloric intake in children and 20 to 35% in adults, and fatty acid composition has a direct effect on health and development, including inflammatory status and cognitive development.^50,51^ Our transcriptomic analysis was sensitive enough to detect several genes including ApoB that were downregulated in nutritionally deficient epithelium, and we confirmed that the expression of this molecule is decreased using immunofluorescence microscopy, and that fatty acid absorption is impaired in both EED -N/-T and Healthy -N/-T Chips (when compared to their respective controls), using a fatty acid uptake assay.

Given that dietary protein is an important macronutrient directly linked to linear growth and the results from our transcriptome-wide analysis revealed downregulation of amino acid transporters in EED Chips, we also conducted a untargeted metabolomic analysis that identified several amino acids which are significantly reduced in the EED Control Chips when compared to Healthy Control Chips. More interestingly, we identified metabolites that were secreted by the intestinal epithelium into the lower channel. Thus, the Intestine chip may be used as a nutritional and metabolic screening tool where uptake, utilization, and secretion of specific metabolites by and through the intestinal epithelium can be followed in high resolution and quantified over time. Moreover, future analysis of molecules released into the lower flow channel could lead to identification of biomarkers of disease severity and/or progression that might be detectable in blood.

While reduction of intestinal absorptive surface area due to villus blunting caused by nutritional deficiency will impair nutrient uptake by the intestine, exposure to nutritional deficiency also directly suppressed expression of multiple genes related to nutrient absorption specifically in chips lined by cells from EED patients. Thus, these results suggest that nutritional deficiency has a two-fold effect in these patients, which would likely manifest in a greater degree of intestinal dysfunction and a more severe EED phenotype. In our study, we also found that EED Intestine Chips exposed to nutritional deficiencies produced greater amounts of inflammatory cytokines compared to Healthy Chips grown under the same -N/-T conditions. This is a critical detail as inflamed intestine has higher caloric demands for basic maintenance and renewal, which could result in a negative caloric balance unable to support catch-up growth. Furthermore, chronic inflammation may negatively affect the efficacy of oral vaccines in the EED intestine. As such, it is possible that the lack of catch-up growth in EED children receiving nutritional intervention is due to current interventions only supplying nutrients aimed at replenishing the deficiency in tissues responsible for linear growth (bone, muscles). A more effective approach might be to first administer a diet composition that preferentially promotes intestinal recovery with new formation of villi and restores increased intestinal absorptive area before moving to supplementation required for catch-up growth.

The unique ability of the Intestine Chips to allow environmental factors (e.g., cell source, nutrient levels) individually or in combination in a controlle manner and to explore multiple clinically relevant outcomes, such as cell and tissue morphology, barrier function, nutrients metabolism and absorption, inflammatory status and transcriptome modifications, enabled us to distinguish between manifestations more common to multiple intestinal diseases versus responses that are more unique to EED. For example, we identified the cluster of differentiation 36 (CD36), gene, which has several functions relevant to EED as well as intestinal cancer and other intestinal diseases (e.g., fatty acid translocase, regulator of inflammation, oxidative stress, angiogenesis) to be significantly down regulated under nutritional deficiencies in both Healthy and EED Intestine Chips. Remarkably, our findings relating to EED are directly in line with recently published clinical data that explored unique signatures of EED affected children compared to healthy controls and children with celiac disease^33^. Thus, this *in vitro* EED model may be useful for gaining further insight into the pathophysiology of this disease as well as for development of potent therapeutics. The Intestine Chip also could provide a platform for personalized medicine and nutrition when cultured with clinical biopsies, enabling personalized (patient-specific) digestion, absorption, and allergic reactions to be assessed for different nutrients without putting the patient in danger.

## METHODS

### Organoid cultures and Intestine Chips

Organoids from Healthy donors or EED patients were generated from biopsy samples collected during exploratory gastroscopy following a procedure previously described^52^. A total of 3 Healthy and 2 EED donors were used to generate the data in this study (**Supplementary Table 2)**. For the Healthy Chips, de-identified endoscopic tissue biopsies were collected from grossly unaffected (macroscopically normal) areas of the duodenum in patients undergoing endoscopy for gastrointestinal complaints. Informed consent was obtained at Boston Children’s Hospital from the donors’ guardian. All methods were carried out in accordance with the Institutional Review Board of Boston Children’s Hospital (Protocol number IRB-P00000529) approval. For the EED Chips, de-identified endoscopic tissue biopsies were collected from affected areas of the duodenum in patients undergoing endoscopy following unsuccessful educational and nutritional intervention for wasting. Informed consent was obtained at the household level in a rural district of Matiari, Sind, Pakistan from the donors’ guardians. All methods were carried out in accordance with the AKU Ethical Review Committee’s approval (ERC number 3836-Ped-ERC-15). Organoids were kept in complete growth medium^27,52^, and passaged every 7 days in a 1:4 ratio. Before cell seeding, S-1 Chips (Emulate) were activated using ER1/2 (Emulate) and UV exposure for 20 minutes. Chips were then coated with 200 μg ml^-1^ collagen I (BD Corning) and 100 μg ml^-1^ Matrigel (BD Corning) in serum-free DMEM-F12 (Gibco) for 2 hours at 37 °C. After wash, organoids were broken into smaller fragments using enzymatic activity (TryplE, Gibco) and seeded in the luminal upper channel of the chips. They were then allowed to adhere for 24h before introduction of flow and mechanical deformation as described before^27^. For the -N/-T treatment, niacinamide and tryptophan were removed from the basal medium (DMEM-F12, Gibco) used to prepare the expansion culture medium (used for the luminal and lower channels) and no additional niacinamide was added^27^. After 16-18 days in culture with continuous flow (60 µl/h) and mechanical deformation (10%, 0.15 Hz), medium was changed to differentiation medium (serum and Wnt-3A free^27^, and -N/-T free for the respective group), in the luminal top channel and expansion culture medium in the lower channel for 4 additional days.

### Microarray sequencing and bioinformatics analysis

Initial microarray experiments were carried out using one healthy donor (n=3 biological replicates) and one EED donor (n=3 biological replicates) and were reflective of a recent clinical EED transcriptomic signature derived from a larger population (SEEM study, n=25 healthy donors, 52 EED donors). Subsequent validation studies were carried out using 1-3 healthy donors (n=3-9 biological replicates) and 1-2 EED donors (n=3-8 biological replicates) per experiment. RNA samples were processed using the GeneChip WT PLUS Reagent Kit and hybridized to Affymetrix Human Clariom D arrays. Samples were pre-processed with SCAN (SCAN.UPC package) and resulting expression values were quantile-normalized within the experiment, and differential expression analysis was performed with limma^53^ R package for each comparison pair; gene expression values were averaged for each condition. Template matching was used to extract genes that are differentially expressed between these conditions. Differential gene expression heatmap analysis was preformed using Euclidean distance and McQuitty’s linkage within the R package heatmaply ^34^. To normalize values across experiments, columns were scaled to generate a Z-score. Pathway analysis was performed using the natural language processing algorithm COmprehensive Multi-omics Platform for Biological InterpretatiOn (COMPBIO) to generate a holistic, contextual map of the core biological themes associated with gene expression changes. Enriched concepts associated with differentially expressed genes were compiled from PubMed abstracts using contextual language processing. Themes were scored using a complex function that incorporates an empirical p value and a standard score similar to a Z-score to give a final value representing fold enrichment over a random clustering as follows: 3-9 = weak relationship, 10-99 = modest relationship, 100-999 = strong relationship, 1000+ = very strong relationship. A theme map was generated where themes are represented as nodes and interconnections between themes are represented as edges with the thickness of an edge relating to the degree of interconnection.

### Immunofluorescence microscopy

Immunofluorescence microscopic imaging was carried out using the following steps: the apical and basal channels of the chips were gently washed with PBS and fixed with 4% paraformaldehyde (Electron Microscopy Sciences, Cat#: 157-4) in PBS for 30 min, then washed twice with PBS and kept at 4°C. The fixed samples were sectioned to 150-250 µm sections using a vibratome (Leica), and then permeabilized and blocked with 0.1% Triton X-100 solution and 10% Donkey serum in PBS, for 30 min at room temperature. Then primary antibody (Apo-B, Abcam, Cat#:ab20737) was added (1:100 in 1.5% BSA/PBS solution) and incubated overnight at 4 °C, followed by multiple PBS washes. Cells were then incubated with secondary fluorescent antibody (Invitrogen Cat#:SA5-10038) and Phalloidin (Invitrogen Cat#: A12380) at room temperature for 60 min and washed with PBS; nuclei were co-stained with hoechst 33342 (Sigma, Cat#: 14533). Microscopy was performed with a laser scanning confocal microscope (Leica SP5 X MP DMI-6000 or Zeiss TIRF/LSM 710).

### Paracellular permeability measurements

To assess paracellular permeability, 50 μg ml^-1^ of Cascade blue (5.9 kDa; Thermo Fisher, C687) was introduced to the luminal channel (at 60 ml h^-1^). After flowing over-night to saturate the microfluidic channels, outflows were discarded and collection for measurements started for ∼24 hours. The fluorescence intensities (390nm/420nm) of the top and bottom channel effluents were measured using a multimode plate reader (BioTek NEO). The apical to-basolateral flux of the paracellular marker was calculated using the following equation: Papp=(dQ/dt)/AdC. dQ/dt (g s^-1^) is molecular flux, A (cm^2^) is the total area of diffusion and dC (mg ml^-1^) is the average gradient.

### Mucus assessment

Mucus was visualized using a WGA-Alexa Fluor 488 conjugate (Thermo Fisher Scientific, Cat#: W11261) for live cell imaging, as described previously^25^ with some modifications. Briefly WGA solution (25 μg ml^-1^ in HBSS) was flowed through the epithelium channel for 30 minutes and then washed with continuous flow of HBSS for 30 min. Intestine chips were then cut sideways parallel to the length of the channel and imaged with an epifluorescence microscope (Zeiss Axio Observer Z1) with 5x objective.

### Fatty Acid Uptake

Chips were starved for 1 hour by replacing the luminal and lower channel media with HBSS. Then, fluorescently-labelled dodecanoic acid combined with a quencher (to eliminate any unspecific signal), were added according to the manufacturer instructions (BioVision, Cat#: K408) to the luminal upper channel of the intestine chips. The entire length of the channels was then imaged with an epifluorescence microscope (Ex/Em = 488/523 nm, Zeiss Axio Observer Z1) with 5x objective, at 5, 10, 30 and 60 minutes.

### Metabolomics

Medium in the lower channel of the chips was changed to HBSS and flowed for 30 minutes to clear any residues. Then collection of outflows was conducted over 5 hours. Samples were frozen immediately after collection (-80 °C), and submitted for LC-MS/MS analysis. The Metabolon global metabolomics platform was used to measure biochemicals in cell and media samples. Samples were prepared using the automated MicroLab STAR^®^ system from Hamilton Company. Several recovery standards were added prior to the first step in the extraction process for QC purposes. To remove protein, dissociate small molecules bound to protein or trapped in the precipitated protein matrix, and to recover chemically diverse metabolites, proteins were precipitated with methanol under vigorous shaking for 2 min (Glen Mills GenoGrinder 2000) followed by centrifugation. The resulting extract was divided into five fractions: two for analysis by two separate reverse phase (RP)/UPLC-MS/MS methods with positive ion mode electrospray ionization (ESI), one for analysis by RP/UPLC-MS/MS with negative ion mode ESI, one for analysis by HILIC/UPLC-MS/MS with negative ion mode ESI, and one sample was reserved for backup. Samples were placed briefly on a TurboVap^®^ (Zymark) to remove the organic solvent. Raw data was extracted, peak-identified and QC processed using Metabolon’s hardware and software.

Beginning with the OrigScale values from Metabolon, which are normalized in terms of raw area counts, we calculated total ion count in outflow to visualize sample-wise variance.

Next, we normalized each sample by its total ion count and rescaled each metabolite values by dividing each metabolite by its root mean square. We visualized metabolite abundance using a heatmap. Statistical analysis and heatmap generation was performed using R (R Foundation for Statistical Computing, Vienna, Austria). We performed differential expression analysis by using the limma^53^ R package (v3.32.10) was used to fit a linear model to the data. Log_2_ fold change, *p*-value and adjusted *p*-value were calculated for each comparison using an unmoderated Student’s t-test and the FDR method for multiple testing correction^54^. Adjusted *p*-values are shown in the volcano plots. To assess transporter mediated uptake and transfer from the luminal upper channel to the lower channel, glycyl-sarcosine (Gly-Sar, 1 mM, Sigma) alone or in combination with the specific PEPT1 transporter inhibitor Gly-Gly dipeptide (1 mM, Sigma), were added to luminal medium and their abundance in the lower channel outflow was assessed.

### Statistical analysis

Each Intestine Chip was used as a biological repeat for one terminal assay. Either a Student’s t-test or 2-way ANOVA was performed to determine statistical significance, as indicated in the figure legends (error bars indicate standard error of the mean (SEM); *p*-values < 0.05 were considered to be significant).

## Data Availability

All data produced in the present study are available upon reasonable request to the authors

## ACKNOWLEDGMENTS

This research was sponsored by funding from the Bill and Melinda Gates Foundation (independent support to D.E.I), NIH award DK119488 (to D.T.B), and the Wyss Institute for Biologically Inspired Engineering (to D.E.I). This work was conducted with the support of a KL2 award (an appointed KL2 award) from Harvard Catalyst | The Harvard Clinical and Translational Science Center (National Center for Advancing Translational Sciences, National Institutes of Health Award KL2 TR002542). The content is solely the responsibility of the authors and does not necessarily represent the official views of Harvard Catalyst, Harvard University and its affiliated academic healthcare centers, or the National Institutes of Health.

## AUTHOR CONTRIBUTIONS

A.B, C.W.F, G.G and D.E.I. designed the research. A.B, C.W.F, B.S, W.C, A.N, N.L, S.S, S.K, and S.J.F performed experiments. A.B, C.W.F, W.C, R.K.P, D.M.C, A.P, J.G, R.P.B, G.G and D.E.I. analysed and interpreted the data. D.T.B. established and prepared human Healthy organoids.

J.I and A.A established and prepared human EED organoids. L.A.D and S.R.M provided the clinical data. A.B, C.W.F and D.E.I. wrote the Article with input from G.G. All authors reviewed, discussed and edited the manuscript.

## DATA AVAILABILITY STATEMENT

Organ chip microarray data that support the findings of this study have been deposited in Gene Expression Omnibus (GEO) with the accession codes to be determined prior to publication. Clinical mRNASeq data referenced as a comparison are deposited in Gene Expression Omnibus under accession number <u>GSE159495</u>. Unique biological materials used in this study are available from the corresponding author on reasonable request.

---

**Supplementary Figure 1.**
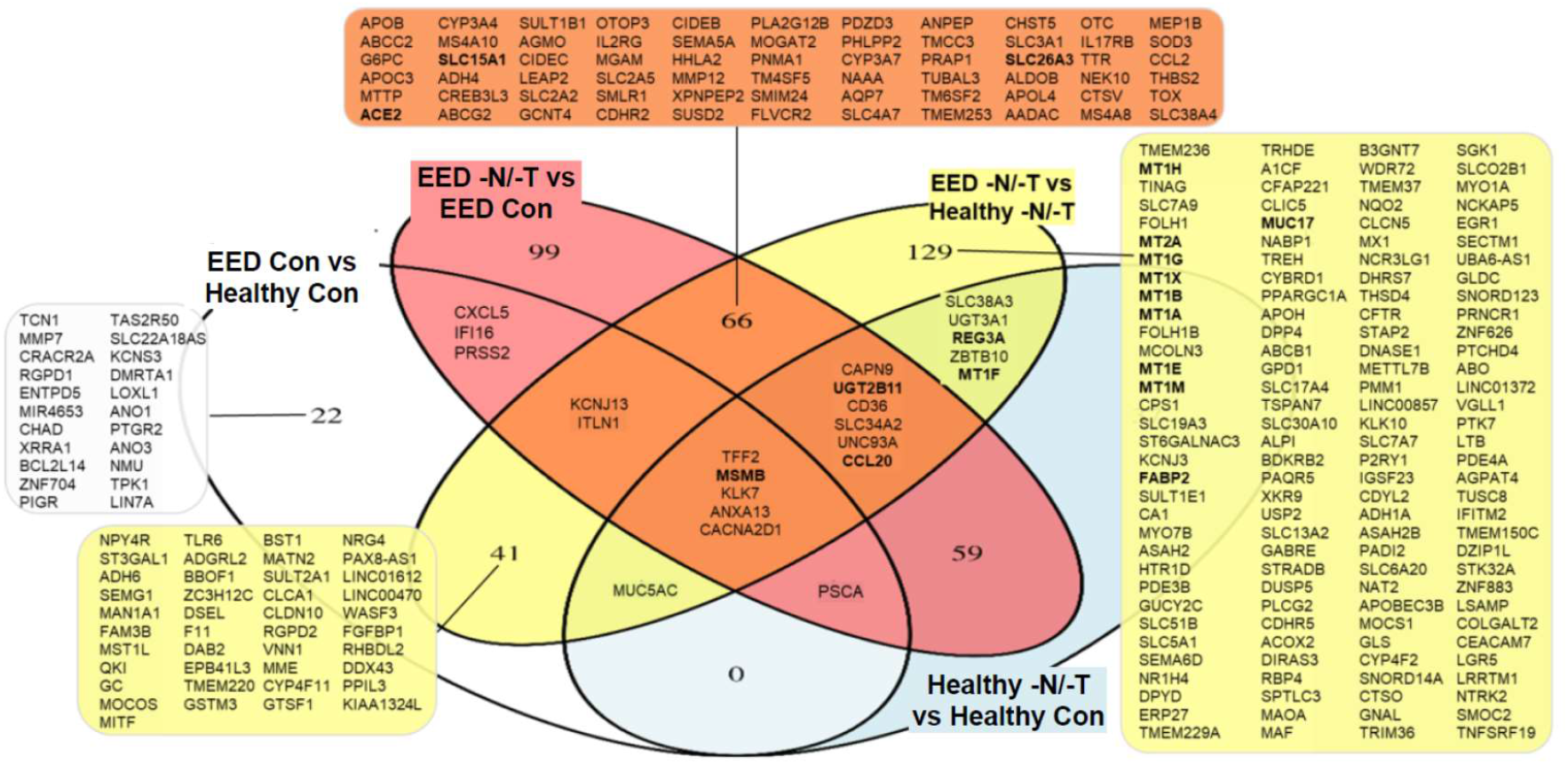
A Venn diagram representation showing overlap of differentially regulated genes (*p*-value < 0.01, fold change > 2) for EED and Healthy Chips exposed to complete and nutritionally deficient media. Affected genes are listed and genes that were also differentially expressed in the clinical EED gene signature are in bold. n = 3 chips for each condition.

**Supplementary Figure 2.**
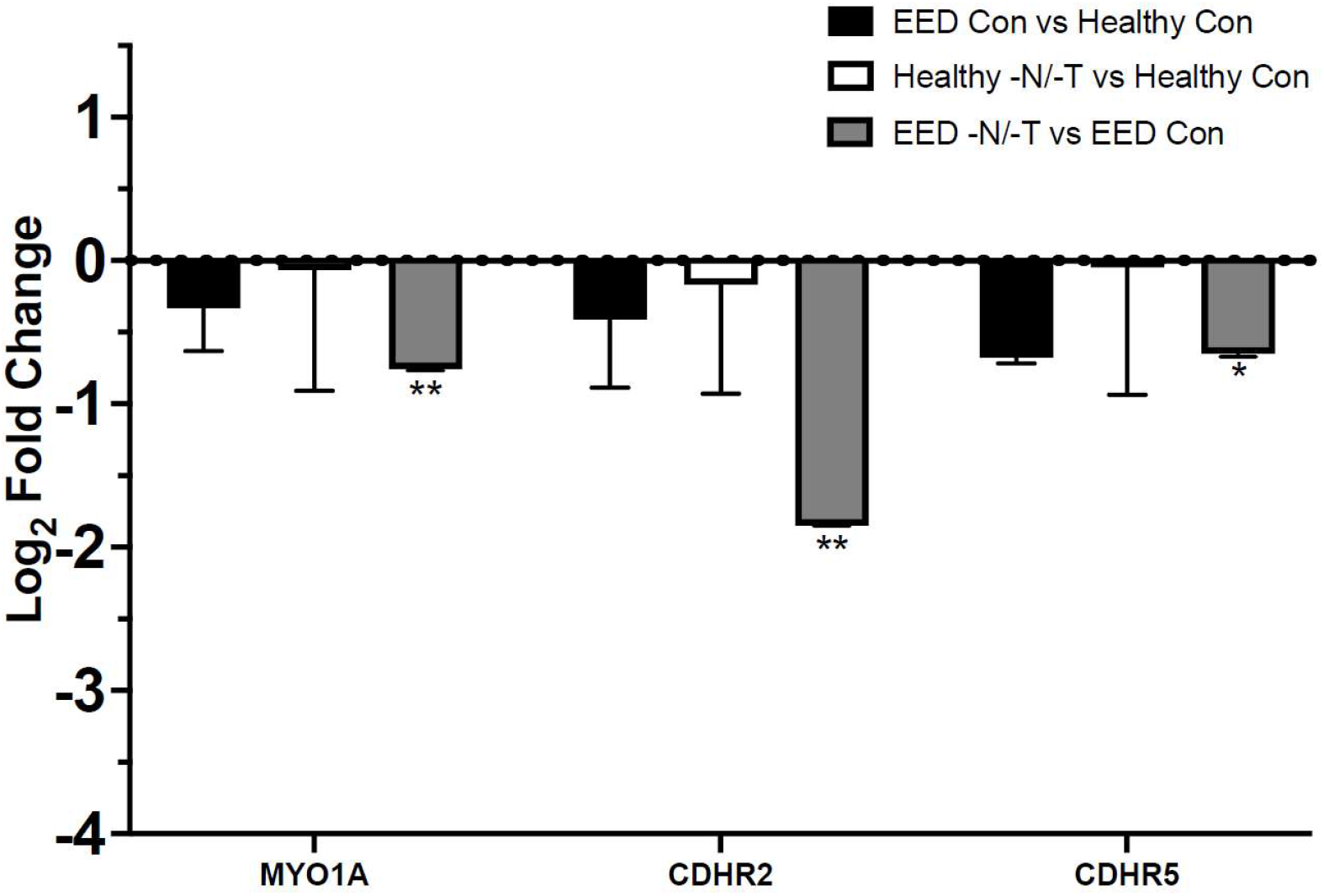
Transcriptional pathway analysis revealed a strong theme of downregulation for genes related to brush border localization and structural integrity when EED Chips were exposed to -N/-T media. This included a 3.6-fold downregulation of CDHR2 (FDR < 0.01) and more modest downregulation of MYO1A (1.7-fold, FDR <0.01) and CDHR5 (1.6-fold, FDR < 0.05). n = 3 chips for each condition.

**Supplementary Figure 3.**
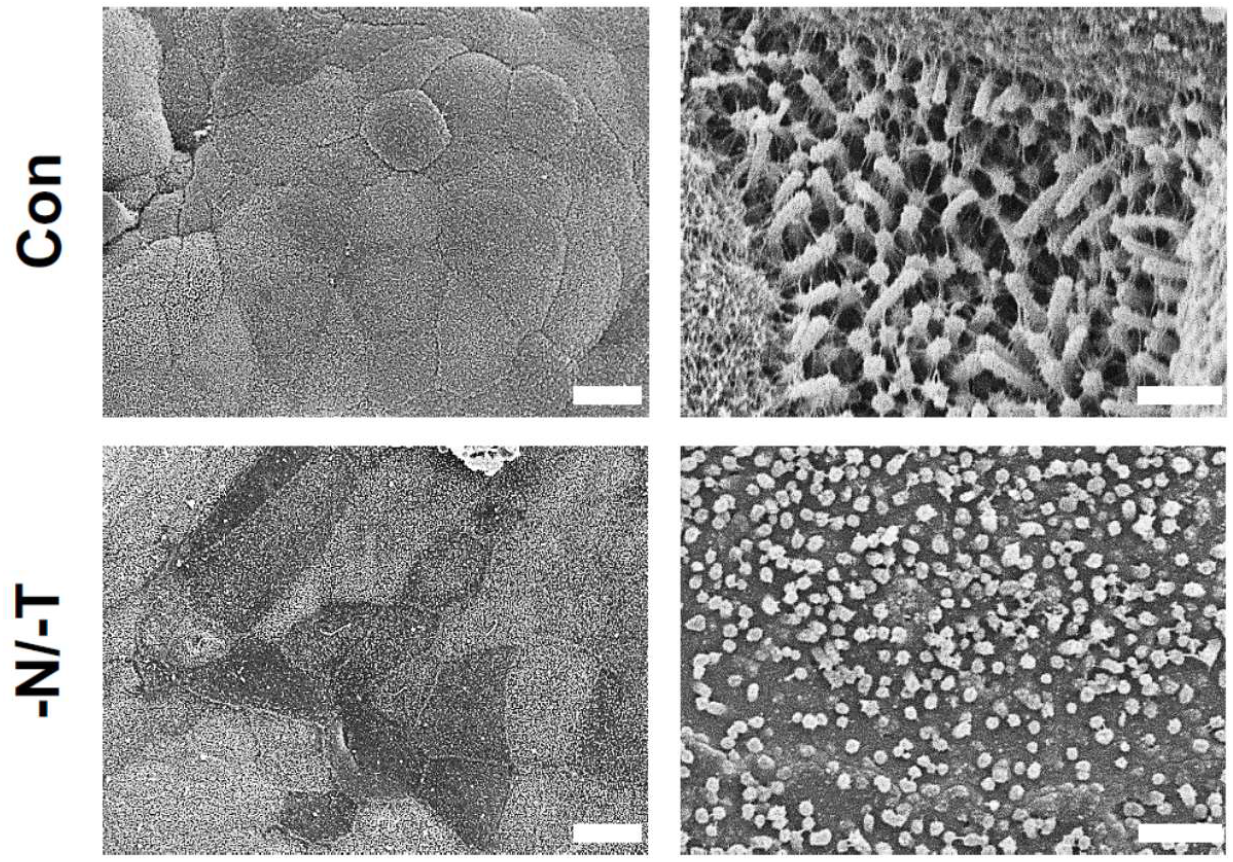
SEM views of the apical surface in the Intestine Chip comparing the morphology of Control and -N/-T treated Healthy chips. Scale bar, 20 μm (left) and 2 μm (right). Representative images.

**Supplementary Figure 4.**
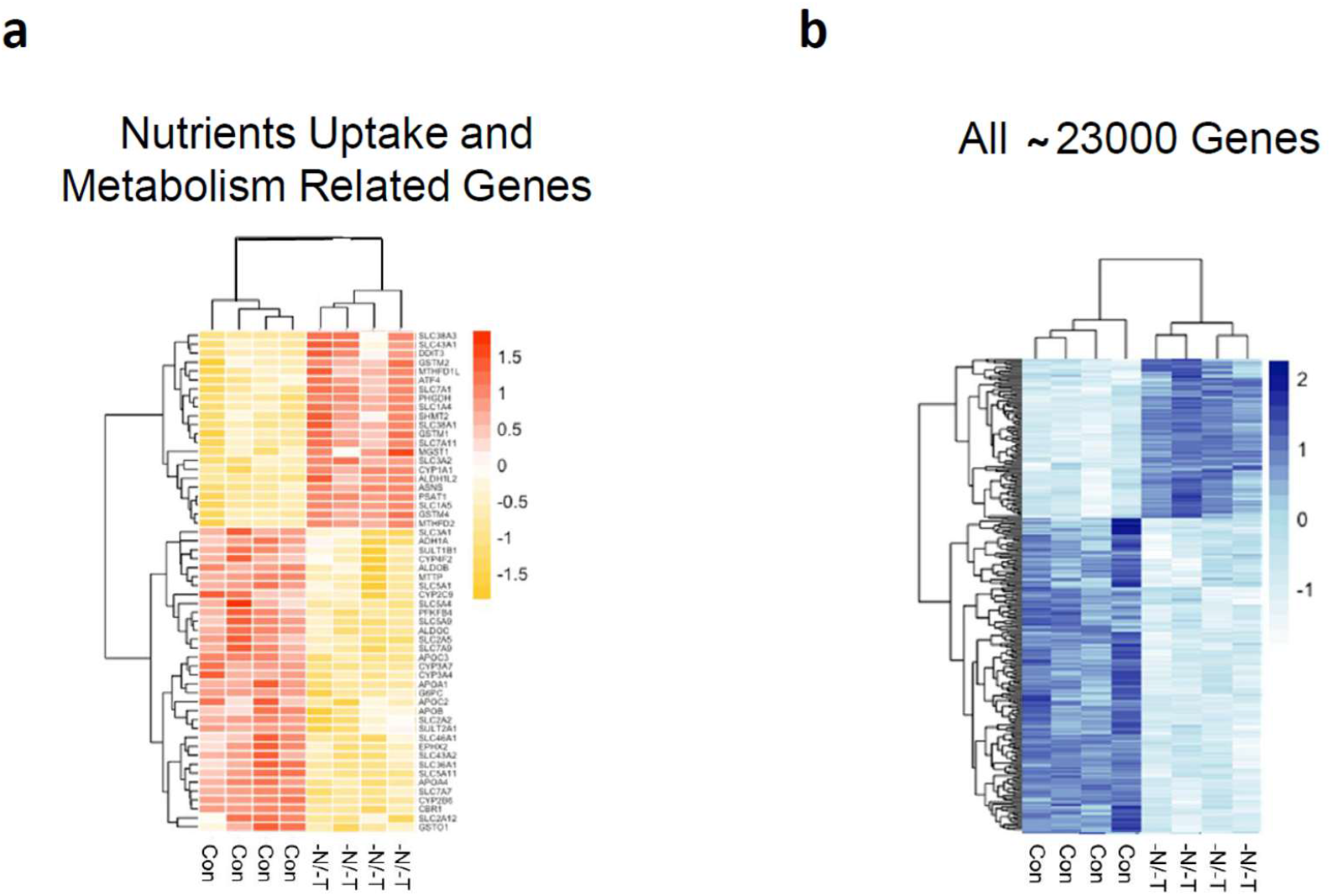
a) Heatmap showing up and down regulated nutrients uptake and metabolism related genes in Control and -N/-T Healthy Chips. b) Heatmap showing all ∼ 23000 up and down regulated genes in control and -N/-T healthy chips. n=4 for each condition.

**Supplementary Figure 5.**
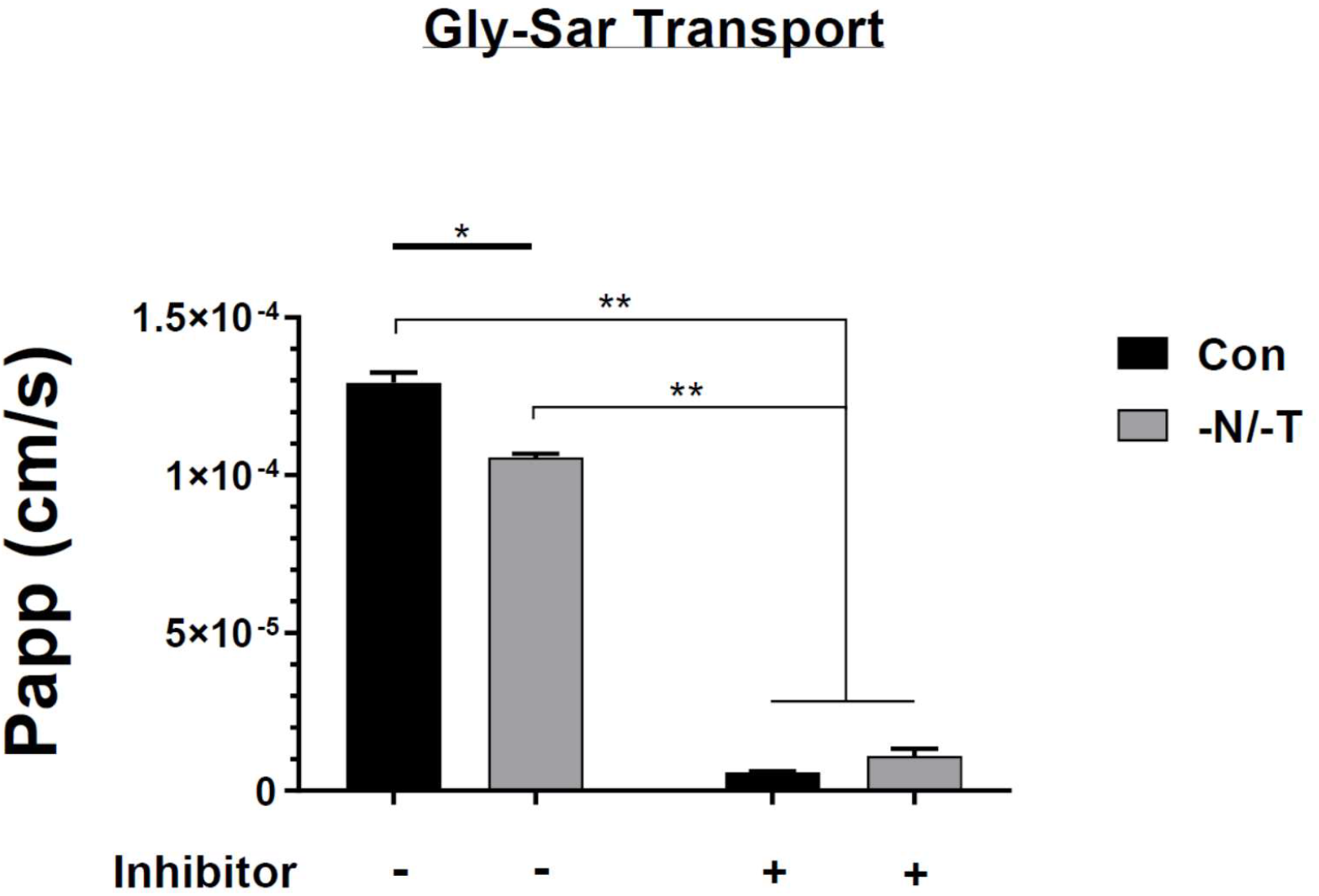
Calculated apparent permeability (Papp) of Gly-Sar dipeptide transfer from the lumen to the lower channel showing differences between Con and -N/-T, in Healthy Intestine Chips, in the presence or absence of Gly-Gly inhibitor. Con vs -N/-T, p= 0.0022, with vs without inhibitor *p*= 0.000003. n=3 for each condition.

**Supplementary Figure 6.**
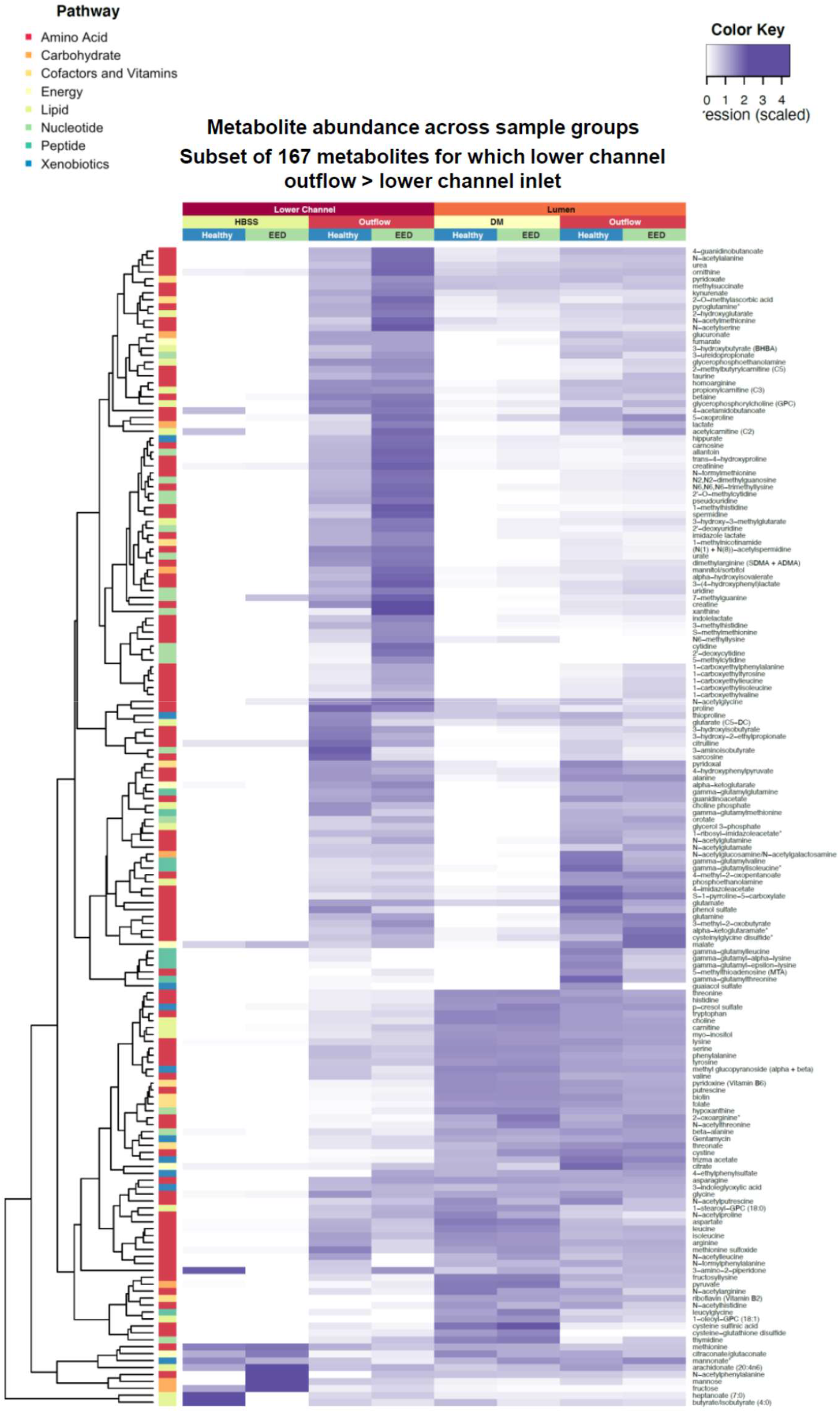
Heatmap showing 167 metabolites found to be at higher abundance in the lower channel outflow vs lower channel inlet. n=3 for each condition.

**Supplementary Figure 7.**
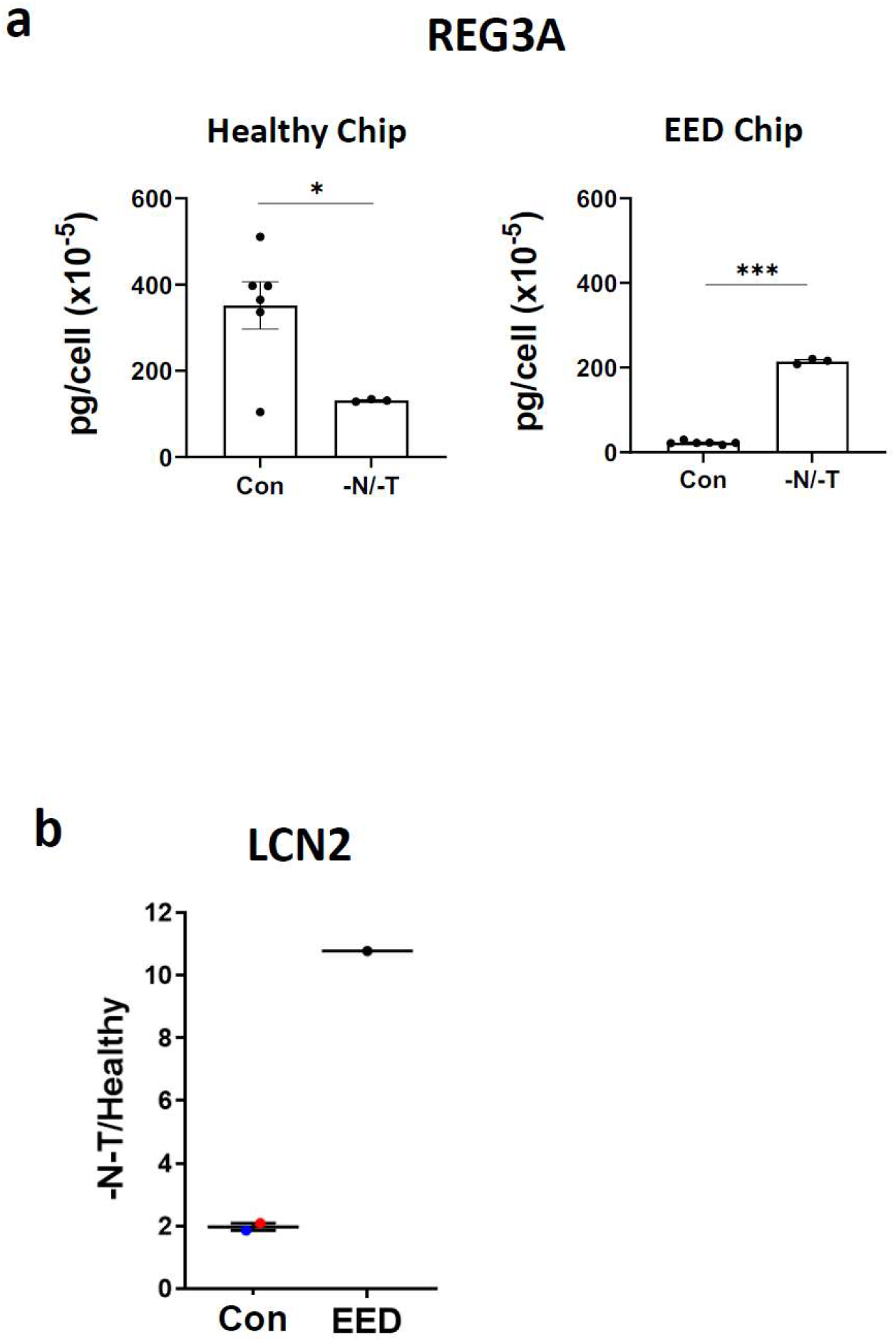
a) Secreted REG3A levels measured in Intestine Chips luminal outflows. Healthy Chip, *p*=0.029, EED Chip, *p*<0.000001. n=3 to 6 for each condition. **b)** Lipocalin 2 (LCN2) relative expression measured by ELISA.

**Supplementary Figure 8.**
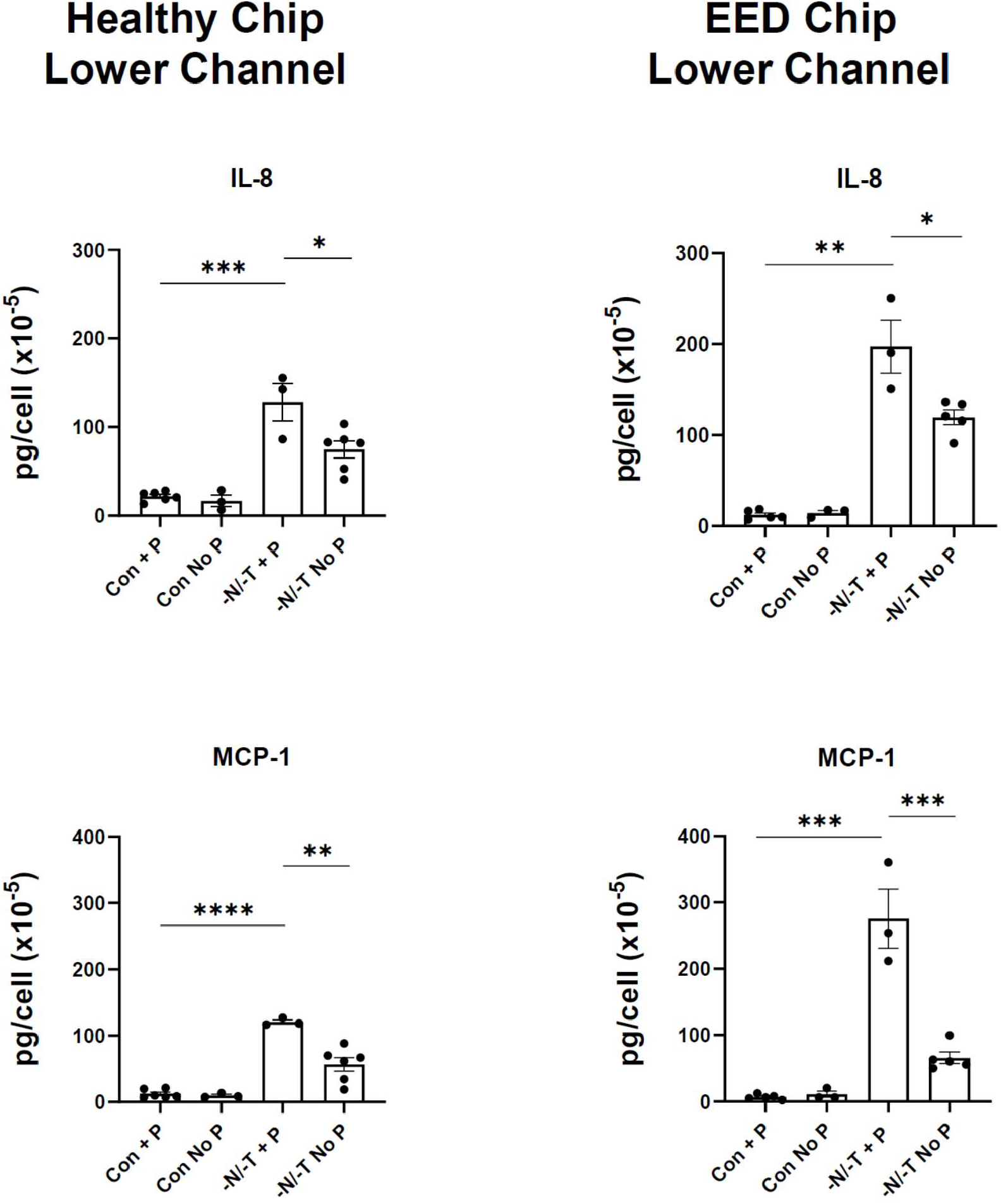
Secreted IL-8 levels measured in the Intestine Chips lower channel outflows, with and without peristalsis (P). Healthy Chip Con + P vs -N/-T + P, *p*= 0.000148; -N/-T + P vs -N/-T no P, *p*= 0.029388. EED Chip Con + P vs -N/-T + P, *p*= 0.000129; -N/-T + P vs -N/-T no P, *p*= 0.016852. Secreted MCP-1 levels measured in the Intestine Chips lower channel outflows, with and without peristalsis. Healthy Chip Con + P vs -N/-T + P, *p* <0.000001; -N/-T + P vs -N/-T no P, p= 0.004174. EED Chip Con + P vs -N/-T + P, p= 0.000165; -N/-T + P vs -N/-T no P, p= 0.000884. n=3 to 6 for each condition.

**Supplementary Figure 9.**
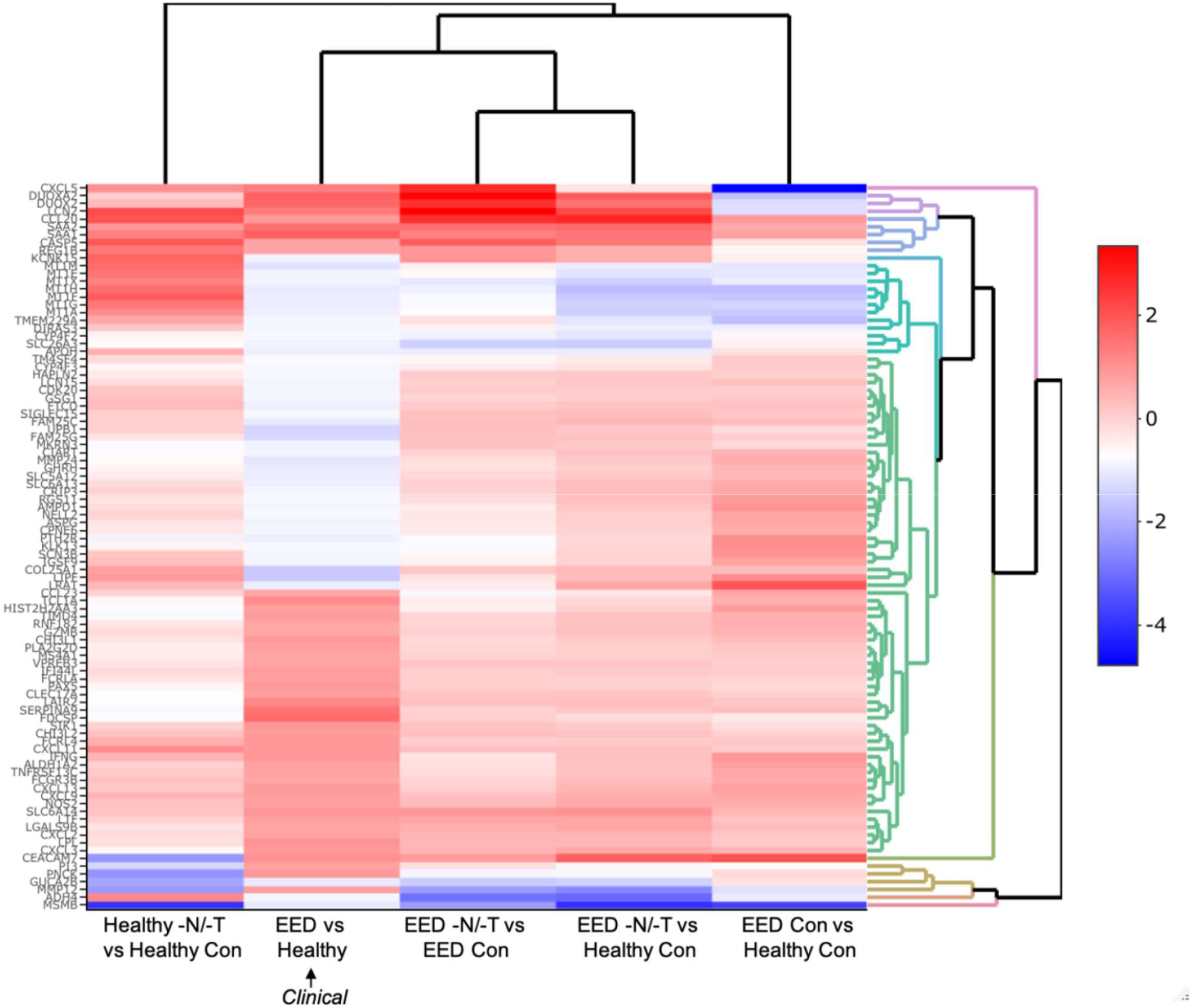
A comparison of the most up- and downregulated genes from the clinical EED signature with Healthy or EED Chip gene expression is depicted as a heatmap (red = upregulation, blue = downregulation) showing the expression pattern for EED Chips in -N/-T media vs Healthy Chips in control media. n = 3 chips for each condition.

**Supplementary Figure 10.**
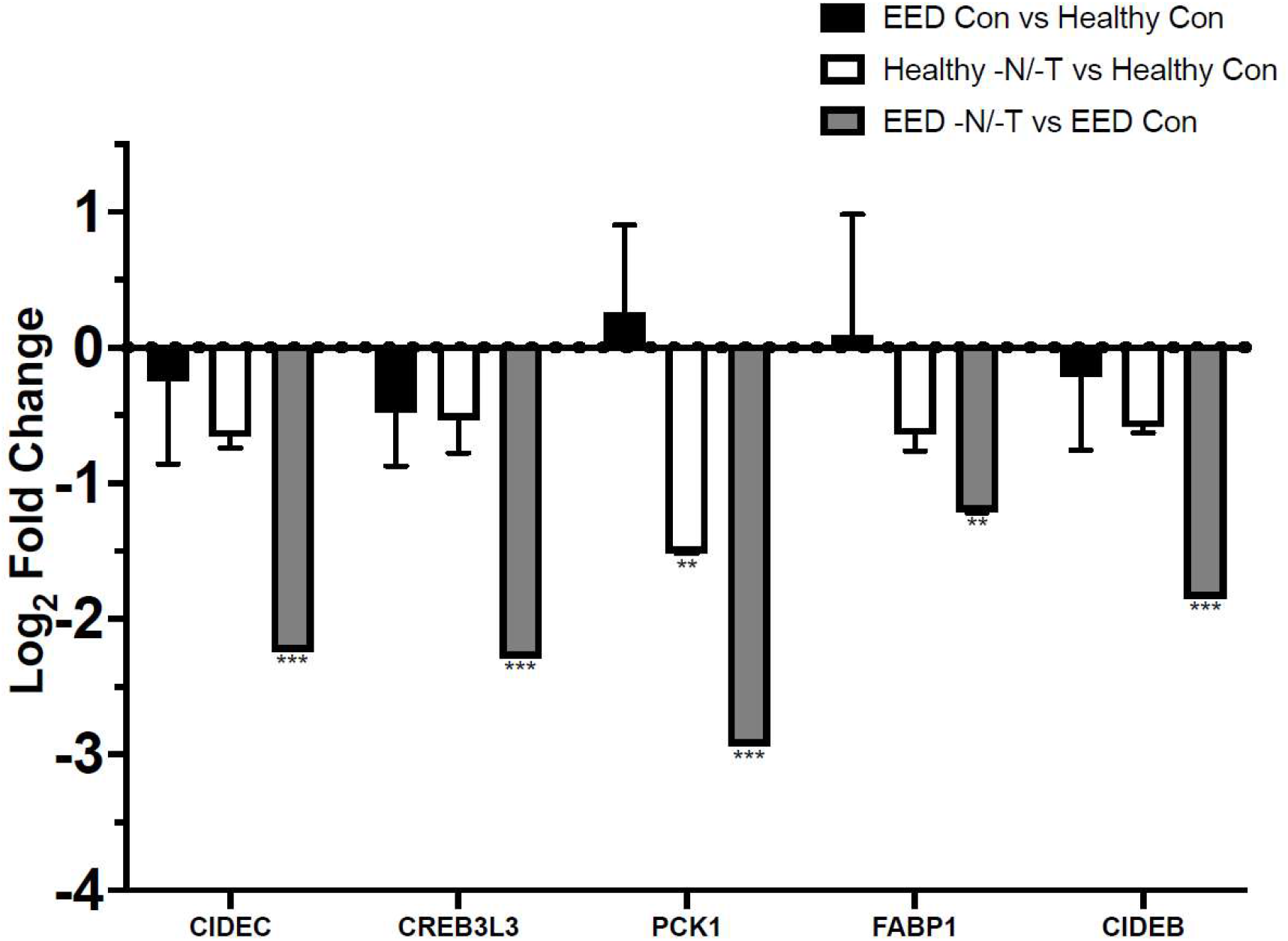
Transcriptional pathway analysis revealed a strong theme of downregulation for genes related to fatty acid absorption and processing when EED Chips were exposed to -N/-T media. This included a 4.7-fold downregulation of CIDEC (FDR < 0.0001), a 4.9-fold downregulation of CREB3L3 (FDR <0.001) and a 7.7-fold downregulation of PCK1 (FDR < 0.0001). n = 3 chips for each condition.

