## Supplementary Table 1 for "Nutritional deficiency recapitulates intestinal injury associated with environmental enteric dysfunction in patient-derived Organ Chips"

**EED Con vs Healthy Con**

| Gene Symbol | Entrez ID | Gene Name | log2 Fold Change | fdr p-value |
|---|---|---|---|---|
| GTSF1 | 121355 | gametocyte specific factor 1 | 4.41793983 | 1.10E-08 |
| NRG4 | 145957 | neuregulin 4 | 3.12293652 | 5.68E-06 |
| PAX8-AS1 | 654433 | PAX8 antisense RNA 1 | 2.98478243 | 4.13E-08 |
| LIN7A | 8825 | lin-7 homolog A, crumbs cell polarity co | 2.37283159 | 7.42E-05 |
| LINC01612 | 101928223 | long intergenic non-protein coding RNA | 2.16604493 | 0.00035298 |
| TPK1 | 27010 | thiamin pyrophosphokinase 1 | 2.00378543 | 7.42E-05 |
| MUC5AC | 4586 | mucin 5AC, oligomeric mucus/gel-form | 1.88649966 | 0.00154969 |
| KLK7 | 5650 | kallikrein related peptidase 7 | 1.88634197 | 3.97E-06 |
| LINC00470 | 56651 | long intergenic non-protein coding RNA | 1.83680982 | 0.00573765 |
| WASF3 | 10810 | WAS protein family member 3 | 1.83588184 | 0.0002455 |
| FGFBP1 | 9982 | fibroblast growth factor binding protei | 1.77181911 | 5.33E-05 |
| LRRTM1 | 347730 | leucine rich repeat transmembrane neu | 1.74864665 | 0.02874077 |
| RHBDL2 | 54933 | rhomboid like 2 | 1.73562738 | 1.82E-05 |
| DDX43 | 55510 | DEAD-box helicase 43 | 1.67361597 | 0.00086137 |
| NMU | 10874 | neuromedin U | 1.59025692 | 0.00621385 |
| ANO3 | 63982 | anoctamin 3 | 1.549945 | 0.00127621 |
| MAL | 4118 | mal, T cell differentiation protein | 1.51479921 | 0.01108006 |
| LSAMP | 4045 | limbic system associated membrane p | 1.4319161 | 0.01507643 |
| PPIL3 | 53938 | peptidylprolyl isomerase like 3 | 1.37696011 | 1.82E-05 |
| KIAA1324L | 222223 | KIAA1324 like | 1.3755227 | 0.00084403 |
| STOX1 | 219736 | storkhead box 1 | 1.31352735 | 0.04197635 |
| PTGR2 | 145482 | prostaglandin reductase 2 | 1.25204061 | 0.00017019 |
| RPL22L1 | 200916 | ribosomal protein L22 like 1 | 1.21651009 | 0.03356424 |
| ANO1 | 55107 | anoctamin 1 | 1.2118483 | 0.00158989 |
| LOXL1 | 4016 | lysyl oxidase like 1 | 1.19401316 | 0.00920896 |
| DMRTA1 | 63951 | DMRT like family A1 | 1.17942479 | 0.00547763 |
| TFF2 | 7032 | trefoil factor 2 | 1.16793288 | 0.00192117 |
| TNNC1 | 7134 | troponin C1, slow skeletal and cardiac t | 1.14613529 | 0.01512077 |
| EFNA1 | 1942 | ephrin A1 | 1.09930958 | 0.03289728 |
| KCNS3 | 3790 | potassium voltage-gated channel modi | 1.0620124 | 0.00030281 |
| SLC22A18AS | 5003 | solute carrier family 22 member 18 an | 1.05187831 | 0.00088552 |
| VGLL1 | 51442 | vestigial like family member 1 | 1.04815764 | 0.02722893 |
| MBOAT2 | 129642 | membrane bound O-acyltransferase do | 1.0338448 | 0.01112151 |
| PSCA | 8000 | prostate stem cell antigen | 1.02895018 | 0.00625832 |
| IL1R2 | 7850 | interleukin 1 receptor type 2 | 1.02351285 | 0.01876271 |
| TAS2R50 | 259296 | taste 2 receptor member 50 | 1.0083446 | 0.00845289 |
| SCGB2A1 | 4246 | secretoglobin family 2A member 1 | 0.98545257 | 0.04345233 |
| TDRD9 | 122402 | tudor domain containing 9 | 0.97333407 | 0.04927637 |
| S100A4 | 6275 | S100 calcium binding protein A4 | 0.96129845 | 0.00400244 |
| LUM | 4060 | lumican | 0.94612036 | 0.04197635 |
| AKR1E2 | 83592 | aldo-keto reductase family 1 member E | 0.94560323 | 0.00435715 |
| PRLR | 5618 | prolactin receptor | 0.93476547 | 0.00208963 |
| MUC20 | 200958 | mucin 20, cell surface associated | 0.92984964 | 0.01952194 |
| FABP12 | 646486 | fatty acid binding protein 12 | 0.91195454 | 0.01015691 |
| ABO | 28 | ABO, alpha 1-3-N-acetylgalactosaminy | 0.90755082 | 0.00594187 |
| NUDT2 | 318 | nudix hydrolase 2 | 0.87181384 | 0.0034493 |
| MIR4482 | 100616323 | microRNA 4482 | 0.87161899 | 0.02939543 |
| PROCR | 10544 | protein C receptor | 0.86009933 | 0.01214527 |
| SCARNA6 | 677772 | small Cajal body-specific RNA 6 | 0.85789833 | 0.00088552 |
| TDRP | 157695 | testis development related protein | 0.85397025 | 0.01431371 |
| ZNF626 | 199777 | zinc finger protein 626 | 0.84360423 | 0.02231447 |
| RASSF2 | 9770 | Ras association domain family membe | 0.83354822 | 0.0270896 |
| APIP | 51074 | APAF1 interacting protein | 0.82393041 | 0.01796505 |
| TMEM44-AS | 100507297 | TMEM44 antisense RNA 1 | 0.81432749 | 0.01831417 |
| SNORD100 | 594838 | small nucleolar RNA, C/D box 100 | 0.8109594 | 0.00085403 |
| PRMT8 | 56341 | protein arginine methyltransferase 8 | 0.81048877 | 0.0270896 |
| FAM90A1 | 55138 | family with sequence similarity 90 mer | 0.79435505 | 0.02631144 |
| SNORD123 | 100113384 | small nucleolar RNA, C/D box 123 | 0.78632376 | 0.04789339 |
| LRAT | 9227 | lecithin retinol acyltransferase | 0.78172839 | 0.00578872 |
| HMSD | 284293 | histocompatibility minor serpin domain | 0.77678607 | 0.03883982 |
| IFI27 | 3429 | interferon alpha inducible protein 27 | 0.77628042 | 0.00298835 |
| EDEM1 | 9695 | ER degradation enhancing alpha-mann | 0.76237503 | 0.02677005 |
| MVB12B | 89853 | multivesicular body subunit 12B | 0.75436655 | 0.02814819 |
| HRASLS2 | 54979 | HRAS like suppressor 2 | 0.75251072 | 0.00723793 |
| LOXL1-AS1 | 100287616 | LOXL1 antisense RNA 1 | 0.73585552 | 0.04269821 |
| ID1 | 3397 | inhibitor of DNA binding 1, HLH protein | 0.73433831 | 0.00847148 |
| TMPRSS3 | 64699 | transmembrane serine protease 3 | 0.72952633 | 0.0357315 |
| GSTO2 | 119391 | glutathione S-transferase omega 2 | 0.68387013 | 0.03738974 |
| GSTO1 | 9446 | glutathione S-transferase omega 1 | 0.67900013 | 0.00529258 |
| RNASEH2B | 79621 | ribonuclease H2 subunit B | 0.67289754 | 0.035878 |
| LTB | 4050 | lymphotoxin beta | 0.66906488 | 0.03757834 |

**Healthy -N/-T vs Healthy Con**

| Gene Symbol | Entrez ID | Gene Name | log2 Fold Change | fdr p-value |
|---|---|---|---|---|
| ALDH1L2 | 160428 | aldehyde dehydrogenase 1 famil | 3.08035169 | 2.17E-05 |
| SLC34A2 | 10568 | solute carrier family 34 member | 2.74018488 | 3.28E-05 |
| CBS | 875 | cystathionine-beta-synthase | 2.63670607 | 6.11E-06 |
| SLC7A5 | 8140 | solute carrier family 7 member 5 | 2.49469706 | 1.27E-05 |
| CHAC1 | 79094 | ChaC glutathione specific gamm | 2.45779746 | 2.73E-05 |
| GPR87 | 53836 | G protein-coupled receptor 87 | 2.42118487 | 0.00029568 |
| CTH | 1491 | cystathionine gamma-lyase | 2.30988468 | 2.17E-05 |
| STC2 | 8614 | stanniocalcin 2 | 2.23778545 | 4.18E-05 |
| PHGDH | 26227 | phosphoglycerate dehydrogenase | 2.08144874 | 2.17E-05 |
| SLC7A11 | 23657 | solute carrier family 7 member 1 | 2.06908589 | 2.17E-05 |
| PSAT1 | 29968 | phosphoserine aminotransferase | 1.99268486 | 2.17E-05 |
| ASNS | 440 | asparagine synthetase (glutamin | 1.96899283 | 0.0002635 |
| CACNA2D1 | 781 | calcium voltage-gated channel a | 1.91168203 | 0.00026353 |
| SLC16A7 | 9194 | solute carrier family 16 member | 1.83400879 | 0.0089459 |
| TRIB3 | 57761 | tribbles pseudokinase 3 | 1.83150638 | 2.17E-05 |
| SLC6A9 | 6536 | solute carrier family 6 member 9 | 1.78963919 | 3.28E-05 |
| CPE | 1363 | carboxypeptidase E | 1.78405554 | 0.00114511 |
| UNC93A | 54346 | unc-93 homolog A | 1.74236802 | 3.28E-05 |
| NMUR2 | 56923 | neuromedin U receptor 2 | 1.67014071 | 0.01428227 |
| PSTPIP2 | 9050 | proline-serine-threonine phospha | 1.66706736 | 0.0014969 |
| CNDP1 | 84735 | carnosine dipeptidase 1 | 1.62943183 | 0.00263169 |
| EFHC2 | 80258 | EF-hand domain containing 2 | 1.59438732 | 0.00776682 |
| YARS | 8565 | tyrosyl-tRNA synthetase | 1.56695697 | 7.41E-05 |
| SLC38A3 | 10991 | solute carrier family 38 member | 1.54941652 | 0.0013548 |
| UGT3A1 | 133688 | UDP glycosyltransferase family 3 | 1.5398462 | 0.00220868 |
| CASP1 | 834 | caspase 1 | 1.52538289 | 0.00274473 |
| TINAG | 27283 | tubulointerstitial nephritis antige | 1.52139025 | 0.02211568 |
| NR1H4 | 9971 | nuclear receptor subfamily 1 gro | 1.50768784 | 0.01868743 |
| MTHFD1L | 25902 | methylenetetrahydrofolate dehyd | 1.48923805 | 7.41E-05 |
| AJUBA | 84962 | ajuba LIM protein | 1.4644514 | 2.17E-05 |
| SORBS2 | 8470 | sorbin and SH3 domain containin | 1.41220694 | 0.0229408 |
| PCK2 | 5106 | phosphoenolpyruvate carboxykina | 1.39639368 | 0.00114511 |
| SLC1A5 | 6510 | solute carrier family 1 member 5 | 1.37282339 | 0.0002599 |
| VLDLR-AS1 | 401491 | VLDLR antisense RNA 1 | 1.36885218 | 0.00334678 |
| DDIT3 | 1649 | DNA damage inducible transcript | 1.36020178 | 0.00244751 |
| GARS | 2617 | glycyl-tRNA synthetase | 1.35796964 | 3.28E-05 |
| PTPRB | 5787 | protein tyrosine phosphatase, re | 1.35742881 | 0.00428372 |
| PPP1R1B | 84152 | protein phosphatase 1 regulatory | 1.35343256 | 8.27E-05 |
| IRAK2 | 3656 | interleukin 1 receptor associated | 1.33762918 | 6.58E-05 |
| SERPINB7 | 8710 | serpin family B member 7 | 1.33427254 | 0.00244751 |
| EIF4EBP1 | 1978 | eukaryotic translation initiation f | 1.3339503 | 0.00090126 |
| TBL1X | 6907 | transducin beta like 1 X-linked | 1.31060804 | 0.0002599 |
| IRF8 | 3394 | interferon regulatory factor 8 | 1.31027432 | 0.00024267 |
| SLC7A1 | 6541 | solute carrier family 7 member 1 | 1.30973845 | 0.00031413 |
| GSTM2 | 2946 | glutathione S-transferase mu 2 | 1.30297194 | 8.59E-05 |
| REG3A | 5068 | regenerating family member 3 a | 1.29890128 | 0.00047911 |
| SLC7A2 | 6542 | solute carrier family 7 member 2 | 1.29138248 | 0.0130601 |
| GSTM1 | 2944 | glutathione S-transferase mu 1 | 1.28994123 | 7.71E-05 |
| LURAP1L-AS1 | 101929467 | LURAP1L antisense RNA 1 | 1.28982237 | 0.0182953 |
| ACSM3 | 6296 | acyl-CoA synthetase medium cha | 1.28574323 | 0.0020531 |
| SLC1A4 | 6509 | solute carrier family 1 member 4 | 1.27918992 | 0.00058234 |
| LAMB1 | 3912 | laminin subunit beta 1 | 1.27903294 | 0.01252953 |
| ANXA13 | 312 | annexin A13 | 1.27258405 | 0.00204158 |
| EPB41L4A-AS | 114915 | EPB41L4A antisense RNA 1 | 1.27179272 | 0.01597138 |
| GPC4 | 2239 | glypican 4 | 1.27115366 | 0.00578861 |
| MTHFD2 | 10797 | methylenetetrahydrofolate dehyd | 1.26985188 | 7.71E-05 |
| TSC22D3 | 1831 | TSC22 domain family member 3 | 1.25969788 | 0.00238505 |
| VLDLR | 7436 | very low density lipoprotein rece | 1.24790908 | 0.0385069 |
| ANKRD30B | 374860 | ankyrin repeat domain 30B | 1.2453363 | 0.02352005 |
| AARS | 16 | alanyl-tRNA synthetase | 1.23123322 | 0.00021703 |
| SARS | 6301 | seryl-tRNA synthetase | 1.22959942 | 0.00021703 |
| GRB10 | 2887 | growth factor receptor bound pro | 1.22742499 | 0.00270845 |
| GSTM4 | 2948 | glutathione S-transferase mu 4 | 1.22110364 | 4.63E-05 |
| KCNJ3 | 3760 | potassium voltage-gated channe | 1.21240081 | 0.01330834 |
| EFNA5 | 1946 | ephrin A5 | 1.20925205 | 0.00065494 |
| IARS | 3376 | isoleucyl-tRNA synthetase | 1.20743396 | 0.00038205 |
| LCN2 | 3934 | lipocalin 2 | 1.19492939 | 0.00268158 |
| CDNF | 441549 | cerebral dopamine neurotrophic | 1.19043811 | 0.00334678 |
| SHMT2 | 6472 | serine hydroxymethyltransferase | 1.18486535 | 0.0004868 |
| BVES | 11149 | blood vessel epicardial substance | 1.17970442 | 0.01288736 |
| IFRD1 | 3475 | interferon related developmenta | 1.16854453 | 0.00194046 |

**EED -N/-T vs EED Con**

| Gene Symbol | Entrez ID | Gene Name | log2 Fold Change | fdr p-value |
|---|---|---|---|---|
| SMOC2 | 64094 | SPARC related modular calcium binding 2 | 4.48634349 | 0.020645 |
| CYP4X1 | 260293 | cytochrome P450 family 4 subfamily X mem | 3.15295227 | 0.00161934 |
| LCN2 | 3934 | lipocalin 2 | 2.71459683 | 1.26E-05 |
| DUOXA2 | 405753 | dual oxidase maturation factor 2 | 2.64785989 | 0.00451952 |
| CPVL | 54504 | carboxypeptidase, vitellogenic like | 2.39793383 | 0.00987488 |
| GPR87 | 53836 | G protein-coupled receptor 87 | 2.34034135 | 0.00022764 |
| CXCL5 | 6374 | C-X-C motif chemokine ligand 5 | 2.19376047 | 6.84E-05 |
| CASP1 | 834 | caspase 1 | 2.13068643 | 0.0001383 |
| ALDH1L2 | 160428 | aldehyde dehydrogenase 1 family member | 2.12030114 | 0.00017329 |
| CCL20 | 6364 | C-C motif chemokine ligand 20 | 2.11944603 | 4.76E-05 |
| NMUR2 | 56923 | neuromedin U receptor 2 | 2.1120437 | 0.00248926 |
| DUOX2 | 50506 | dual oxidase 2 | 2.10072554 | 0.0146251 |
| SLC7A5 | 8140 | solute carrier family 7 member 5 | 2.09728229 | 1.45E-05 |
| CHAC1 | 79094 | ChaC glutathione specific gamma-glutamylc | 2.06106263 | 6.89E-05 |
| SERPINA3 | 12 | serpin family A member 3 | 2.02221873 | 0.00017377 |
| BCAT1 | 586 | branched chain amino acid transaminase 1 | 2.00430496 | 0.00035662 |
| PHGDH | 26227 | phosphoglycerate dehydrogenase | 2.00192792 | 2.46E-05 |
| SORBS2 | 8470 | sorbin and SH3 domain containing 2 | 1.987657 | 0.00247061 |
| CBS | 875 | cystathionine-beta-synthase | 1.96868666 | 1.45E-05 |
| ASNS | 440 | asparagine synthetase (glutamine-hydrolyzi | 1.96248706 | 0.00015689 |
| SLC38A4 | 55089 | solute carrier family 38 member 4 | 1.93673855 | 0.00112841 |
| TOX | 9760 | thymocyte selection associated high mobilit | 1.90088652 | 0.00062814 |
| SLC34A2 | 10568 | solute carrier family 34 member 2 | 1.90070311 | 0.00033995 |
| LAMB1 | 3912 | laminin subunit beta 1 | 1.85472633 | 0.00080724 |
| STC2 | 8614 | stanniocalcin 2 | 1.80626433 | 0.0001383 |
| THBS2 | 7058 | thrombospondin 2 | 1.78276879 | 0.00088709 |
| PSAT1 | 29968 | phosphoserine aminotransferase 1 | 1.7288413 | 3.97E-05 |
| ANXA13 | 312 | annexin A13 | 1.6896341 | 0.0001383 |
| TGFBI | 7045 | transforming growth factor beta induced | 1.64928826 | 0.02388068 |
| PARM1 | 25849 | prostate androgen-regulated mucin-like pro | 1.64056859 | 0.00013484 |
| COLGALT2 | 23127 | collagen beta(1-O)galactosyltransferase 2 | 1.60155099 | 0.0249487 |
| LRRTM1 | 347730 | leucine rich repeat transmembrane neurona | 1.58167585 | 0.01803743 |
| SLC6A9 | 6536 | solute carrier family 6 member 9 | 1.56151935 | 6.89E-05 |
| SLC7A11 | 23657 | solute carrier family 7 member 11 | 1.5405555 | 9.88E-05 |
| CADPS | 8618 | calcium dependent secretion activator | 1.53791894 | 0.00012323 |
| CASP5 | 838 | caspase 5 | 1.52686645 | 0.0056327 |
| NME5 | 8382 | NME/NM23 family member 5 | 1.49073679 | 0.0382558 |
| CTH | 1491 | cystathionine gamma-lyase | 1.48429777 | 0.00035662 |
| CCL2 | 6347 | C-C motif chemokine ligand 2 | 1.4840763 | 0.00242605 |
| UNC93A | 54346 | unc-93 homolog A | 1.47696445 | 7.68E-05 |
| PRSS2 | 5645 | serine protease 2 | 1.45973088 | 1.26E-05 |
| PSTPIP2 | 9050 | proline-serine-threonine phosphatase intera | 1.44668341 | 0.00213768 |
| PPP1R1B | 84152 | protein phosphatase 1 regulatory inhibitor s | 1.44252837 | 4.76E-05 |
| ACSM3 | 6296 | acyl-CoA synthetase medium chain family m | 1.44034999 | 0.00052419 |
| PTPRB | 5787 | protein tyrosine phosphatase, receptor type | 1.42766734 | 0.00177135 |
| IARS | 3376 | isoleucyl-tRNA synthetase | 1.42312063 | 6.89E-05 |
| PDZK1IP1 | 10158 | PDZK1 interacting protein 1 | 1.42246434 | 0.00225348 |
| ALDH3A1 | 218 | aldehyde dehydrogenase 3 family member A | 1.41357403 | 0.00100252 |
| EPB41L4A-AS | 114915 | EPB41L4A antisense RNA 1 | 1.40945115 | 0.00634031 |
| GSTM4 | 2948 | glutathione S-transferase mu 4 | 1.39096057 | 1.45E-05 |
| CPE | 1363 | carboxypeptidase E | 1.39081403 | 0.00328806 |
| GSTM2 | 2946 | glutathione S-transferase mu 2 | 1.38646909 | 5.01E-05 |
| IFI16 | 3428 | interferon gamma inducible protein 16 | 1.38438889 | 0.00095263 |
| GAS5 | 60674 | growth arrest specific 5 (non-protein coding | 1.37822367 | 6.49E-05 |
| EFHC2 | 80258 | EF-hand domain containing 2 | 1.36732773 | 0.01164398 |
| BCL11A | 53335 | B cell CLL/lymphoma 11A | 1.33773031 | 0.03744559 |
| CACNA2D1 | 781 | calcium voltage-gated channel auxiliary sub | 1.33340509 | 0.00212874 |
| CFI | 3426 | complement factor I | 1.33156566 | 0.00574928 |
| SLFN5 | 162394 | schlafen family member 5 | 1.32475096 | 0.00038634 |
| CLIC6 | 54102 | chloride intracellular channel 6 | 1.3151349 | 0.00829425 |
| EPHB2 | 2048 | EPH receptor B2 | 1.31141496 | 0.01527915 |
| SAA2 | 6289 | serum amyloid A2 | 1.29652785 | 0.00194292 |
| AJUBA | 84962 | ajuba LIM protein | 1.27087559 | 2.94E-05 |
| AADAT | 51166 | aminoadipate aminotransferase | 1.26707446 | 0.0056327 |
| CNDP1 | 84735 | carnosine dipeptidase 1 | 1.26437429 | 0.00671843 |
| SOD3 | 6649 | superoxide dismutase 3 | 1.25977122 | 0.00264127 |
| ETV5 | 2119 | ETS variant 5 | 1.25157614 | 0.02604542 |
| MUC6 | 4588 | mucin 6, oligomeric mucus/gel-forming | 1.25112025 | 0.00429075 |
| ERV3-1 | 2086 | endogenous retrovirus group 3 member 1, e | 1.25094081 | 0.01136341 |
| TRIB3 | 57761 | tribbles pseudokinase 3 | 1.24249618 | 0.00022484 |
| CXCL6 | 6372 | C-X-C motif chemokine ligand 6 | 1.24144881 | 0.00052497 |

| | | | | |
|---|---|---|---|---|
| ZNF253 | 56242 | zinc finger protein 253 | 0.668627 | 0.02230624 |
| ADNP2 | 22850 | ADNP homeobox 2 | 0.66836105 | 0.03122628 |
| AIFM2 | 84883 | apoptosis inducing factor, mitochondria | 0.66231693 | 0.04550894 |
| VKORC1 | 79001 | vitamin K epoxide reductase complex s | 0.64798189 | 0.01010582 |
| SNORA65 | 26783 | small nucleolar RNA, H/ACA box 65 | 0.63618027 | 0.04197635 |
| MRPL23 | 6150 | mitochondrial ribosomal protein L23 | 0.63302795 | 0.04675753 |
| CCDC167 | 154467 | coiled-coil domain containing 167 | 0.62867055 | 0.02572743 |
| PLEK2 | 26499 | pleckstrin 2 | 0.62203855 | 0.00573765 |
| LYNX1-SLUR | 111188157 | LYNX1-SLURP2 readthrough | 0.61993487 | 0.02230624 |
| MSI1 | 4440 | musashi RNA binding protein 1 | 0.61538272 | 0.02694007 |
| ENTPD3-AS1 | 285266 | ENTPD3 antisense RNA 1 | 0.61366981 | 0.04927573 |
| ARL17B | 100506084 | ADP ribosylation factor like GTPase 17B | 0.6104499 | 0.04686783 |
| ENOSF1 | 55556 | enolase superfamily member 1 | 0.59964408 | 0.00621385 |
| MPP7 | 143098 | membrane palmitoylated protein 7 | 0.58517891 | 0.01214527 |
| SNORA71D | 677840 | small nucleolar RNA, H/ACA box 71D | 0.58497642 | 0.01475082 |
| RGPD4 | 285190 | RANBP2-like and GRIP domain contain | -0.5857676 | 0.03959624 |
| SYNPO | 11346 | synaptopodin | -0.5928603 | 0.02230624 |
| HIBCH | 26275 | 3-hydroxyisobutyryl-CoA hydrolase | -0.5953505 | 0.04711205 |
| SLC22A3 | 6581 | solute carrier family 22 member 3 | -0.596363 | 0.04398883 |
| GLIS3 | 169792 | GLIS family zinc finger 3 | -0.5976854 | 0.0357315 |
| STAP2 | 55620 | signal transducing adaptor family mem | -0.599028 | 0.00845289 |
| ABHD4 | 63874 | abhydrolase domain containing 4 | -0.6010948 | 0.01138367 |
| SLCO2B1 | 11309 | solute carrier organic anion transporter | -0.6021678 | 0.035878 |
| SLC28A3 | 64078 | solute carrier family 28 member 3 | -0.6041667 | 0.04943809 |
| RGPD6 | 729540 | RANBP2-like and GRIP domain contain | -0.6060312 | 0.03044073 |
| ANO9 | 338440 | anoctamin 9 | -0.6071596 | 0.04546514 |
| RAB3D | 9545 | RAB3D, member RAS oncogene family | -0.6085996 | 0.0341013 |
| ATP6V0A1 | 535 | ATPase H+ transporting V0 subunit a1 | -0.6094194 | 0.03356424 |
| CIITA | 4261 | class II major histocompatibility compl | -0.6098478 | 0.02079863 |
| CASC19 | 103021165 | cancer susceptibility 19 (non-protein co | -0.6103209 | 0.0325232 |
| KRT23 | 25984 | keratin 23 | -0.6142352 | 0.01928252 |
| SLC39A14 | 23516 | solute carrier family 39 member 14 | -0.6158999 | 0.03173279 |
| CTSH | 1512 | cathepsin H | -0.6182198 | 0.01216356 |
| ENAM | 10117 | enamelin | -0.6206703 | 0.03681681 |
| POU3F2 | 5454 | POU class 3 homeobox 2 | -0.6315745 | 0.03080136 |
| POU6F2-AS1 | 100861520 | POU6F2 antisense RNA 1 | -0.631734 | 0.04796362 |
| ABCA7 | 10347 | ATP binding cassette subfamily A mem | -0.6335189 | 0.00860224 |
| SERPINA6 | 866 | serpin family A member 6 | -0.6335306 | 0.0435178 |
| GLS | 2744 | glutaminase | -0.6340147 | 0.03668675 |
| ANO5 | 203859 | anoctamin 5 | -0.6386074 | 0.0386933 |
| AFAP1L2 | 84632 | actin filament associated protein 1 like | -0.640988 | 0.03738974 |
| KCNE3 | 10008 | potassium voltage-gated channel subfa | -0.6429688 | 0.02814819 |
| BLOC1S5-TXI | 100526836 | BLOC1S5-TXNDC5 readthrough (NMD c | -0.6480668 | 0.01966968 |
| SEMA3F | 6405 | semaphorin 3F | -0.6546697 | 0.0335239 |
| GSTM2 | 2946 | glutathione S-transferase mu 2 | -0.6551581 | 0.01966968 |
| CFB | 629 | complement factor B | -0.6562668 | 0.00772465 |
| ADGRE5 | 976 | adhesion G protein-coupled receptor E5 | -0.6582561 | 0.00782629 |
| NBPF1 | 55672 | NBPF member 1 | -0.6599492 | 0.01102652 |
| PLEKHG5 | 57449 | pleckstrin homology and RhoGEF doma | -0.662363 | 0.03881281 |
| PPT2 | 9374 | palmitoyl-protein thioesterase 2 | -0.6626372 | 0.03189816 |
| HHLA1 | 10086 | HERV-H LTR-associating 1 | -0.6645835 | 0.02079863 |
| METTL7B | 196410 | methyltransferase like 7B | -0.6652334 | 0.00625832 |
| PDE4C | 5143 | phosphodiesterase 4C | -0.6659495 | 0.02230624 |
| KLK10 | 5655 | kallikrein related peptidase 10 | -0.6688031 | 0.02873255 |
| FAR2 | 55711 | fatty acyl-CoA reductase 2 | -0.6705745 | 0.02004453 |
| CDHR5 | 53841 | cadherin related family member 5 | -0.6795516 | 0.03912011 |
| TIMP1 | 7076 | TIMP metallopeptidase inhibitor 1 | -0.6810235 | 0.03959624 |
| ADAMTS9 | 56999 | ADAM metallopeptidase with thrombo | -0.6811933 | 0.04546514 |
| LRG1 | 116844 | leucine rich alpha-2-glycoprotein 1 | -0.6815102 | 0.01062902 |
| DHRS4-AS1 | 55449 | DHRS4 antisense RNA 1 | -0.6921064 | 0.04565479 |
| ARHGEF35 | 445328 | Rho guanine nucleotide exchange facto | -0.6943434 | 0.01342092 |
| SPTLC3 | 55304 | serine palmitoyltransferase long chain | -0.6955085 | 0.00724541 |
| ZNF829 | 374899 | zinc finger protein 829 | -0.6956072 | 0.04367796 |
| MT1X | 4501 | metallothionein 1X | -0.7012308 | 0.02079863 |
| DNAH3 | 55567 | dynein axonemal heavy chain 3 | -0.7126973 | 0.04565868 |
| CNKSR1 | 10256 | connector enhancer of kinase suppress | -0.712907 | 0.02382858 |
| TXNDC5 | 81567 | thioredoxin domain containing 5 | -0.7166532 | 0.02399044 |
| GSTM4 | 2948 | glutathione S-transferase mu 4 | -0.7175126 | 0.00384047 |
| RNF130 | 55819 | ring finger protein 130 | -0.7179063 | 0.01704716 |
| NPC1L1 | 29881 | NPC1 like intracellular cholesterol trans | -0.7186945 | 0.01966968 |
| CELF2 | 10659 | CUGBP Elav-like family member 2 | -0.71891 | 0.03044073 |
| PHLDB2 | 90102 | pleckstrin homology like domain family | -0.7210859 | 0.04369222 |
| FAIM | 55179 | Fas apoptotic inhibitory molecule | -0.7222386 | 0.02404967 |
| STEAP3 | 55240 | STEAP3 metalloreductase | -0.7222582 | 0.02004453 |

| | | | | |
|---|---|---|---|---|
| NAALADL2 | 254827 | N-acetylated alpha-linked acidic | 1.16845636 | 0.00336931 |
| XPOT | 11260 | exportin for tRNA | 1.16212099 | 0.00090947 |
| RCN1 | 5954 | reticulocalbin 1 | 1.16166131 | 4.74E-05 |
| MEF2C | 4208 | myocyte enhancer factor 2C | 1.15672972 | 0.00502614 |
| NEB | 4703 | nebulin | 1.1505149 | 0.02096246 |
| JMY | 133746 | junction mediating and regulator | 1.15047976 | 0.00693113 |
| SLC3A2 | 6520 | solute carrier family 3 member 2 | 1.14971744 | 0.00038205 |
| ALDH3A1 | 218 | aldehyde dehydrogenase 3 family | 1.14934698 | 0.00620708 |
| MSI1 | 4440 | musashi RNA binding protein 1 | 1.14880652 | 0.0002599 |
| SLC43A1 | 8501 | solute carrier family 43 member | 1.14217642 | 0.00304581 |
| SERPINA3 | 12 | serpin family A member 3 | 1.13648299 | 0.01258945 |
| ALKBH8 | 91801 | alkB homolog 8, tRNA methyltra | 1.13110067 | 0.00828114 |
| FBXO25 | 26260 | F-box protein 25 | 1.12463532 | 0.00204344 |
| CCL20 | 6364 | C-C motif chemokine ligand 20 | 1.12072856 | 0.00578861 |
| EXOSC8 | 11340 | exosome component 8 | 1.11117074 | 0.00293345 |
| FAM27E3 | 100131997 | family with sequence similarity 2 | 1.11018399 | 0.01252953 |
| CARS | 833 | cysteinyl-tRNA synthetase | 1.10927735 | 0.00069814 |
| KCNIP3 | 30818 | potassium voltage-gated channe | 1.10135956 | 0.03992953 |
| CASP5 | 838 | caspase 5 | 1.08689223 | 0.03939784 |
| SNORD12C | 26765 | small nucleolar RNA, C/D box 12 | 1.07786226 | 0.00090947 |
| ZNF25 | 219749 | zinc finger protein 25 | 1.07541485 | 0.0040642 |
| SCGN | 10590 | secretagogin, EF-hand calcium b | 1.0733444 | 0.00961469 |
| EPRS | 2058 | glutamyl-prolyl-tRNA synthetase | 1.07233582 | 0.00066341 |
| ZBTB10 | 65986 | zinc finger and BTB domain cont | 1.07215462 | 0.00410122 |
| DSG3 | 1830 | desmoglein 3 | 1.06518964 | 0.03190712 |
| NUDT19 | 390916 | nudix hydrolase 19 | 1.06342195 | 0.02077962 |
| WARS | 7453 | tryptophanyl-tRNA synthetase | 1.0604578 | 0.00136706 |
| THSD4 | 79875 | thrombospondin type 1 domain c | 1.05967074 | 0.01428227 |
| METAP1D | 254042 | methionyl aminopeptidase type 1 | 1.05720972 | 0.00475156 |
| CADPS | 8618 | calcium dependent secretion act | 1.04861745 | 0.00317002 |
| EPHB2 | 2048 | EPH receptor B2 | 1.04827167 | 0.04952039 |
| TPD52L1 | 7164 | tumor protein D52 like 1 | 1.04530156 | 0.00273332 |
| MT1F | 4494 | metallothionein 1F | 1.03973992 | 0.00961469 |
| MAOA | 4128 | monoamine oxidase A | 1.02220766 | 0.02306633 |
| ST6GALNAC3 | 256435 | ST6 N-acetylgalactosaminide alp | 1.014046 | 0.01262385 |
| FCHSD2 | 9873 | FCH and double SH3 domains 2 | 1.01223421 | 0.01568881 |
| GPT2 | 84706 | glutamic--pyruvic transaminase 2 | 1.01208169 | 0.00114511 |
| LINC01612 | 101928223 | long intergenic non-protein codir | 1.0114463 | 0.0264936 |
| PDGFA | 5154 | platelet derived growth factor su | 1.0106597 | 0.00578988 |
| ARSE | 415 | arylsulfatase E (chondrodysplasia | 1.01044394 | 0.02536567 |
| SH2B3 | 10019 | SH2B adaptor protein 3 | 1.00758288 | 0.00696825 |
| SNTB1 | 6641 | syntrophin beta 1 | 1.00701349 | 0.00578861 |
| PNPLA1 | 285848 | patatin like phospholipase doma | 1.00643063 | 0.01780573 |
| PTPN14 | 5784 | protein tyrosine phosphatase, no | 0.99668753 | 0.00038205 |
| SNORD50A | 26799 | small nucleolar RNA, C/D box 50 | 0.99266132 | 0.0130601 |
| MAP2K5 | 5607 | mitogen-activated protein kinase | 0.99111675 | 0.00089839 |
| CPA2 | 1358 | carboxypeptidase A2 | 0.98761245 | 0.03337349 |
| MOCS1 | 4337 | molybdenum cofactor synthesis 1 | 0.98749824 | 0.00578988 |
| KLHL5 | 51088 | kelch like family member 5 | 0.98417659 | 0.01137687 |
| SESN2 | 83667 | sestrin 2 | 0.98328674 | 0.00100837 |
| CCND2 | 894 | cyclin D2 | 0.98236338 | 0.00054987 |
| KCNK15 | 60598 | potassium two pore domain char | 0.98160309 | 0.01790219 |
| CEP126 | 57562 | centrosomal protein 126 | 0.9798946 | 0.03985085 |
| SCRN1 | 9805 | secernin 1 | 0.97928993 | 0.01045399 |
| BCAT1 | 586 | branched chain amino acid trans | 0.9792507 | 0.03347772 |
| LDLRAD3 | 143458 | low density lipoprotein receptor | 0.97910068 | 0.0046388 |
| GAS5 | 60674 | growth arrest specific 5 (non-pro | 0.97870913 | 0.00114511 |
| LRPPRC | 10128 | leucine rich pentatricopeptide re | 0.97634592 | 0.00114511 |
| MOCOS | 55034 | molybdenum cofactor sulfurase | 0.97575628 | 0.00318161 |
| PRKAR1B | 5575 | protein kinase cAMP-dependent | 0.97575506 | 0.00189106 |
| PAPSS2 | 9060 | 3'-phosphoadenosine 5'-phospho | 0.97318966 | 0.04303136 |
| ACKR4 | 51554 | atypical chemokine receptor 4 | 0.97241275 | 0.00148065 |
| ANO1 | 55107 | anoctamin 1 | 0.97119961 | 0.00500975 |
| SNORD1A | 677848 | small nucleolar RNA, C/D box 1A | 0.96350387 | 0.00576562 |
| CLIC4 | 25932 | chloride intracellular channel 4 | 0.9621097 | 0.00273332 |
| PDE3B | 5140 | phosphodiesterase 3B | 0.95642141 | 0.01756491 |
| RHOQ | 23433 | ras homolog family member Q | 0.95588489 | 0.01522005 |
| PPP1R36 | 145376 | protein phosphatase 1 regulatory | 0.94892844 | 0.00915944 |
| EDA2R | 60401 | ectodysplasin A2 receptor | 0.94802775 | 0.01787727 |
| RIN3 | 79890 | Ras and Rab interactor 3 | 0.94491367 | 0.01627355 |
| DDIT4 | 54541 | DNA damage inducible transcript | 0.93977453 | 0.00861386 |
| RASGRF2 | 5924 | Ras protein specific guanine nucl | 0.93860563 | 0.03021069 |
| WDR63 | 126820 | WD repeat domain 63 | 0.93769302 | 0.02997331 |
| FZD6 | 8323 | frizzled class receptor 6 | 0.93649769 | 0.03004045 |

| | | | | |
|---|---|---|---|---|
| GSTM1 | 2944 | glutathione S-transferase mu 1 | 1.2312683 | 7.58E-05 |
| GARS | 2617 | glycyl-tRNA synthetase | 1.20923113 | 6.49E-05 |
| CXCL14 | 9547 | C-X-C motif chemokine ligand 14 | 1.20392719 | 0.02422437 |
| C3 | 718 | complement C3 | 1.20362162 | 0.00029253 |
| MTHFD1L | 25902 | methylenetetrahydrofolate dehydrogenase | 1.20193006 | 0.00024374 |
| EIF4EBP1 | 1978 | eukaryotic translation initiation factor 4E bi | 1.20100621 | 0.001042 |
| SLC1A5 | 6510 | solute carrier family 1 member 5 | 1.1951721 | 0.00041762 |
| TMEM150C | 441027 | transmembrane protein 150C | 1.18358352 | 0.01684697 |
| TPD52L1 | 7164 | tumor protein D52 like 1 | 1.18306205 | 0.00065434 |
| ASIC1 | 41 | acid sensing ion channel subunit 1 | 1.17932696 | 0.00178747 |
| VLDLR-AS1 | 401491 | VLDLR antisense RNA 1 | 1.17570513 | 0.00481385 |
| GRIN2B | 2904 | glutamate ionotropic receptor NMDA type s | 1.17350825 | 0.04555666 |
| YARS | 8565 | tyrosyl-tRNA synthetase | 1.17193423 | 0.00043388 |
| WARS | 7453 | tryptophanyl-tRNA synthetase | 1.17106554 | 0.00036503 |
| STK32A | 202374 | serine/threonine kinase 32A | 1.16341082 | 0.01245643 |
| SLC7A2 | 6542 | solute carrier family 7 member 2 | 1.158475 | 0.01597283 |
| TRAF5 | 7188 | TNF receptor associated factor 5 | 1.15624667 | 0.00282706 |
| DRAM1 | 55332 | DNA damage regulated autophagy modulat | 1.14826813 | 0.00079302 |
| ADGRG1 | 9289 | adhesion G protein-coupled receptor G1 | 1.14696157 | 0.00044144 |
| SDC3 | 9672 | syndecan 3 | 1.13587272 | 0.00407527 |
| ZNF532 | 55205 | zinc finger protein 532 | 1.12801006 | 0.03596635 |
| SLC1A4 | 6509 | solute carrier family 1 member 4 | 1.12164701 | 0.00083444 |
| KRT73-AS1 | 100127967 | KRT73 antisense RNA 1 | 1.12161614 | 0.03056874 |
| NFE2L3 | 9603 | nuclear factor, erythroid 2 like 3 | 1.11398307 | 0.00471369 |
| RIN3 | 79890 | Ras and Rab interactor 3 | 1.10885609 | 0.00454239 |
| ALDH1B1 | 219 | aldehyde dehydrogenase 1 family member | 1.10704803 | 0.00740245 |
| GSTM5 | 2949 | glutathione S-transferase mu 5 | 1.10281642 | 0.0001413 |
| SNORD12C | 26765 | small nucleolar RNA, C/D box 12C | 1.09539694 | 0.00044144 |
| MAP2K6 | 5608 | mitogen-activated protein kinase kinase 6 | 1.09093531 | 0.00656663 |
| XPOT | 11260 | exportin for tRNA | 1.08224453 | 0.00083444 |
| IRF8 | 3394 | interferon regulatory factor 8 | 1.07197148 | 0.00062313 |
| HNRNPA1L2 | 144983 | heterogeneous nuclear ribonucleoprotein A1 | 1.06848544 | 0.04556628 |
| CYP4F11 | 57834 | cytochrome P450 family 4 subfamily F mem | 1.06347849 | 0.01565592 |
| FBXO25 | 26260 | F-box protein 25 | 1.06131525 | 0.00165453 |
| EPRS | 2058 | glutamyl-prolyl-tRNA synthetase | 1.06119413 | 0.00039855 |
| DZIP1L | 199221 | DAZ interacting zinc finger protein 1 like | 1.0608734 | 0.02731519 |
| DOCK11 | 139818 | dedicator of cytokinesis 11 | 1.06034522 | 0.03571338 |
| TAF4B | 6875 | TATA-box binding protein associated factor | 1.05977326 | 0.01374743 |
| RPS6KA2 | 6196 | ribosomal protein S6 kinase A2 | 1.05573669 | 0.00278167 |
| RCN1 | 5954 | reticulocalbin 1 | 1.05393832 | 6.89E-05 |
| TCEA3 | 6920 | transcription elongation factor A3 | 1.04490418 | 0.00057541 |
| HMGCS2 | 3158 | 3-hydroxy-3-methylglutaryl-CoA synthase 2 | 1.04279355 | 0.01289253 |
| ATP6V1E2 | 90423 | ATPase H+ transporting V1 subunit E2 | 1.04091296 | 0.00643942 |
| PTK7 | 5754 | protein tyrosine kinase 7 (inactive) | 1.04001662 | 0.01575666 |
| SAA1 | 6288 | serum amyloid A1 | 1.03980447 | 0.01712561 |
| GAB2 | 9846 | GRB2 associated binding protein 2 | 1.03587912 | 0.00468319 |
| SCRN1 | 9805 | secernin 1 | 1.01820821 | 0.00521077 |
| CLUAP1 | 23059 | clusterin associated protein 1 | 1.01537891 | 0.00068172 |
| LDLRAD3 | 143458 | low density lipoprotein receptor class A don | 1.0116512 | 0.00216681 |
| NAALADL2 | 254827 | N-acetylated alpha-linked acidic dipeptidase | 1.01020704 | 0.00470812 |
| DBN1 | 1627 | drebrin 1 | 1.00911548 | 0.00072374 |
| IRAK2 | 3656 | interleukin 1 receptor associated kinase 2 | 1.00284191 | 0.00037487 |
| CFAP70 | 118491 | cilia and flagella associated protein 70 | 1.00183663 | 0.00920027 |
| PRKCG | 5582 | protein kinase C gamma | 1.00140145 | 0.00080724 |
| SNORD50A | 26799 | small nucleolar RNA, C/D box 50A | 0.99926821 | 0.00859144 |
| GPT2 | 84706 | glutamic--pyruvic transaminase 2 | 0.99737686 | 0.00071695 |
| FCHSD2 | 9873 | FCH and double SH3 domains 2 | 0.99646682 | 0.01220602 |
| FAM124B | 79843 | family with sequence similarity 124 membe | 0.99624241 | 0.01977039 |
| FAM129A | 116496 | family with sequence similarity 129 membe | 0.99489385 | 0.00222082 |
| MYB | 4602 | MYB proto-oncogene, transcription factor | 0.99291049 | 0.0436447 |
| KCNH2 | 3757 | potassium voltage-gated channel subfamily | 0.97998168 | 0.00803446 |
| DSG3 | 1830 | desmoglein 3 | 0.97786833 | 0.03697445 |
| SLC38A1 | 81539 | solute carrier family 38 member 1 | 0.97383988 | 0.00029253 |
| EDA2R | 60401 | ectodysplasin A2 receptor | 0.97372299 | 0.01128153 |
| SARS | 6301 | seryl-tRNA synthetase | 0.97054498 | 0.00072755 |
| LINC01347 | 731275 | long intergenic non-protein coding RNA 134 | 0.97020116 | 0.00874428 |
| ETS2 | 2114 | ETS proto-oncogene 2, transcription factor | 0.96987476 | 0.00242605 |
| ROR1 | 4919 | receptor tyrosine kinase like orphan recepto | 0.96943669 | 0.00399457 |
| DMD | 1756 | dystrophin | 0.96760454 | 0.03848784 |
| MTHFD2 | 10797 | methylenetetrahydrofolate dehydrogenase | 0.96663336 | 0.00041331 |
| LRRN1 | 57633 | leucine rich repeat neuronal 1 | 0.96500084 | 0.00648967 |
| TNFAIP2 | 7127 | TNF alpha induced protein 2 | 0.96436751 | 0.00039855 |
| GNA14 | 9630 | G protein subunit alpha 14 | 0.9540965 | 0.00265071 |
| REG3A | 5068 | regenerating family member 3 alpha | 0.95111652 | 0.00242605 |

| | | | | |
|---|---|---|---|---|
| FAM198B | 51313 | family with sequence similarity 198 m | -0.7241571 | 0.00860224 |
| ENOX2 | 10495 | ecto-NOX disulfide-thiol exchanger 2 | -0.7291533 | 0.035878 |
| AQP7 | 364 | aquaporin 7 | -0.7326773 | 0.02631375 |
| LRPAP1 | 4043 | LDL receptor related protein associated | -0.7386115 | 0.00576376 |
| CHRAC1 | 54108 | chromatin accessibility complex 1 | -0.7394916 | 0.00514603 |
| TTR | 7276 | transthyretin | -0.7395433 | 0.03788852 |
| GLRB | 2743 | glycine receptor beta | -0.7403241 | 0.04823551 |
| TEX9 | 374618 | testis expressed 9 | -0.7451101 | 0.04789339 |
| CPT1B | 1375 | carnitine palmitoyltransferase 1B | -0.7454951 | 0.04269821 |
| MT1M | 4499 | metallothionein 1M | -0.7542781 | 0.04388234 |
| GNAZ | 2781 | G protein subunit alpha z | -0.7599237 | 0.04245187 |
| NQO2 | 4835 | N-ribosyldihydronicotinamide:quinone | -0.7680005 | 0.00860224 |
| GLIPR1 | 11010 | GLI pathogenesis related 1 | -0.7696399 | 0.03300677 |
| SAPCD1-AS1 | 104413891 | SAPCD1 antisense RNA 1 | -0.7824771 | 0.00578872 |
| ENPP7 | 339221 | ectonucleotide pyrophosphatase/phosp | -0.7832326 | 0.04510983 |
| UNC93A | 54346 | unc-93 homolog A | -0.7839681 | 0.01723965 |
| DDX11L2 | 84771 | DEAD/H-box helicase 11 like 2 | -0.7855532 | 0.04228551 |
| B3GNT7 | 93010 | UDP-GlcNAc:betaGal beta-1,3-N-acety | -0.7932578 | 0.0033726 |
| CSF1 | 1435 | colony stimulating factor 1 | -0.7975355 | 0.00292153 |
| CATSPERB | 79820 | cation channel sperm associated auxili | -0.7994522 | 0.00158989 |
| MLXIPL | 51085 | MLX interacting protein like | -0.7996988 | 0.04627729 |
| LINC00417 | 100874164 | long intergenic non-protein coding RNA | -0.8064693 | 0.00643867 |
| FBXO27 | 126433 | F-box protein 27 | -0.8125384 | 0.00573765 |
| LETM2 | 137994 | leucine zipper and EF-hand containing | -0.8128154 | 0.02530861 |
| ADAM28 | 10863 | ADAM metallopeptidase domain 28 | -0.8185934 | 0.00204872 |
| DNAJA4 | 55466 | DnaJ heat shock protein family (Hsp40) | -0.8193794 | 0.02382858 |
| SSTR1 | 6751 | somatostatin receptor 1 | -0.8256895 | 0.00285547 |
| MIR3662 | 100500880 | microRNA 3662 | -0.8292129 | 0.04890784 |
| ACCS | 84680 | 1-aminocyclopropane-1-carboxylate sy | -0.8301623 | 0.02939543 |
| LCN2 | 3934 | lipocalin 2 | -0.8321147 | 0.03396343 |
| KLK1 | 3816 | kallikrein 1 | -0.8451087 | 0.0087324 |
| LINC01363 | 101928484 | long intergenic non-protein coding RNA | -0.8459353 | 0.01966968 |
| NBPF9 | 400818 | NBPF member 9 | -0.8524776 | 0.03598858 |
| POU2F3 | 25833 | POU class 2 homeobox 3 | -0.8586509 | 0.01704716 |
| NBR2 | 10230 | neighbor of BRCA1 gene 2 (non-protei | -0.8668901 | 0.00175681 |
| CYP3A7 | 1551 | cytochrome P450 family 3 subfamily A | -0.8727416 | 0.0307449 |
| PYGL | 5836 | glycogen phosphorylase L | -0.8800073 | 0.00723793 |
| TMEM253 | 643382 | transmembrane protein 253 | -0.8801948 | 0.00350706 |
| KCTD21-AS1 | 100289388 | KCTD21 antisense RNA 1 | -0.8831598 | 0.00446717 |
| CAPN3 | 825 | calpain 3 | -0.8837457 | 0.00761744 |
| REG3A | 5068 | regenerating family member 3 alpha | -0.8843374 | 0.0099524 |
| RCAN2 | 10231 | regulator of calcineurin 2 | -0.9025192 | 0.01424431 |
| XKR9 | 389668 | XK related 9 | -0.9042769 | 0.01950239 |
| CYP4V2 | 285440 | cytochrome P450 family 4 subfamily V | -0.9129285 | 0.00319437 |
| PRRT3-AS1 | 100874032 | PRRT3 antisense RNA 1 | -0.9146809 | 0.00298835 |
| SNORD14A | 26822 | small nucleolar RNA, C/D box 14A | -0.9151929 | 0.02572743 |
| FAS | 355 | Fas cell surface death receptor | -0.9249071 | 0.00818411 |
| MIR487A | 619555 | microRNA 487a | -0.925463 | 0.02364315 |
| CLU | 1191 | clusterin | -0.9259675 | 0.02814819 |
| TPPP | 11076 | tubulin polymerization promoting prote | -0.9267572 | 0.00085403 |
| DHRS7 | 51635 | dehydrogenase/reductase 7 | -0.9283408 | 0.00252983 |
| ST6GALNAC3 | 256435 | ST6 N-acetylgalactosaminide alpha-2,6 | -0.9301093 | 0.035878 |
| TAPBPL | 55080 | TAP binding protein like | -0.9329288 | 0.00512693 |
| NT5C3B | 115024 | 5'-nucleotidase, cytosolic IIIB | -0.9426682 | 0.00033023 |
| CLOCK | 9575 | clock circadian regulator | -0.9463106 | 0.04228551 |
| CLPTM1 | 1209 | CLPTM1, transmembrane protein | -0.9471087 | 0.0034493 |
| SORD | 6652 | sorbitol dehydrogenase | -0.9630053 | 0.00104966 |
| NABP1 | 64859 | nucleic acid binding protein 1 | -0.9717105 | 0.02830921 |
| PDPR | 55066 | pyruvate dehydrogenase phosphatase r | -0.9743154 | 0.00350706 |
| MT1A | 4489 | metallothionein 1A | -0.9850724 | 0.04686783 |
| XKRX | 402415 | XK related, X-linked | -0.9908661 | 0.01214527 |
| ZPLD1 | 131368 | zona pellucida like domain containing 1 | -0.9964158 | 0.02426736 |
| PIGR | 5284 | polymeric immunoglobulin receptor | -1.0028728 | 0.00030281 |
| HTR1D | 3352 | 5-hydroxytryptamine receptor 1D | -1.0083755 | 0.02814819 |
| ARSJ | 79642 | arylsulfatase family member J | -1.0196878 | 0.04197635 |
| MT1F | 4494 | metallothionein 1F | -1.0232378 | 0.01896965 |
| MAF | 4094 | MAF bZIP transcription factor | -1.0271778 | 0.04565868 |
| PRSS2 | 5645 | serine protease 2 | -1.0393026 | 0.00017095 |
| NPY4R | 5540 | neuropeptide Y receptor Y4 | -1.0519103 | 0.00413421 |
| ST3GAL1 | 6482 | ST3 beta-galactoside alpha-2,3-sialyltr | -1.0623097 | 0.00104966 |
| TMEM229A | 730130 | transmembrane protein 229A | -1.0650344 | 0.04565868 |
| ADH6 | 130 | alcohol dehydrogenase 6 (class V) | -1.0678659 | 0.00581797 |
| ZNF704 | 619279 | zinc finger protein 704 | -1.0729972 | 0.00319437 |
| MT1H | 4496 | metallothionein 1H | -1.0893761 | 0.02230624 |

| | | | | |
|---|---|---|---|---|
| PSD4 | 23550 | pleckstrin and Sec7 domain cont | 0.93183896 | 0.01514225 |
| PITPNM3 | 83394 | PITPNM family member 3 | 0.92780961 | 0.00041406 |
| MT1M | 4499 | metallothionein 1M | 0.9240359 | 0.00885861 |
| H6PD | 9563 | hexose-6-phosphate dehydrogen | 0.92307168 | 0.00389187 |
| BIRC3 | 330 | baculoviral IAP repeat containing | 0.92033428 | 0.01288736 |
| LINC00266-1 | 140849 | long intergenic non-protein codir | 0.91906949 | 0.04010817 |
| LONRF1 | 91694 | LON peptidase N-terminal doma | 0.91699294 | 0.00406827 |
| LARS | 51520 | leucyl-tRNA synthetase | 0.91447391 | 0.00083332 |
| ATP8A1 | 10396 | ATPase phospholipid transportin | 0.91229747 | 0.00449329 |
| MT1H | 4496 | metallothionein 1H | 0.91144795 | 0.02622626 |
| GSTZ1 | 2954 | glutathione S-transferase zeta 1 | 0.90893457 | 0.00766691 |
| KIF21B | 23046 | kinesin family member 21B | 0.90642042 | 0.00204158 |
| G0S2 | 50486 | G0/G1 switch 2 | 0.90417821 | 0.02282916 |
| SLC25A36 | 55186 | solute carrier family 25 member | 0.90244343 | 0.01780573 |
| ANK1 | 286 | ankyrin 1 | 0.90203099 | 0.0083859 |
| AIFM2 | 84883 | apoptosis inducing factor, mitoc | 0.89884674 | 0.00538163 |
| ATF4 | 468 | activating transcription factor 4 | 0.89734827 | 0.00068564 |
| NTF3 | 4908 | neurotrophin 3 | 0.89711572 | 0.02824866 |
| VWA2 | 340706 | von Willebrand factor A domain | 0.89129709 | 0.01328335 |
| IDH2 | 3418 | isocitrate dehydrogenase (NADP | 0.88853456 | 0.00274473 |
| TARS | 6897 | threonyl-tRNA synthetase | 0.88516407 | 0.00353101 |
| MAT1A | 4143 | methionine adenosyltransferase | 0.88375693 | 0.01749727 |
| SERPINB2 | 5055 | serpin family B member 2 | 0.88363255 | 0.02495778 |
| LINC00886 | 730091 | long intergenic non-protein codir | 0.88307601 | 0.01077355 |
| KBTBD11-OT | 104266957 | KBTBD11 overlapping transcript | 0.88018366 | 0.01189168 |
| SLC38A1 | 81539 | solute carrier family 38 member | 0.8792529 | 0.00114511 |
| COX18 | 285521 | COX18, cytochrome c oxidase ass | 0.87746646 | 0.03791723 |
| WAC-AS1 | 220906 | WAC antisense RNA 1 (head to h | 0.877381 | 0.01070603 |
| CDKL1 | 8814 | cyclin dependent kinase like 1 | 0.87661238 | 0.03024505 |
| LINC00909 | 400657 | long intergenic non-protein codir | 0.87493049 | 0.01569705 |
| RPUSD4 | 84881 | RNA pseudouridylate synthase d | 0.87467156 | 0.01194033 |
| LRRIQ1 | 84125 | leucine rich repeats and IQ motif | 0.86770711 | 0.04077882 |
| FAM131B | 9715 | family with sequence similarity | 0.86750214 | 0.01176105 |
| HOMER2 | 9455 | homer scaffold protein 2 | 0.86154221 | 0.00285119 |
| NUDT3 | 11165 | nudix hydrolase 3 | 0.8569471 | 0.00092452 |
| FASTKD2 | 22868 | FAST kinase domains 2 | 0.85658785 | 0.00475156 |
| JADE2 | 23338 | jade family PHD finger 2 | 0.85650601 | 0.01381932 |
| RORA | 6095 | RAR related orphan receptor A | 0.85280233 | 0.01477118 |
| ERP27 | 121506 | endoplasmic reticulum protein 2 | 0.84855923 | 0.00753522 |
| NFE2L3 | 9603 | nuclear factor, erythroid 2 like 3 | 0.84797808 | 0.02686928 |
| MT1E | 4493 | metallothionein 1E | 0.84608667 | 0.02210353 |
| HSPA13 | 6782 | heat shock protein family A (Hsp | 0.84471469 | 0.0020531 |
| ROBO1 | 6091 | roundabout guidance receptor 1 | 0.84461946 | 0.00738307 |
| BUD31 | 8896 | BUD31 homolog | 0.84424345 | 0.00578988 |
| SULF2 | 55959 | sulfatase 2 | 0.84180975 | 0.00731415 |
| JDP2 | 122953 | Jun dimerization protein 2 | 0.84034159 | 0.00181597 |
| NEDD4 | 4734 | neural precursor cell expressed, | 0.8402335 | 0.03777574 |
| MARS | 4141 | methionyl-tRNA synthetase | 0.84017281 | 0.00164205 |
| PYROXD1 | 79912 | pyridine nucleotide-disulphide o | 0.84001517 | 0.02353273 |
| NRP2 | 8828 | neuropilin 2 | 0.83969116 | 0.02812302 |
| PTPRG | 5793 | protein tyrosine phosphatase, re | 0.83853776 | 0.01993575 |
| MT1G | 4495 | metallothionein 1G | 0.83774771 | 0.0357167 |
| DRAM1 | 55332 | DNA damage regulated autopha | 0.83769827 | 0.00915219 |
| APOL6 | 80830 | apolipoprotein L6 | 0.83671487 | 0.00578861 |
| MPP6 | 51678 | membrane palmitoylated protei | 0.83523276 | 0.01477118 |
| NFIC | 4782 | nuclear factor I C | 0.83191668 | 0.00157199 |
| CASC1 | 55259 | cancer susceptibility 1 | 0.83095741 | 0.03739957 |
| FKTN | 2218 | fukutin | 0.82932817 | 0.02210353 |
| ALDH1B1 | 219 | aldehyde dehydrogenase 1 famil | 0.82930015 | 0.03873331 |
| MYO10 | 4651 | myosin X | 0.82898282 | 0.00684957 |
| ERN1 | 2081 | endoplasmic reticulum to nucleu | 0.82700792 | 0.01002957 |
| HKDC1 | 80201 | hexokinase domain containing 1 | 0.82518843 | 0.00274473 |
| PYCR1 | 5831 | pyrroline-5-carboxylate reducta | 0.8245891 | 0.00353101 |
| ZNF618 | 114991 | zinc finger protein 618 | 0.82425081 | 0.01250152 |
| CIB2 | 10518 | calcium and integrin binding fam | 0.82244189 | 0.00850555 |
| LONP1 | 9361 | lon peptidase 1, mitochondrial | 0.82015022 | 0.01077355 |
| RASGRP1 | 10125 | RAS guanyl releasing protein 1 | 0.818889 | 0.01175754 |
| RPS6KA2 | 6196 | ribosomal protein S6 kinase A2 | 0.81862044 | 0.01787183 |
| DYNC1H1 | 1778 | dynein cytoplasmic 1 heavy chai | 0.81730375 | 0.01317843 |
| CRYL1 | 51084 | crystallin lambda 1 | 0.81712075 | 0.02286197 |
| PCCA | 5095 | propionyl-CoA carboxylase alpha | 0.81685304 | 0.00766691 |
| METTL8 | 79828 | methyltransferase like 8 | 0.81446819 | 0.00645954 |
| TGM2 | 7052 | transglutaminase 2 | 0.81347962 | 0.02437293 |
| CCPG1 | 9236 | cell cycle progression 1 | 0.81318996 | 0.01395726 |

| | | | | |
|---|---|---|---|---|
| MYO1B | 4430 | myosin IB | 0.94502292 | 0.00553008 |
| CACNA2D3 | 55799 | calcium voltage-gated channel auxiliary sub | 0.94350044 | 0.04194009 |
| VWA2 | 340706 | von Willebrand factor A domain containing | 0.94304236 | 0.00649818 |
| FHL3 | 2275 | four and a half LIM domains 3 | 0.94002158 | 0.02705905 |
| SERPINB7 | 8710 | serpin family B member 7 | 0.93858672 | 0.01138615 |
| ZMAT3 | 64393 | zinc finger matrin-type 3 | 0.93851217 | 0.00127925 |
| CXCL1 | 2919 | C-X-C motif chemokine ligand 1 | 0.93478563 | 0.00185005 |
| BMP4 | 652 | bone morphogenetic protein 4 | 0.93295304 | 0.01137402 |
| ARSE | 415 | arylsulfatase E (chondrodysplasia punctata | 0.93287318 | 0.02818064 |
| ROBO1 | 6091 | roundabout guidance receptor 1 | 0.92379346 | 0.00256603 |
| MDM2 | 4193 | MDM2 proto-oncogene | 0.92257009 | 0.00068044 |
| HSPA12A | 259217 | heat shock protein family A (Hsp70) membe | 0.92219955 | 0.00521077 |
| TBC1D19 | 55296 | TBC1 domain family member 19 | 0.92035134 | 0.0027805 |
| TP53 | 7157 | tumor protein p53 | 0.91758067 | 0.00063291 |
| ZC3H12C | 85463 | zinc finger CCCH-type containing 12C | 0.91594485 | 0.00112841 |
| DCP1B | 196513 | decapping mRNA 1B | 0.91516526 | 0.00215339 |
| CIITA | 4261 | class II major histocompatibility complex tr | 0.91056655 | 0.00059134 |
| LRIG1 | 26018 | leucine rich repeats and immunoglobulin lik | 0.90888011 | 0.03705029 |
| CD74 | 972 | CD74 molecule | 0.90870605 | 0.00386342 |
| SLC3A2 | 6520 | solute carrier family 3 member 2 | 0.90797936 | 0.00119272 |
| SEMA3C | 10512 | semaphorin 3C | 0.90588728 | 0.00944718 |
| SLC7A1 | 6541 | solute carrier family 7 member 1 | 0.90548153 | 0.00251212 |
| LARS | 51520 | leucyl-tRNA synthetase | 0.90403074 | 0.00050772 |
| HIF1A | 3091 | hypoxia inducible factor 1 alpha subunit | 0.90359028 | 0.00190252 |
| SHMT2 | 6472 | serine hydroxymethyltransferase 2 | 0.90354266 | 0.0018911 |
| ZNF618 | 114991 | zinc finger protein 618 | 0.90023011 | 0.00487858 |
| RHOQ | 23433 | ras homolog family member Q | 0.89770085 | 0.0151941 |
| MAGED1 | 9500 | MAGE family member D1 | 0.89759588 | 0.02756235 |
| PTCD3 | 55037 | pentatricopeptide repeat domain 3 | 0.89732389 | 0.01722169 |
| KCNQ1 | 3784 | potassium voltage-gated channel subfamily | 0.89654886 | 0.0010954 |
| ERN2 | 10595 | endoplasmic reticulum to nucleus signaling | 0.89605472 | 0.00081817 |
| LTB | 4050 | lymphotoxin beta | 0.89558445 | 0.00261623 |
| CCND2 | 894 | cyclin D2 | 0.89248975 | 0.00062963 |
| UBE2Q2 | 92912 | ubiquitin conjugating enzyme E2 Q2 | 0.89003518 | 0.0114678 |
| CEP57 | 9702 | centrosomal protein 57 | 0.88707109 | 0.004053 |
| APBB2 | 323 | amyloid beta precursor protein binding fam | 0.8859306 | 0.00427446 |
| PCDHGC3 | 5098 | protocadherin gamma subfamily C, 3 | 0.87559276 | 0.00340751 |
| PIGR | 5284 | polymeric immunoglobulin receptor | 0.87207396 | 0.00039468 |
| RGMB-AS1 | 503569 | RGMB antisense RNA 1 | 0.87178199 | 0.04727269 |
| CARD16 | 114769 | caspase recruitment domain family membe | 0.86903535 | 0.00343595 |
| ZDHHC1 | 29800 | zinc finger DHHC-type containing 1 | 0.86833285 | 0.02422437 |
| ZCCHC11 | 23318 | zinc finger CCHC-type containing 11 | 0.86831036 | 0.04407094 |
| TRPS1 | 7227 | transcriptional repressor GATA binding 1 | 0.86565574 | 0.03299749 |
| DEPTOR | 64798 | DEP domain containing MTOR interacting p | 0.86327723 | 0.00213837 |
| JMY | 133746 | junction mediating and regulatory protein, | 0.86249885 | 0.02059006 |
| SNORD41 | 26810 | small nucleolar RNA, C/D box 41 | 0.86006134 | 0.02853881 |
| BCKDHA | 593 | branched chain keto acid dehydrogenase E1 | 0.85924175 | 0.00277438 |
| CARS | 833 | cysteinyl-tRNA synthetase | 0.85757698 | 0.00234724 |
| PTPRG | 5793 | protein tyrosine phosphatase, receptor type | 0.85749623 | 0.01319979 |
| SNTB1 | 6641 | syntrophin beta 1 | 0.8563837 | 0.00900021 |
| H6PD | 9563 | hexose-6-phosphate dehydrogenase/glucos | 0.85596179 | 0.00347366 |
| RNF144B | 255488 | ring finger protein 144B | 0.85567239 | 0.00328806 |
| NUP133 | 55746 | nucleoporin 133 | 0.85263386 | 0.01093972 |
| ANGPT2 | 285 | angiopoietin 2 | 0.85078613 | 0.02205739 |
| CYFIP2 | 26999 | cytoplasmic FMR1 interacting protein 2 | 0.85058448 | 0.00168906 |
| METAP1D | 254042 | methionyl aminopeptidase type 1D, mitoch | 0.84872364 | 0.00997376 |
| HSCB | 150274 | HscB mitochondrial iron-sulfur cluster coch | 0.84718126 | 0.00552578 |
| TSHZ2 | 128553 | teashirt zinc finger homeobox 2 | 0.84365966 | 0.02398959 |
| ECM1 | 1893 | extracellular matrix protein 1 | 0.8423639 | 0.02658288 |
| ZNF423 | 23090 | zinc finger protein 423 | 0.83874501 | 0.00346431 |
| LINC00266-1 | 140849 | long intergenic non-protein coding RNA 266 | 0.83723495 | 0.04985729 |
| RRH | 10692 | retinal pigment epithelium-derived rhodops | 0.83672186 | 0.0398527 |
| FAIM | 55179 | Fas apoptotic inhibitory molecule | 0.83513768 | 0.00361143 |
| HOXA5 | 3202 | homeobox A5 | 0.83280777 | 0.02437858 |
| SNORD22 | 9304 | small nucleolar RNA, C/D box 22 | 0.83263402 | 0.04727269 |
| CLIC4 | 25932 | chloride intracellular channel 4 | 0.83139442 | 0.0037364 |
| MTRNR2L3 | 100462983 | MT-RNR2 like 3 | 0.83082334 | 0.00400308 |
| ANXA9 | 8416 | annexin A9 | 0.83059983 | 0.01824248 |
| EPB41L2 | 2037 | erythrocyte membrane protein band 4.1 like | 0.8294234 | 0.00550342 |
| KCNK15 | 60598 | potassium two pore domain channel subfar | 0.82798315 | 0.0290284 |
| DDIT3 | 1649 | DNA damage inducible transcript 3 | 0.8279422 | 0.02344581 |
| ATF4 | 468 | activating transcription factor 4 | 0.82749742 | 0.00070283 |
| ALKBH8 | 91801 | alkB homolog 8, tRNA methyltransferase | 0.82678191 | 0.02691539 |
| CELF2 | 10659 | CUGBP Elav-like family member 2 | 0.82672704 | 0.00497444 |

| | | | | |
|---|---|---|---|---|
| SLC5A1 | 6523 | solute carrier family 5 member 1 | -1.0929192 | 0.02572743 |
| DEPDC4 | 120863 | DEP domain containing 4 | -1.0957499 | 0.04789339 |
| BCL2L14 | 79370 | BCL2 like 14 | -1.0958644 | 4.86E-05 |
| ASAH2B | 653308 | N-acylsphingosine amidohydrolase 2B | -1.1022943 | 0.01417659 |
| XRRA1 | 143570 | X-ray radiation resistance associated 1 | -1.1067407 | 0.0034493 |
| CHAD | 1101 | chondroadherin | -1.1109076 | 0.00430211 |
| CFTR | 1080 | cystic fibrosis transmembrane conduct | -1.1110544 | 0.01973317 |
| MYO7B | 4648 | myosin VIIB | -1.1112777 | 0.0130349 |
| PDE3B | 5140 | phosphodiesterase 3B | -1.1145466 | 0.0143189 |
| SEMG1 | 6406 | semenogelin 1 | -1.1162015 | 0.00298835 |
| HLA-DPA1 | 3113 | major histocompatibility complex, clas | -1.123075 | 0.01214527 |
| SOX6 | 55553 | SRY-box 6 | -1.1242668 | 0.04686783 |
| MAN1A1 | 4121 | mannosidase alpha class 1A member 1 | -1.1275669 | 0.00120433 |
| MS4A8 | 83661 | membrane spanning 4-domains A8 | -1.1312032 | 0.01120699 |
| MT1B | 4490 | metallothionein 1B | -1.1458125 | 0.0270896 |
| FAM3B | 54097 | family with sequence similarity 3 mem | -1.1711542 | 0.0029489 |
| ITLN1 | 55600 | intelectin 1 | -1.1869924 | 0.00627113 |
| MST1L | 11223 | macrophage stimulating 1 like | -1.1967782 | 0.00047264 |
| MTTP | 4547 | microsomal triglyceride transfer protei | -1.2022836 | 0.0357315 |
| QKI | 9444 | QKI, KH domain containing RNA binding | -1.2102905 | 0.00014807 |
| PLA2G7 | 7941 | phospholipase A2 group VII | -1.2112852 | 0.035878 |
| PPARGC1A | 10891 | PPARG coactivator 1 alpha | -1.2266074 | 0.01966968 |
| HABP2 | 3026 | hyaluronan binding protein 2 | -1.2551966 | 0.02359651 |
| MT2A | 4502 | metallothionein 2A | -1.2791889 | 0.01507643 |
| GC | 2638 | GC, vitamin D binding protein | -1.3066747 | 0.00632075 |
| MIR4653 | 100616117 | microRNA 4653 | -1.3345083 | 0.00088552 |
| MOCOS | 55034 | molybdenum cofactor sulfurase | -1.370338 | 0.00035063 |
| ASAH2 | 56624 | N-acylsphingosine amidohydrolase 2 | -1.3786735 | 0.01704716 |
| MITF | 4286 | melanogenesis associated transcriptio | -1.3913143 | 0.00047264 |
| GABRE | 2564 | gamma-aminobutyric acid type A recep | -1.4121293 | 0.01876271 |
| SLC19A3 | 80704 | solute carrier family 19 member 3 | -1.4214145 | 0.01876271 |
| CFI | 3426 | complement factor I | -1.4257336 | 0.01027145 |
| APOB | 338 | apolipoprotein B | -1.4282766 | 0.04164816 |
| DPP4 | 1803 | dipeptidyl peptidase 4 | -1.4732159 | 0.01703235 |
| TLR6 | 10333 | toll like receptor 6 | -1.4917205 | 0.00384047 |
| MIR221 | 407006 | microRNA 221 | -1.4993994 | 0.04688034 |
| ADGRL2 | 23266 | adhesion G protein-coupled receptor L2 | -1.5176201 | 0.00147349 |
| BBOF1 | 80127 | basal body orientation factor 1 | -1.5298209 | 0.0002455 |
| ANXA13 | 312 | annexin A13 | -1.5372503 | 0.00067867 |
| ENTPD5 | 957 | ectonucleoside triphosphate diphospho | -1.5392904 | 0.00078564 |
| ZC3H12C | 85463 | zinc finger CCCH-type containing 12C | -1.5476016 | 5.31E-05 |
| DSEL | 92126 | dermatan sulfate epimerase like | -1.5854592 | 0.00648883 |
| F11 | 2160 | coagulation factor XI | -1.6292778 | 0.00384047 |
| RGPD1 | 400966 | RANBP2-like and GRIP domain contain | -1.6930974 | 0.00319437 |
| TINAG | 27283 | tubulointerstitial nephritis antigen | -1.7016638 | 0.02530861 |
| DCLK2 | 166614 | doublecortin like kinase 2 | -1.7816778 | 0.02231447 |
| CRACR2A | 84766 | calcium release activated channel regu | -1.8060397 | 7.42E-05 |
| IFI16 | 3428 | interferon gamma inducible protein 16 | -1.8304287 | 0.00026554 |
| DAB2 | 1601 | DAB2, clathrin adaptor protein | -1.8530388 | 0.00035298 |
| MMP7 | 4316 | matrix metallopeptidase 7 | -1.9383093 | 0.00636817 |
| MCOLN3 | 55283 | mucolipin 3 | -1.974876 | 0.02677005 |
| MSMB | 4477 | microseminoprotein beta | -2.049306 | 0.00095814 |
| EPB41L3 | 23136 | erythrocyte membrane protein band 4. | -2.0985123 | 0.00744709 |
| TMEM220 | 388335 | transmembrane protein 220 | -2.10351 | 0.00022077 |
| GSTM3 | 2947 | glutathione S-transferase mu 3 | -2.130355 | 8.13E-05 |
| KCNJ13 | 3769 | potassium voltage-gated channel subfa | -2.1787941 | 0.00204872 |
| TCN1 | 6947 | transcobalamin 1 | -2.1960233 | 0.00088552 |
| BST1 | 683 | bone marrow stromal cell antigen 1 | -2.2918998 | 0.00067832 |
| CACNA2D1 | 781 | calcium voltage-gated channel auxiliar | -2.307905 | 7.42E-05 |
| MATN2 | 4147 | matrilin 2 | -2.4853764 | 3.97E-06 |
| SULT2A1 | 6822 | sulfotransferase family 2A member 1 | -2.5785729 | 0.00319437 |
| CXCL5 | 6374 | C-X-C motif chemokine ligand 5 | -2.586426 | 3.65E-05 |
| CLCA1 | 1179 | chloride channel accessory 1 | -2.6038289 | 0.0039952 |
| CLDN10 | 9071 | claudin 10 | -3.0859619 | 0.00022215 |
| RGPD2 | 729857 | RANBP2-like and GRIP domain contain | -3.33129 | 2.87E-06 |
| VNN1 | 8876 | vanin 1 | -3.6155352 | 7.42E-05 |
| MME | 4311 | membrane metalloendopeptidase | -3.9127912 | 7.42E-05 |
| CYP4F11 | 57834 | cytochrome P450 family 4 subfamily F | -5.282361 | 9.40E-08 |

| | | | | |
|---|---|---|---|---|
| GRPEL2 | 134266 | GrpE like 2, mitochondrial | 0.81271828 | 0.00352094 |
| TUBE1 | 51175 | tubulin epsilon 1 | 0.8126797 | 0.02896774 |
| ZHX2 | 22882 | zinc fingers and homeoboxes 2 | 0.81234536 | 0.01360211 |
| ZBTB37 | 84614 | zinc finger and BTB domain cont | 0.8121692 | 0.01045399 |
| TIMM44 | 10469 | translocase of inner mitochondri | 0.81106584 | 0.00721138 |
| AGMAT | 79814 | agmatinase | 0.80956522 | 0.00318161 |
| ZFP69B | 65243 | ZFP69 zinc finger protein B | 0.80832831 | 0.01209222 |
| TRHDE | 29953 | thyrotropin releasing hormone de | 0.8067556 | 0.04446091 |
| SLFN5 | 162394 | schlafen family member 5 | 0.8056256 | 0.01375796 |
| OR14C36 | 127066 | olfactory receptor family 14 subf | 0.80537954 | 0.03640358 |
| PIM1 | 5292 | Pim-1 proto-oncogene, serine/th | 0.80466069 | 0.01152192 |
| PADI2 | 11240 | peptidyl arginine deiminase 2 | 0.80409713 | 0.03739957 |
| LOXL1 | 4016 | lysyl oxidase like 1 | 0.80294588 | 0.03650166 |
| RAB15 | 376267 | RAB15, member RAS oncogene f | 0.80290768 | 0.00334678 |
| TCP11L2 | 255394 | t-complex 11 like 2 | 0.80130852 | 0.02438117 |
| ANKRD6 | 22881 | ankyrin repeat domain 6 | 0.80023693 | 0.01855795 |
| RAP1GAP2 | 23108 | RAP1 GTPase activating protein 2 | 0.79931757 | 0.00186408 |
| ZNF425 | 155054 | zinc finger protein 425 | 0.79803189 | 0.03798587 |
| TTC39B | 158219 | tetratricopeptide repeat domain | 0.79703568 | 0.00961469 |
| IGFBP1 | 3484 | insulin like growth factor binding | 0.79267923 | 0.01330834 |
| SLC7A11-AS1 | 641364 | SLC7A11 antisense RNA 1 | 0.79223946 | 0.02114272 |
| CDX1 | 1044 | caudal type homeobox 1 | 0.79187896 | 0.00502693 |
| PPIL3 | 53938 | peptidylprolyl isomerase like 3 | 0.79186407 | 0.00114511 |
| ANKRD29 | 147463 | ankyrin repeat domain 29 | 0.79080629 | 0.01868408 |
| YARS2 | 51067 | tyrosyl-tRNA synthetase 2 | 0.7907692 | 0.01225833 |
| ZNF566 | 84924 | zinc finger protein 566 | 0.79021742 | 0.03257954 |
| PTPRM | 5797 | protein tyrosine phosphatase, re | 0.79005642 | 0.03112259 |
| NDUFA4L2 | 56901 | NDUFA4, mitochondrial complex | 0.78835711 | 0.0053669 |
| CDYL2 | 124359 | chromodomain Y like 2 | 0.78760186 | 0.00508418 |
| ADGRG1 | 9289 | adhesion G protein-coupled rece | 0.78571578 | 0.00857319 |
| ASIC1 | 41 | acid sensing ion channel subunit | 0.78348432 | 0.02439953 |
| VWA8 | 23078 | von Willebrand factor A domain | 0.7833225 | 0.00213881 |
| CBX4 | 8535 | chromobox 4 | 0.78323454 | 0.00824841 |
| DNASE2 | 1777 | deoxyribonuclease 2, lysosomal | 0.78096 | 0.00347115 |
| EXOSC5 | 56915 | exosome component 5 | 0.78079378 | 0.01920852 |
| CKAP4 | 10970 | cytoskeleton associated protein 4 | 0.78041049 | 0.01045399 |
| DHX33 | 56919 | DEAH-box helicase 33 | 0.77971694 | 0.01077355 |
| LONRF3 | 79836 | LON peptidase N-terminal doma | 0.77901134 | 0.02335221 |
| RNF217 | 154214 | ring finger protein 217 | 0.77854237 | 0.02078528 |
| SRD5A1 | 6715 | steroid 5 alpha-reductase 1 | 0.77830051 | 0.00824841 |
| MIR548Y | 100500919 | microRNA 548y | 0.77818609 | 0.02014679 |
| HBEGF | 1839 | heparin binding EGF like growth | 0.77678622 | 0.01424078 |
| CSF1 | 1435 | colony stimulating factor 1 | 0.7763415 | 0.00238927 |
| DNAJA3 | 9093 | DnaJ heat shock protein family (H | 0.77488814 | 0.00177634 |
| PRKCB | 5579 | protein kinase C beta | 0.77472743 | 0.02144448 |
| INSL6 | 11172 | insulin like 6 | 0.77337091 | 0.02930689 |
| TMEM182 | 130827 | transmembrane protein 182 | 0.7721357 | 0.00611026 |
| PTGES3L-AAR | 100885850 | PTGES3L-AARSD1 readthrough | 0.77170107 | 0.03044696 |
| PLAU | 5328 | plasminogen activator, urokinase | 0.77096993 | 0.00553995 |
| TFB2M | 64216 | transcription factor B2, mitochor | 0.76929809 | 0.00353101 |
| AKAP7 | 9465 | A-kinase anchoring protein 7 | 0.76910644 | 0.0135338 |
| TMEM156 | 80008 | transmembrane protein 156 | 0.76821054 | 0.02439953 |
| KBTBD11 | 9920 | kelch repeat and BTB domain co | 0.76601296 | 0.02759414 |
| PRMT3 | 10196 | protein arginine methyltransfera | 0.76542867 | 0.04004042 |
| SOAT1 | 6646 | sterol O-acyltransferase 1 | 0.76525144 | 0.02014679 |
| PSPH | 5723 | phosphoserine phosphatase | 0.76505398 | 0.01318555 |
| ZMAT3 | 64393 | zinc finger matrin-type 3 | 0.76490402 | 0.00768409 |
| QRSL1 | 55278 | glutaminyl-tRNA synthase (gluta | 0.76305448 | 0.00496767 |
| GGACT | 87769 | gamma-glutamylamine cyclotran | 0.76298395 | 0.01288736 |
| ABCA13 | 154664 | ATP binding cassette subfamily A | 0.76218764 | 0.02106641 |
| E2F5 | 1875 | E2F transcription factor 5 | 0.76217446 | 0.00620708 |
| TCEA3 | 6920 | transcription elongation factor A | 0.76186881 | 0.00738307 |
| PLEC | 5339 | plectin | 0.76099166 | 0.03305594 |
| PADI4 | 23569 | peptidyl arginine deiminase 4 | 0.76044027 | 0.03992953 |
| FAM129A | 116496 | family with sequence similarity 1 | 0.76042396 | 0.01535683 |
| APLF | 200558 | aprataxin and PNKP like factor | 0.7583103 | 0.00578988 |
| DCDC2 | 51473 | doublecortin domain containing 2 | 0.75745892 | 0.03669796 |
| PRKDC | 5591 | protein kinase, DNA-activated, ca | 0.75715098 | 0.01253396 |
| GAB2 | 9846 | GRB2 associated binding protein | 0.75653781 | 0.03202817 |
| KLLN | 100144748 | killin, p53 regulated DNA replicat | 0.75451926 | 0.02211568 |
| MR1 | 3140 | major histocompatibility comple | 0.75417722 | 0.00526073 |
| LRRC31 | 79782 | leucine rich repeat containing 31 | 0.75411557 | 0.0095712 |
| CRNDE | 643911 | colorectal neoplasia differentiall | 0.75343648 | 0.03044696 |
| HERPUD1 | 9709 | homocysteine inducible ER prote | 0.75280524 | 0.00213615 |

| | | | | |
|---|---|---|---|---|
| DLGAP1-AS2 | 84777 | DLGAP1 antisense RNA 2 | 0.82551896 | 0.00949621 |
| ENAH | 55740 | ENAH, actin regulator | 0.82461093 | 0.00336054 |
| VSTM5 | 387804 | V-set and transmembrane domain containi | 0.82159367 | 0.00251694 |
| ZNF391 | 346157 | zinc finger protein 391 | 0.82153339 | 0.04424537 |
| NPL | 80896 | N-acetylneuraminate pyruvate lyase | 0.82108546 | 0.03393019 |
| EIF3L | 51386 | eukaryotic translation initiation factor 3 sub | 0.81981581 | 0.00404372 |
| ZHX2 | 22882 | zinc fingers and homeoboxes 2 | 0.81884945 | 0.00906551 |
| SLC12A2 | 6558 | solute carrier family 12 member 2 | 0.81590439 | 0.01877074 |
| WAC-AS1 | 220906 | WAC antisense RNA 1 (head to head) | 0.81287441 | 0.01062257 |
| ZNF827 | 152485 | zinc finger protein 827 | 0.81258219 | 0.0048841 |
| LRRN4 | 164312 | leucine rich repeat neuronal 4 | 0.81226521 | 0.00108344 |
| NFIX | 4784 | nuclear factor I X | 0.81097981 | 0.00188893 |
| TTC39C | 125488 | tetratricopeptide repeat domain 39C | 0.81076337 | 0.03133 |
| KCNN4 | 3783 | potassium calcium-activated channel subfa | 0.80712382 | 0.0056327 |
| MCF2L | 23263 | MCF.2 cell line derived transforming sequer | 0.80668571 | 0.00316067 |
| AARS | 16 | alanyl-tRNA synthetase | 0.8062457 | 0.00248926 |
| PVT1 | 5820 | Pvt1 oncogene (non-protein coding) | 0.80581031 | 0.03375477 |
| FLRT3 | 23767 | fibronectin leucine rich transmembrane pro | 0.80253073 | 0.00831486 |
| PITPNC1 | 26207 | phosphatidylinositol transfer protein cytopla | 0.79895033 | 0.01611112 |
| CALB2 | 794 | calbindin 2 | 0.79890561 | 0.03098601 |
| GSAP | 54103 | gamma-secretase activating protein | 0.79767512 | 0.00505435 |
| TNFSF12-TNI | 407977 | TNFSF12-TNFSF13 readthrough | 0.79740021 | 0.00112027 |
| UBE2E2 | 7325 | ubiquitin conjugating enzyme E2 E2 | 0.7973506 | 0.00386342 |
| NOB1 | 28987 | NIN1 (RPN12) binding protein 1 homolog | 0.79721166 | 0.00165149 |
| REV3L | 5980 | REV3 like, DNA directed polymerase zeta ca | 0.7971538 | 0.00481385 |
| GPC4 | 2239 | glypican 4 | 0.79652508 | 0.03908079 |
| PSPH | 5723 | phosphoserine phosphatase | 0.79619609 | 0.00707865 |
| CDK6 | 1021 | cyclin dependent kinase 6 | 0.79610633 | 0.004053 |
| ULBP3 | 79465 | UL16 binding protein 3 | 0.79460697 | 0.01945689 |
| ZC3H12A | 80149 | zinc finger CCCH-type containing 12A | 0.79454613 | 0.00227777 |
| ASAP1 | 50807 | ArfGAP with SH3 domain, ankyrin repeat an | 0.79434397 | 0.01729513 |
| MCCC2 | 64087 | methylcrotonoyl-CoA carboxylase 2 | 0.79395132 | 0.00404372 |
| TNFSF13 | 8741 | TNF superfamily member 13 | 0.79367824 | 0.00127925 |
| DNMT3B | 1789 | DNA methyltransferase 3 beta | 0.78916903 | 0.01644933 |
| PARP15 | 165631 | poly(ADP-ribose) polymerase family membe | 0.78877236 | 0.02506923 |
| ZNF25 | 219749 | zinc finger protein 25 | 0.78697216 | 0.01431609 |
| PRPF40B | 25766 | pre-mRNA processing factor 40 homolog B | 0.78484738 | 0.01139366 |
| COLCA2 | 120376 | colorectal cancer associated 2 | 0.78194549 | 0.00373215 |
| PTPRM | 5797 | protein tyrosine phosphatase, receptor type | 0.78077388 | 0.02604542 |
| ASS1 | 445 | argininosuccinate synthase 1 | 0.78018027 | 0.00135158 |
| GCKR | 2646 | glucokinase regulator | 0.77957488 | 0.01767177 |
| SPATA18 | 132671 | spermatogenesis associated 18 | 0.77826085 | 0.00080724 |
| APOBEC3C | 27350 | apolipoprotein B mRNA editing enzyme cat | 0.7779163 | 0.00380658 |
| EFNA5 | 1946 | ephrin A5 | 0.77711045 | 0.00655449 |
| KIF26B | 55083 | kinesin family member 26B | 0.77594077 | 0.01917458 |
| DRD1 | 1812 | dopamine receptor D1 | 0.77466625 | 0.02783437 |
| ZNF805 | 390980 | zinc finger protein 805 | 0.77453125 | 0.00633356 |
| INHBB | 3625 | inhibin beta B subunit | 0.77446918 | 0.01431609 |
| RNF130 | 55819 | ring finger protein 130 | 0.77377151 | 0.00383012 |
| KIAA1549 | 57670 | KIAA1549 | 0.77326741 | 0.01824248 |
| BANK1 | 55024 | B cell scaffold protein with ankyrin repeats | 0.77267244 | 0.00583327 |
| TP53BP1 | 7158 | tumor protein p53 binding protein 1 | 0.77234996 | 0.04060562 |
| DLG4 | 1742 | discs large MAGUK scaffold protein 4 | 0.77195678 | 0.00490627 |
| DAGLA | 747 | diacylglycerol lipase alpha | 0.77143971 | 0.00143325 |
| IFRD1 | 3475 | interferon related developmental regulator | 0.7711819 | 0.01321794 |
| VANGL2 | 57216 | VANGL planar cell polarity protein 2 | 0.76683504 | 0.00648967 |
| LRRC31 | 79782 | leucine rich repeat containing 31 | 0.76520326 | 0.00552578 |
| DNAL1 | 83544 | dynein axonemal light chain 1 | 0.7649416 | 0.01242009 |
| ROCK2 | 9475 | Rho associated coiled-coil containing protei | 0.76416805 | 0.00276248 |
| TSC22D3 | 1831 | TSC22 domain family member 3 | 0.75998984 | 0.02380517 |
| ZBTB47 | 92999 | zinc finger and BTB domain containing 47 | 0.75985627 | 0.01793152 |
| HUNK | 30811 | hormonally up-regulated Neu-associated ki | 0.75952771 | 0.03137534 |
| CDRT1 | 374286 | CMT1A duplicated region transcript 1 | 0.75821594 | 0.02621956 |
| TMPRSS3 | 64699 | transmembrane serine protease 3 | 0.75488864 | 0.01129331 |
| SNORD1A | 677848 | small nucleolar RNA, C/D box 1A | 0.75192958 | 0.01424128 |
| PDCD11 | 22984 | programmed cell death 11 | 0.75120725 | 0.0109246 |
| PRNCR1 | 101867536 | prostate cancer associated non-coding RNA | 0.75111672 | 0.04562339 |
| SPX | 80763 | spexin hormone | 0.75044706 | 0.03613612 |
| POC5 | 134359 | POC5 centriolar protein | 0.74920607 | 0.02966812 |
| FXYD5 | 53827 | FXYD domain containing ion transport regul | 0.74839329 | 0.02730782 |
| TMEM173 | 340061 | transmembrane protein 173 | 0.7478014 | 0.02264169 |
| STEAP1 | 26872 | STEAP family member 1 | 0.74759508 | 0.01138615 |
| LRRC37A | 9884 | leucine rich repeat containing 37A | 0.74725955 | 0.02147687 |
| MRAS | 22808 | muscle RAS oncogene homolog | 0.7472017 | 0.0158275 |

| | | | | |
|---|---|---|---|---|
| ABCA1 | 19 | ATP binding cassette subfamily A | 0.75268937 | 0.01818588 |
| DEPTOR | 64798 | DEP domain containing MTOR in | 0.75176622 | 0.00796029 |
| AKR1C2 | 1646 | aldo-keto reductase family 1 me | 0.75162289 | 0.04446091 |
| SLC25A43 | 203427 | solute carrier family 25 member | 0.75130352 | 0.01249066 |
| RHOBTB3 | 22836 | Rho related BTB domain contain | 0.7508895 | 0.03294735 |
| CD22 | 933 | CD22 molecule | 0.7505902 | 0.0385069 |
| UBE2E2 | 7325 | ubiquitin conjugating enzyme E2 | 0.74910098 | 0.0089459 |
| DBNDD2 | 55861 | dysbindin domain containing 2 | 0.7490577 | 0.00701598 |
| PRDM1 | 639 | PR/SET domain 1 | 0.74841263 | 0.01915875 |
| KIZ | 55857 | kizuna centrosomal protein | 0.74833644 | 0.03330983 |
| MT1X | 4501 | metallothionein 1X | 0.74680743 | 0.00836144 |
| PRKCH | 5583 | protein kinase C eta | 0.746354 | 0.02014679 |
| TRIAP1 | 51499 | TP53 regulated inhibitor of apopt | 0.74622205 | 0.01262385 |
| GYG2 | 8908 | glycogenin 2 | 0.74598494 | 0.00836144 |
| TRAPPC2L | 51693 | trafficking protein particle compl | 0.74559967 | 0.02434502 |
| TP53 | 7157 | tumor protein p53 | 0.74204019 | 0.00463362 |
| RNF180 | 285671 | ring finger protein 180 | 0.74079045 | 0.00553995 |
| HLCS | 3141 | holocarboxylase synthetase | 0.73960971 | 0.00836144 |
| PIK3R1 | 5295 | phosphoinositide-3-kinase regula | 0.73890233 | 0.00526073 |
| SLC25A10 | 1468 | solute carrier family 25 member | 0.73715199 | 0.00445433 |
| PMAIP1 | 5366 | phorbol-12-myristate-13-acetate | 0.73690376 | 0.0317179 |
| MRPL40 | 64976 | mitochondrial ribosomal protein | 0.73456316 | 0.01189168 |
| TNFRSF10D | 8793 | TNF receptor superfamily memb | 0.7330767 | 0.01176105 |
| FAM84B | 157638 | family with sequence similarity 8 | 0.72968687 | 0.01194033 |
| CCND2-AS1 | 103752584 | CCND2 antisense RNA 1 | 0.7295872 | 0.01428227 |
| HSPA9 | 3313 | heat shock protein family A (Hsp | 0.72933383 | 0.00238505 |
| CHCHD4 | 131474 | coiled-coil-helix-coiled-coil-helix | 0.72918587 | 0.00843853 |
| CEBPG | 1054 | CCAAT enhancer binding protein | 0.72751652 | 0.01176105 |
| UTP20 | 27340 | UTP20, small subunit processom | 0.72618243 | 0.04785988 |
| R3HDM2 | 22864 | R3H domain containing 2 | 0.72465781 | 0.00774445 |
| ERAP2 | 64167 | endoplasmic reticulum aminope | 0.72315998 | 0.01146655 |
| TANC2 | 26115 | tetratricopeptide repeat, ankyrin | 0.72215987 | 0.01993575 |
| ZZZ3 | 26009 | zinc finger ZZ-type containing 3 | 0.7207009 | 0.01330834 |
| NOX1 | 27035 | NADPH oxidase 1 | 0.72011942 | 0.0135338 |
| SENP8 | 123228 | SUMO peptidase family member | 0.71935227 | 0.03478557 |
| CCDC51 | 79714 | coiled-coil domain containing 51 | 0.71791104 | 0.00429197 |
| SUPV3L1 | 6832 | Suv3 like RNA helicase | 0.71731028 | 0.00285119 |
| NEK3 | 4752 | NIMA related kinase 3 | 0.71696256 | 0.02227439 |
| NPRL3 | 8131 | NPR3 like, GATOR1 complex sub | 0.71543494 | 0.01194033 |
| ARHGEF10L | 55160 | Rho guanine nucleotide exchange | 0.71416707 | 0.0088443 |
| CCNB1IP1 | 57820 | cyclin B1 interacting protein 1 | 0.71366856 | 0.01818588 |
| KLHL8 | 57563 | kelch like family member 8 | 0.71264115 | 0.01278099 |
| ZC3H6 | 376940 | zinc finger CCCH-type containing | 0.71240972 | 0.01721836 |
| TNRC6C-AS1 | 100131096 | TNRC6C antisense RNA 1 | 0.7098267 | 0.03255531 |
| ZMYND12 | 84217 | zinc finger MYND-type containing | 0.70963531 | 0.01143723 |
| SNORA84 | 100124534 | small nucleolar RNA, H/ACA box | 0.70805608 | 0.00500975 |
| SLC7A6 | 9057 | solute carrier family 7 member 6 | 0.70782464 | 0.03236057 |
| TTC27 | 55622 | tetratricopeptide repeat domain | 0.70761394 | 0.0338229 |
| GFM1 | 85476 | G elongation factor mitochondria | 0.7070175 | 0.0089459 |
| SPIRE1 | 56907 | spire type actin nucleation factor | 0.70691863 | 0.02824866 |
| RASL11A | 387496 | RAS like family 11 member A | 0.70606126 | 0.02287055 |
| PIGL | 9487 | phosphatidylinositol glycan anch | 0.7042721 | 0.02851726 |
| GSTM5 | 2949 | glutathione S-transferase mu 5 | 0.70337974 | 0.00571217 |
| ZSWIM7 | 125150 | zinc finger SWIM-type containing | 0.70337411 | 0.0089459 |
| LAMC1 | 3915 | laminin subunit gamma 1 | 0.70277588 | 0.0252848 |
| THEM4 | 117145 | thioesterase superfamily membe | 0.70272465 | 0.00768409 |
| AMACR | 23600 | alpha-methylacyl-CoA racemase | 0.70246624 | 0.00474492 |
| PDCD11 | 22984 | programmed cell death 11 | 0.70199687 | 0.02096661 |
| SYS1-DBNDD | 767557 | SYS1-DBNDD2 readthrough (NMI | 0.70148562 | 0.00824841 |
| MIR3923 | 100500877 | microRNA 3923 | 0.6992825 | 0.01412733 |
| PRSS2 | 5645 | serine protease 2 | 0.69843913 | 0.00263394 |
| METTL4 | 64863 | methyltransferase like 4 | 0.69772989 | 0.00204344 |
| NFIX | 4784 | nuclear factor I X | 0.69718566 | 0.00774445 |
| GLRX2 | 51022 | glutaredoxin 2 | 0.69675998 | 0.0095712 |
| GSAP | 54103 | gamma-secretase activating pro | 0.69598806 | 0.01568881 |
| DEFB4B | 100289462 | defensin beta 4B | 0.69453523 | 0.04579475 |
| NOB1 | 28987 | NIN1 (RPN12) binding protein 1 | 0.69387005 | 0.00638247 |
| ASS1 | 445 | argininosuccinate synthase 1 | 0.69084911 | 0.0051245 |
| CREBRF | 153222 | CREB3 regulatory factor | 0.68981773 | 0.03202817 |
| EIF2S2 | 8894 | eukaryotic translation initiation f | 0.68965401 | 0.00586671 |
| ROPN1L | 83853 | rhophilin associated tail protein | 0.68958754 | 0.02661554 |
| PINK1-AS | 100861548 | PINK1 antisense RNA | 0.68886762 | 0.04843504 |
| UTP15 | 84135 | UTP15, small subunit processom | 0.68846493 | 0.02114272 |
| ANAPC1 | 64682 | anaphase promoting complex su | 0.6882883 | 0.00353101 |

| | | | | |
|---|---|---|---|---|
| POLR1A | 25885 | RNA polymerase I subunit A | 0.74513267 | 0.00634031 |
| VPS13C | 54832 | vacuolar protein sorting 13 homolog C | 0.74493241 | 0.00099227 |
| WDR73 | 84942 | WD repeat domain 73 | 0.74469151 | 0.025184 |
| GTF2IRD2B | 389524 | GTF2I repeat domain containing 2B | 0.74394316 | 0.02756235 |
| RHOBTB3 | 22836 | Rho related BTB domain containing 3 | 0.74369263 | 0.02766207 |
| DDX43 | 55510 | DEAD-box helicase 43 | 0.74290567 | 0.04157054 |
| CCDC92 | 80212 | coiled-coil domain containing 92 | 0.74213153 | 0.03700889 |
| SH2B3 | 10019 | SH2B adaptor protein 3 | 0.73995828 | 0.02283134 |
| EPB41L4A | 64097 | erythrocyte membrane protein band 4.1 like | 0.7378022 | 0.00188893 |
| PCK2 | 5106 | phosphoenolpyruvate carboxykinase 2, mito | 0.737107 | 0.0277438 |
| ORAOV1 | 220064 | oral cancer overexpressed 1 | 0.73476872 | 0.00769698 |
| MTX3 | 345778 | metaxin 3 | 0.73455029 | 0.0225104 |
| DHX9 | 1660 | DExH-box helicase 9 | 0.73244984 | 0.00099227 |
| NEIL2 | 252969 | nei like DNA glycosylase 2 | 0.73194388 | 0.01706339 |
| SNORA47 | 677828 | small nucleolar RNA, H/ACA box 47 | 0.73174804 | 0.001317 |
| CHN1 | 1123 | chimerin 1 | 0.73156551 | 0.01814983 |
| SLC43A1 | 8501 | solute carrier family 43 member 1 | 0.73152541 | 0.02229176 |
| LINC00662 | 148189 | long intergenic non-protein coding RNA 662 | 0.7313714 | 0.02584432 |
| PRKAR1B | 5575 | protein kinase cAMP-dependent type I regul | 0.73001757 | 0.00633413 |
| SYNGAP1 | 8831 | synaptic Ras GTPase activating protein 1 | 0.72915347 | 0.0427988 |
| NHS | 4810 | NHS actin remodeling regulator | 0.72856334 | 0.01188103 |
| ACKR4 | 51554 | atypical chemokine receptor 4 | 0.72793979 | 0.00507443 |
| LRIG3 | 121227 | leucine rich repeats and immunoglobulin lik | 0.72612621 | 0.00429075 |
| NPY5R | 4889 | neuropeptide Y receptor Y5 | 0.72561536 | 0.00873348 |
| KIF21B | 23046 | kinesin family member 21B | 0.72263528 | 0.00454836 |
| ZNF136 | 7695 | zinc finger protein 136 | 0.72244973 | 0.03516321 |
| USP27X | 389856 | ubiquitin specific peptidase 27, X-linked | 0.72134227 | 0.04143086 |
| CSF1 | 1435 | colony stimulating factor 1 | 0.71978804 | 0.00216681 |
| MIR1200 | 100302113 | microRNA 1200 | 0.71962563 | 0.01724591 |
| TAF1B | 9014 | TATA-box binding protein associated factor, | 0.71917794 | 0.00261623 |
| DNASE2 | 1777 | deoxyribonuclease 2, lysosomal | 0.71801571 | 0.00328806 |
| PTPN14 | 5784 | protein tyrosine phosphatase, non-receptor | 0.71761141 | 0.00228369 |
| HLA-DRB5 | 3127 | major histocompatibility complex, class II, D | 0.71491053 | 0.03822159 |
| SNORA24 | 677809 | small nucleolar RNA, H/ACA box 24 | 0.71464808 | 0.03519158 |
| JDP2 | 122953 | Jun dimerization protein 2 | 0.71410734 | 0.00272237 |
| CEP290 | 80184 | centrosomal protein 290 | 0.71221171 | 0.0139131 |
| SUPV3L1 | 6832 | Suv3 like RNA helicase | 0.71175015 | 0.00170723 |
| GRB10 | 2887 | growth factor receptor bound protein 10 | 0.71174827 | 0.03098601 |
| KLHL23 | 151230 | kelch like family member 23 | 0.71083634 | 0.03087028 |
| DIS3L2 | 129563 | DIS3 like 3'-5' exoribonuclease 2 | 0.70915053 | 0.01444033 |
| NARS | 4677 | asparaginyl-tRNA synthetase | 0.70887007 | 0.00836319 |
| NPIPA1 | 9284 | nuclear pore complex interacting protein fa | 0.70837527 | 0.03205936 |
| RAB15 | 376267 | RAB15, member RAS oncogene family | 0.70675036 | 0.00407527 |
| ADD3 | 120 | adducin 3 | 0.70668485 | 0.00265059 |
| HIP1 | 3092 | huntingtin interacting protein 1 | 0.70602584 | 0.02783287 |
| HERPUD1 | 9709 | homocysteine inducible ER protein with ubi | 0.7060119 | 0.00182614 |
| SLC7A6 | 9057 | solute carrier family 7 member 6 | 0.70376263 | 0.02658288 |
| TUBE1 | 51175 | tubulin epsilon 1 | 0.70359688 | 0.04257296 |
| PLEKHB1 | 58473 | pleckstrin homology domain containing B1 | 0.70336974 | 0.00319103 |
| RBM3 | 5935 | RNA binding motif protein 3 | 0.70297022 | 0.02191409 |
| RFX3 | 5991 | regulatory factor X3 | 0.6982342 | 0.01388357 |
| OR1N1 | 138883 | olfactory receptor family 1 subfamily N mer | 0.69778658 | 0.02705905 |
| ELK3 | 2004 | ELK3, ETS transcription factor | 0.69701402 | 0.01138615 |
| ACAD8 | 27034 | acyl-CoA dehydrogenase family member 8 | 0.69646797 | 0.01847191 |
| MTA1 | 9112 | metastasis associated 1 | 0.6957037 | 0.01943888 |
| NDUFA4L2 | 56901 | NDUFA4, mitochondrial complex associated | 0.69550174 | 0.00655449 |
| TBL1X | 6907 | transducin beta like 1 X-linked | 0.6952864 | 0.01010566 |
| EHF | 26298 | ETS homologous factor | 0.69526126 | 0.00380658 |
| ADAMTSL4 | 54507 | ADAMTS like 4 | 0.69234224 | 0.01684697 |
| LIN52 | 91750 | lin-52 DREAM MuvB core complex compone | 0.69151963 | 0.04157054 |
| ZFAND1 | 79752 | zinc finger AN1-type containing 1 | 0.69104305 | 0.00261623 |
| UST | 10090 | uronyl 2-sulfotransferase | 0.69092807 | 0.02232583 |
| CDH3 | 1001 | cadherin 3 | 0.69092249 | 0.04282902 |
| FMNL2 | 114793 | formin like 2 | 0.68993943 | 0.00429226 |
| CIRBP | 1153 | cold inducible RNA binding protein | 0.68951001 | 0.04342896 |
| DPH5 | 51611 | diphthamide biosynthesis 5 | 0.68785706 | 0.01530409 |
| LINC00909 | 400657 | long intergenic non-protein coding RNA 909 | 0.6868593 | 0.03586163 |
| TMEM254 | 80195 | transmembrane protein 254 | 0.68685818 | 0.02815188 |
| TARS | 6897 | threonyl-tRNA synthetase | 0.68622515 | 0.00946714 |
| PTCHD1 | 139411 | patched domain containing 1 | 0.68610405 | 0.02875237 |
| ANKRD6 | 22881 | ankyrin repeat domain 6 | 0.68606232 | 0.02815958 |
| ATP4B | 496 | ATPase H+/K+ transporting beta subunit | 0.68373083 | 0.03687019 |
| SLC25A36 | 55186 | solute carrier family 25 member 36 | 0.68321201 | 0.04556835 |
| PLK2 | 10769 | polo like kinase 2 | 0.68320562 | 0.0075173 |

| | | | | |
|---|---|---|---|---|
| SLC25A26 | 115286 | solute carrier family 25 member | 0.68812072 | 0.01278099 |
| MRPL39 | 54148 | mitochondrial ribosomal protein | 0.68753226 | 0.00312048 |
| BCKDHA | 593 | branched chain keto acid dehydro | 0.68738536 | 0.01493595 |
| FXN | 2395 | frataxin | 0.68734092 | 0.02167092 |
| MTIF2 | 4528 | mitochondrial translational initia | 0.68715414 | 0.01749727 |
| NPY4R | 5540 | neuropeptide Y receptor Y4 | 0.68709259 | 0.02507486 |
| RNMT | 8731 | RNA guanine-7 methyltransferas | 0.68269315 | 0.0275366 |
| MCF2L | 23263 | MCF.2 cell line derived transform | 0.68232743 | 0.01288736 |
| CTSO | 1519 | cathepsin O | 0.68221792 | 0.04665251 |
| SMIM19 | 114926 | small integral membrane protein | 0.68186146 | 0.01882443 |
| SCMH1 | 22955 | Scm polycomb group protein hon | 0.68115039 | 0.00578988 |
| MRPL23 | 6150 | mitochondrial ribosomal protein | 0.68090981 | 0.01787183 |
| ROR1 | 4919 | receptor tyrosine kinase like orph | 0.68071769 | 0.03353196 |
| RNF130 | 55819 | ring finger protein 130 | 0.68068451 | 0.01252953 |
| DBN1 | 1627 | drebrin 1 | 0.68050362 | 0.01282908 |
| PLSCR5 | 389158 | phospholipid scramblase family | 0.68019121 | 0.04731341 |
| CARD16 | 114769 | caspase recruitment domain fam | 0.67991628 | 0.01937596 |
| MUC20 | 200958 | mucin 20, cell surface associated | 0.67935006 | 0.04043865 |
| REV3L | 5980 | REV3 like, DNA directed polymer | 0.67899094 | 0.01728159 |
| CACNA1D | 776 | calcium voltage-gated channel su | 0.67875956 | 0.00624999 |
| PIK3C2G | 5288 | phosphatidylinositol-4-phosphate | 0.67743296 | 0.03590851 |
| KCNG1 | 3755 | potassium voltage-gated channe | 0.67709709 | 0.02013471 |
| ZNF239 | 8187 | zinc finger protein 239 | 0.67561874 | 0.00386458 |
| KLF11 | 8462 | Kruppel like factor 11 | 0.67514037 | 0.02762773 |
| TNFAIP3 | 7128 | TNF alpha induced protein 3 | 0.67284036 | 0.02176796 |
| GRPEL2-AS1 | 106144529 | GRPEL2 antisense RNA 1 | 0.67230679 | 0.02393189 |
| SNORA47 | 677828 | small nucleolar RNA, H/ACA box | 0.67147458 | 0.00410122 |
| BIRC6 | 57448 | baculoviral IAP repeat containing | 0.67139978 | 0.01561296 |
| ACOT2 | 10965 | acyl-CoA thioesterase 2 | 0.67067115 | 0.02707962 |
| NCOR2 | 9612 | nuclear receptor corepressor 2 | 0.67060334 | 0.00578988 |
| PRLR | 5618 | prolactin receptor | 0.67016548 | 0.01077793 |
| MDM2 | 4193 | MDM2 proto-oncogene | 0.67012207 | 0.00845929 |
| AGL | 178 | amylo-alpha-1, 6-glucosidase, 4- | 0.66930389 | 0.02014679 |
| FAH | 2184 | fumarylacetoacetate hydrolase | 0.6682907 | 0.04779134 |
| TYRO3 | 7301 | TYRO3 protein tyrosine kinase | 0.66823075 | 0.01194033 |
| DFFB | 1677 | DNA fragmentation factor subun | 0.66803648 | 0.01430078 |
| DENND6B | 414918 | DENN domain containing 6B | 0.66788431 | 0.01749727 |
| SLC25A45 | 283130 | solute carrier family 25 member | 0.66687662 | 0.0172259 |
| EIF3F | 8665 | eukaryotic translation initiation f | 0.66575927 | 0.01152695 |
| DCPS | 28960 | decapping enzyme, scavenger | 0.66513886 | 0.03670547 |
| EPPK1 | 83481 | epiplakin 1 | 0.6645744 | 0.03367897 |
| RPTOR | 57521 | regulatory associated protein of | 0.66457368 | 0.00274473 |
| R3HCC1 | 203069 | R3H domain and coiled-coil conta | 0.66386803 | 0.02730758 |
| WDR72 | 256764 | WD repeat domain 72 | 0.66339716 | 0.01400378 |
| WDR18 | 57418 | WD repeat domain 18 | 0.66268326 | 0.03680876 |
| PITPNC1 | 26207 | phosphatidylinositol transfer pro | 0.66263772 | 0.04516487 |
| ATIC | 471 | 5-aminoimidazole-4-carboxamid | 0.66248866 | 0.00976157 |
| MKKS | 8195 | McKusick-Kaufman syndrome | 0.66228483 | 0.02536567 |
| ANKH | 56172 | ANKH inorganic pyrophosphate tr | 0.66168561 | 0.02075294 |
| SNORD49A | 26800 | small nucleolar RNA, C/D box 49 | 0.66165814 | 0.00766691 |
| APBB2 | 323 | amyloid beta precursor protein b | 0.66092978 | 0.0275366 |
| LINC00662 | 148189 | long intergenic non-protein codin | 0.6605372 | 0.0475921 |
| SYBU | 55638 | syntabulin | 0.66028483 | 0.01787727 |
| NUFIP1 | 26747 | NUFIP1, FMR1 interacting protei | 0.65962246 | 0.01762837 |
| KCNQ1 | 3784 | potassium voltage-gated channe | 0.65910142 | 0.01152192 |
| CASP8 | 841 | caspase 8 | 0.65822044 | 0.01045399 |
| SMIM20 | 389203 | small integral membrane protein | 0.65807618 | 0.00878451 |
| HEXA | 3073 | hexosaminidase subunit alpha | 0.65796847 | 0.02096661 |
| ASXL3 | 80816 | additional sex combs like 3, trans | 0.6576062 | 0.03703786 |
| MTA1 | 9112 | metastasis associated 1 | 0.65728595 | 0.03184225 |
| PPL | 5493 | periplakin | 0.65707811 | 0.02266863 |
| HUWE1 | 10075 | HECT, UBA and WWE domain co | 0.65643835 | 0.01868408 |
| CLUAP1 | 23059 | clusterin associated protein 1 | 0.65635597 | 0.01477118 |
| FAM198B | 51313 | family with sequence similarity 1 | 0.65561564 | 0.0095712 |
| TRUB2 | 26995 | TruB pseudouridine synthase fam | 0.65560972 | 0.00563427 |
| TEX2 | 55852 | testis expressed 2 | 0.65548137 | 0.02393189 |
| SRBD1 | 55133 | S1 RNA binding domain 1 | 0.65486016 | 0.01176105 |
| TSPAN16 | 26526 | tetraspanin 16 | 0.65339908 | 0.03004045 |
| RHBDD1 | 84236 | rhomboid domain containing 1 | 0.65324062 | 0.01318555 |
| MBNL2 | 10150 | muscleblind like splicing regulato | 0.65280918 | 0.01252953 |
| ALDH1L1 | 10840 | aldehyde dehydrogenase 1 family | 0.65191117 | 0.02609385 |
| DLGAP1-AS2 | 84777 | DLGAP1 antisense RNA 2 | 0.65048076 | 0.03739957 |
| BORCS5 | 118426 | BLOC-1 related complex subunit | 0.65017488 | 0.00719306 |
| ATP13A4 | 84239 | ATPase 13A4 | 0.64789595 | 0.04483451 |

| | | | | |
|---|---|---|---|---|
| FBXW8 | 26259 | F-box and WD repeat domain containing 8 | 0.68303013 | 0.00550342 |
| CCDC149 | 91050 | coiled-coil domain containing 149 | 0.68284417 | 0.00302793 |
| COL18A1 | 80781 | collagen type XVIII alpha 1 chain | 0.6808538 | 0.03593442 |
| TK2 | 7084 | thymidine kinase 2, mitochondrial | 0.68048627 | 0.01598383 |
| TANC2 | 26115 | tetratricopeptide repeat, ankyrin repeat and | 0.67922386 | 0.01997606 |
| DDIT4 | 54541 | DNA damage inducible transcript 4 | 0.67823285 | 0.02950497 |
| ZNF703 | 80139 | zinc finger protein 703 | 0.67799985 | 0.00798763 |
| PIK3R1 | 5295 | phosphoinositide-3-kinase regulatory subun | 0.6773546 | 0.00511987 |
| ATP13A4 | 84239 | ATPase 13A4 | 0.67696684 | 0.03108987 |
| SCMH1 | 22955 | Scm polycomb group protein homolog 1 | 0.6762775 | 0.00361143 |
| CRNDE | 643911 | colorectal neoplasia differentially expressed | 0.6760645 | 0.038662 |
| DNAJC10 | 54431 | DnaJ heat shock protein family (Hsp40) mer | 0.67579413 | 0.00565361 |
| SNPH | 9751 | syntaphilin | 0.67511755 | 0.00251694 |
| THBS3 | 7059 | thrombospondin 3 | 0.67254455 | 0.0398527 |
| STEAP1B | 256227 | STEAP family member 1B | 0.67229992 | 0.01103297 |
| MROH8 | 140699 | maestro heat like repeat family member 8 | 0.67120268 | 0.00349816 |
| MOB1B | 92597 | MOB kinase activator 1B | 0.67119984 | 0.02342809 |
| SLC7A3 | 84889 | solute carrier family 7 member 3 | 0.67068165 | 0.03687019 |
| AURKC | 6795 | aurora kinase C | 0.67049093 | 0.02992642 |
| HCG25 | 414765 | HLA complex group 25 (non-protein coding) | 0.67037714 | 0.02969454 |
| QARS | 5859 | glutaminyl-tRNA synthetase | 0.67017749 | 0.00997376 |
| PRKAB1 | 5564 | protein kinase AMP-activated non-catalytic | 0.66981533 | 0.00278167 |
| SDAD1 | 55153 | SDA1 domain containing 1 | 0.66963043 | 0.0070524 |
| GLIS3 | 169792 | GLIS family zinc finger 3 | 0.668849 | 0.00705024 |
| CFB | 629 | complement factor B | 0.66864224 | 0.00261623 |
| LZTFL1 | 54585 | leucine zipper transcription factor like 1 | 0.66833668 | 0.03705029 |
| ENC1 | 8507 | ectodermal-neural cortex 1 | 0.66741536 | 0.0158167 |
| CAPN3 | 825 | calpain 3 | 0.66691533 | 0.01506465 |
| NYNRIN | 57523 | NYN domain and retroviral integrase contai | 0.66671383 | 0.04668879 |
| CASC11 | 100270680 | cancer susceptibility 11 (non-protein coding | 0.66657109 | 0.04627661 |
| ADAR | 103 | adenosine deaminase, RNA specific | 0.66602636 | 0.01431609 |
| SLC38A2 | 54407 | solute carrier family 38 member 2 | 0.66548867 | 0.00834812 |
| FAM98B | 283742 | family with sequence similarity 98 member | 0.66524545 | 0.01938765 |
| R3HDM2 | 22864 | R3H domain containing 2 | 0.66520638 | 0.00783112 |
| CD1B | 910 | CD1b molecule | 0.66490899 | 0.00182614 |
| DDX10 | 1662 | DEAD-box helicase 10 | 0.66421634 | 0.02558959 |
| LRPPRC | 10128 | leucine rich pentatricopeptide repeat contai | 0.6642057 | 0.00710066 |
| FAM198B | 51313 | family with sequence similarity 198 membe | 0.66418385 | 0.00558798 |
| ZNF254 | 9534 | zinc finger protein 254 | 0.66408308 | 0.00215339 |
| NFIC | 4782 | nuclear factor I C | 0.66381283 | 0.00360237 |
| NOA1 | 84273 | nitric oxide associated 1 | 0.66330772 | 0.02006959 |
| INPP4B | 8821 | inositol polyphosphate-4-phosphatase type | 0.66096326 | 0.01337344 |
| BARX2 | 8538 | BARX homeobox 2 | 0.65959802 | 0.01864434 |
| SESN2 | 83667 | sestrin 2 | 0.65508376 | 0.00740245 |
| ALDH1L1 | 10840 | aldehyde dehydrogenase 1 family member | 0.65423374 | 0.02008125 |
| TMEM63A | 9725 | transmembrane protein 63A | 0.65376389 | 0.00515273 |
| OSBPL9 | 114883 | oxysterol binding protein like 9 | 0.65315955 | 0.03716755 |
| PHC1 | 1911 | polyhomeotic homolog 1 | 0.65273313 | 0.03864103 |
| RGL1 | 23179 | ral guanine nucleotide dissociation stimulat | 0.64979119 | 0.02623076 |
| PTCHD4 | 442213 | patched domain containing 4 | 0.64824578 | 0.01767177 |
| AAED1 | 195827 | AhpC/TSA antioxidant enzyme domain cont | 0.64773678 | 0.03705029 |
| SNORD116-1 | 727708 | small nucleolar RNA, C/D box 116-19 | 0.64753949 | 0.03613612 |
| NUDT3 | 11165 | nudix hydrolase 3 | 0.64685717 | 0.00329894 |
| MTSS1L | 92154 | MTSS1L, I-BAR domain containing | 0.64627496 | 0.00404372 |
| ZRANB3 | 84083 | zinc finger RANBP2-type containing 3 | 0.64268175 | 0.02783287 |
| NFIA | 4774 | nuclear factor I A | 0.6412425 | 0.00552578 |
| PIGL | 9487 | phosphatidylinositol glycan anchor biosynth | 0.64117314 | 0.03428497 |
| RAP1GAP2 | 23108 | RAP1 GTPase activating protein 2 | 0.64042125 | 0.004053 |
| NUDT6 | 11162 | nudix hydrolase 6 | 0.63987871 | 0.04342748 |
| GLP1R | 2740 | glucagon like peptide 1 receptor | 0.63894465 | 0.02834993 |
| EIF3F | 8665 | eukaryotic translation initiation factor 3 sub | 0.63889753 | 0.00944718 |
| SST | 6750 | somatostatin | 0.6381983 | 0.03075055 |
| ABCC3 | 8714 | ATP binding cassette subfamily C member 3 | 0.63804537 | 0.00643942 |
| COPG2 | 26958 | coatomer protein complex subunit gamma | 0.63707559 | 0.03031073 |
| CRACR2A | 84766 | calcium release activated channel regulator | 0.63664974 | 0.02832247 |
| GYG2 | 8908 | glycogenin 2 | 0.63611267 | 0.01295555 |
| TBC1D16 | 125058 | TBC1 domain family member 16 | 0.63574198 | 0.00261623 |
| TNFRSF10D | 8793 | TNF receptor superfamily member 10d | 0.63556704 | 0.01666103 |
| OTUB2 | 78990 | OTU deubiquitinase, ubiquitin aldehyde bin | 0.63481749 | 0.0158275 |
| PPP3CA | 5530 | protein phosphatase 3 catalytic subunit alph | 0.63251517 | 0.00185005 |
| SULF2 | 55959 | sulfatase 2 | 0.63221369 | 0.02150249 |
| TNKS | 8658 | tankyrase | 0.6321854 | 0.01488624 |
| TNFRSF19 | 55504 | TNF receptor superfamily member 19 | 0.63162332 | 0.04324391 |
| SLC35F2 | 54733 | solute carrier family 35 member F2 | 0.62973278 | 0.02938077 |

| | | | | |
|---|---|---|---|---|
| ATP1B4 | 23439 | ATPase Na+/K+ transporting fam | 0.64700725 | 0.01561296 |
| CLYBL | 171425 | citrate lyase beta like | 0.64676539 | 0.03296826 |
| ZNF805 | 390980 | zinc finger protein 805 | 0.6462601 | 0.02282078 |
| POLR1A | 25885 | RNA polymerase I subunit A | 0.64620982 | 0.01902554 |
| COG5 | 10466 | component of oligomeric golgi co | 0.64578389 | 0.00828949 |
| LOXL1-AS1 | 100287616 | LOXL1 antisense RNA 1 | 0.64427397 | 0.03739957 |
| LRIG3 | 121227 | leucine rich repeats and immuno | 0.64423137 | 0.01301089 |
| DCLRE1B | 64858 | DNA cross-link repair 1B | 0.64361426 | 0.04171616 |
| TRAP1 | 10131 | TNF receptor associated protein | 0.64311341 | 0.04349882 |
| LINC01163 | 101927381 | long intergenic non-protein codir | 0.64308526 | 0.01570928 |
| SARM1 | 23098 | sterile alpha and TIR motif conta | 0.64287372 | 0.02042137 |
| C1QTNF3-AM | 100534612 | C1QTNF3-AMACR readthrough (N | 0.64195399 | 0.00753522 |
| MRPS18B | 28973 | mitochondrial ribosomal protein | 0.64195351 | 0.01070603 |
| ZFAND1 | 79752 | zinc finger AN1-type containing 1 | 0.64078229 | 0.00701532 |
| MTO1 | 25821 | mitochondrial tRNA translation o | 0.6402286 | 0.02333125 |
| PIH1D1 | 55011 | PIH1 domain containing 1 | 0.64004862 | 0.01004469 |
| ATE1 | 11101 | arginyltransferase 1 | 0.63992144 | 0.0229306 |
| ORAOV1 | 220064 | oral cancer overexpressed 1 | 0.63915848 | 0.02211568 |
| SDE2 | 163859 | SDE2 telomere maintenance hon | 0.63877613 | 0.01882443 |
| ANKRA2 | 57763 | ankyrin repeat family A member | 0.63806565 | 0.02385708 |
| COX7A2L | 9167 | cytochrome c oxidase subunit 7A | 0.63791754 | 0.00463362 |
| SNPH | 9751 | syntaphilin | 0.63770839 | 0.00591603 |
| RRS1 | 23212 | ribosome biogenesis regulator ho | 0.63722324 | 0.03992953 |
| LYPD6B | 130576 | LY6/PLAUR domain containing 6I | 0.63473524 | 0.0135338 |
| UROS | 7390 | uroporphyrinogen III synthase | 0.63459454 | 0.0229306 |
| NRP1 | 8829 | neuropilin 1 | 0.63447892 | 0.02437922 |
| MOSPD1 | 56180 | motile sperm domain containing | 0.63427044 | 0.03024505 |
| PKIG | 11142 | cAMP-dependent protein kinase | 0.63410776 | 0.03640358 |
| ENAH | 55740 | ENAH, actin regulator | 0.63408777 | 0.02093051 |
| CHN2 | 1124 | chimerin 2 | 0.63312085 | 0.02595326 |
| ZNF442 | 79973 | zinc finger protein 442 | 0.63230029 | 0.04293624 |
| RGMB | 285704 | repulsive guidance molecule BM | 0.63104041 | 0.02614213 |
| SLC35F2 | 54733 | solute carrier family 35 member | 0.63096777 | 0.03626746 |
| ZNF460 | 10794 | zinc finger protein 460 | 0.63070396 | 0.00475156 |
| SFXN4 | 119559 | sideroflexin 4 | 0.63063029 | 0.01499376 |
| KRTAP5-AS1 | 338651 | KRTAP5-1/KRTAP5-2 antisense R | 0.63057873 | 0.01385154 |
| TK2 | 7084 | thymidine kinase 2, mitochondria | 0.62940418 | 0.02979232 |
| USP42 | 84132 | ubiquitin specific peptidase 42 | 0.6279342 | 0.02619168 |
| PTPRU | 10076 | protein tyrosine phosphatase, rec | 0.6278326 | 0.01426383 |
| RNF144B | 255488 | ring finger protein 144B | 0.62756553 | 0.02486359 |
| AVEN | 57099 | apoptosis and caspase activation | 0.62694181 | 0.00866109 |
| TACO1 | 51204 | translational activator of cytochr | 0.62642 | 0.0089459 |
| MTR | 4548 | 5-methyltetrahydrofolate-homoc | 0.62568194 | 0.01500454 |
| WDR43 | 23160 | WD repeat domain 43 | 0.62355149 | 0.01028522 |
| PYGB | 5834 | glycogen phosphorylase B | 0.6222368 | 0.00602051 |
| FBXL17 | 64839 | F-box and leucine rich repeat pro | 0.62073135 | 0.03396418 |
| FARSB | 10056 | phenylalanyl-tRNA synthetase be | 0.62068106 | 0.01152192 |
| TCN2 | 6948 | transcobalamin 2 | 0.62062712 | 0.03752931 |
| DCUN1D2 | 55208 | defective in cullin neddylation 1 ( | 0.62039559 | 0.04552744 |
| DET1 | 55070 | DET1, COP1 ubiquitin ligase partr | 0.62016031 | 0.02493731 |
| TMPRSS3 | 64699 | transmembrane serine protease | 0.61896901 | 0.03633157 |
| SCUBE2 | 57758 | signal peptide, CUB domain and | 0.61732375 | 0.02155779 |
| DZIP3 | 9666 | DAZ interacting zinc finger prote | 0.61695367 | 0.03739957 |
| RPS4Y1 | 6192 | ribosomal protein S4, Y-linked 1 | 0.61465152 | 0.01070603 |
| NOA1 | 84273 | nitric oxide associated 1 | 0.61464575 | 0.03507443 |
| ORC3 | 23595 | origin recognition complex subur | 0.61448394 | 0.02944652 |
| CNNM1 | 26507 | cyclin and CBS domain divalent n | 0.61304892 | 0.02824866 |
| HEATR1 | 55127 | HEAT repeat containing 1 | 0.61243984 | 0.01861591 |
| GPR143 | 4935 | G protein-coupled receptor 143 | 0.61229624 | 0.04639051 |
| LRRC8D | 55144 | leucine rich repeat containing 8 V | 0.61196762 | 0.01115265 |
| CEP57 | 9702 | centrosomal protein 57 | 0.61150892 | 0.03669796 |
| BARX2 | 8538 | BARX homeobox 2 | 0.61023049 | 0.03321763 |
| RBMS1 | 5937 | RNA binding motif single strande | 0.60983681 | 0.02637176 |
| ABCA3 | 21 | ATP binding cassette subfamily A | 0.60981018 | 0.03992953 |
| MICALL1 | 85377 | MICAL like 1 | 0.6093974 | 0.00576562 |
| RBM43 | 375287 | RNA binding motif protein 43 | 0.60875852 | 0.03025936 |
| TACC2 | 10579 | transforming acidic coiled-coil co | 0.60634404 | 0.01356979 |
| CYC1 | 1537 | cytochrome c1 | 0.60624751 | 0.00433353 |
| OSBP2 | 23762 | oxysterol binding protein 2 | 0.60532659 | 0.02536567 |
| SMCHD1 | 23347 | structural maintenance of chrom | 0.60471506 | 0.00719306 |
| RNLS | 55328 | renalase, FAD dependent amine | 0.60441256 | 0.04490739 |
| UHRF1BP1 | 54887 | UHRF1 binding protein 1 | 0.60401936 | 0.03985085 |
| PDSS1 | 23590 | decaprenyl diphosphate synthase | 0.60372945 | 0.01886764 |
| PDK2 | 5164 | pyruvate dehydrogenase kinase 2 | 0.60352809 | 0.03571342 |

| | | | | |
|---|---|---|---|---|
| SRD5A2 | 6716 | steroid 5 alpha-reductase 2 | 0.62947193 | 0.03516166 |
| SHC3 | 53358 | SHC adaptor protein 3 | 0.62925278 | 0.01835347 |
| SUPT3H | 8464 | SPT3 homolog, SAGA and STAGA complex c | 0.62883725 | 0.01538782 |
| FASLG | 356 | Fas ligand | 0.62867767 | 0.03907963 |
| UBLCP1 | 134510 | ubiquitin like domain containing CTD phosph | 0.62832187 | 0.02576239 |
| ARHGAP23 | 57636 | Rho GTPase activating protein 23 | 0.6280314 | 0.0176175 |
| IVD | 3712 | isovaleryl-CoA dehydrogenase | 0.62621478 | 0.04201122 |
| RNF20 | 56254 | ring finger protein 20 | 0.62570412 | 0.00671844 |
| SH2D7 | 646892 | SH2 domain containing 7 | 0.6255618 | 0.02470951 |
| MARS | 4141 | methionyl-tRNA synthetase | 0.62467225 | 0.0056754 |
| ZNF358 | 140467 | zinc finger protein 358 | 0.62449363 | 0.03697445 |
| CDKN1B | 1027 | cyclin dependent kinase inhibitor 1B | 0.62436898 | 0.00727264 |
| MOCOS | 55034 | molybdenum cofactor sulfurase | 0.62414695 | 0.02299137 |
| BARHL1 | 56751 | BarH like homeobox 1 | 0.61996328 | 0.00813765 |
| MYO10 | 4651 | myosin X | 0.61968369 | 0.020645 |
| HSPG2 | 3339 | heparan sulfate proteoglycan 2 | 0.61905217 | 0.02147687 |
| PCM1 | 5108 | pericentriolar material 1 | 0.61896477 | 0.00395032 |
| NOXO1 | 124056 | NADPH oxidase organizer 1 | 0.61844885 | 0.01138615 |
| BEST1 | 7439 | bestrophin 1 | 0.61810666 | 0.04552835 |
| RASA1 | 5921 | RAS p21 protein activator 1 | 0.6176705 | 0.01538782 |
| CD5L | 922 | CD5 molecule like | 0.61750916 | 0.04396328 |
| ZMYM2 | 7750 | zinc finger MYM-type containing 2 | 0.61663061 | 0.00349649 |
| LINC01363 | 101928484 | long intergenic non-protein coding RNA 136 | 0.61429704 | 0.0355627 |
| HSPA9 | 3313 | heat shock protein family A (Hsp70) membe | 0.61403647 | 0.00381105 |
| TNRC6C-AS1 | 100131096 | TNRC6C antisense RNA 1 | 0.61370994 | 0.04873077 |
| PTPN12 | 5782 | protein tyrosine phosphatase, non-receptor | 0.61173502 | 0.02875237 |
| SLC38A7 | 55238 | solute carrier family 38 member 7 | 0.60993111 | 0.03343719 |
| RRN3 | 54700 | RRN3 homolog, RNA polymerase I transcrip | 0.6089835 | 0.01575344 |
| NBR2 | 10230 | neighbor of BRCA1 gene 2 (non-protein codi | 0.60888614 | 0.00671843 |
| LAMC1 | 3915 | laminin subunit gamma 1 | 0.60872557 | 0.03716755 |
| COBLL1 | 22837 | cordon-bleu WH2 repeat protein like 1 | 0.60869793 | 0.0049046 |
| CUEDC1 | 404093 | CUE domain containing 1 | 0.60840944 | 0.00836422 |
| FAM131B | 9715 | family with sequence similarity 131 membe | 0.60794344 | 0.0432028 |
| TRIM22 | 10346 | tripartite motif containing 22 | 0.60734945 | 0.03011273 |
| TH2LCRR | 101927761 | T helper type 2 locus control region associat | 0.60655035 | 0.04143086 |
| ZMIZ2 | 83637 | zinc finger MIZ-type containing 2 | 0.60621047 | 0.00953026 |
| YBX3 | 8531 | Y-box binding protein 3 | 0.60556369 | 0.00833126 |
| WDR74 | 54663 | WD repeat domain 74 | 0.60502864 | 0.01424128 |
| MCCC1 | 56922 | methylcrotonoyl-CoA carboxylase 1 | 0.60445128 | 0.02470951 |
| RGMB | 285704 | repulsive guidance molecule BMP co-recept | 0.60419687 | 0.0249432 |
| ARHGEF10L | 55160 | Rho guanine nucleotide exchange factor 10 | 0.603452 | 0.0144926 |
| CCPG1 | 9236 | cell cycle progression 1 | 0.60339999 | 0.04093753 |
| TIMM44 | 10469 | translocase of inner mitochondrial membra | 0.60234916 | 0.02234538 |
| ANKRD29 | 147463 | ankyrin repeat domain 29 | 0.60217601 | 0.04729193 |
| KIF2A | 3796 | kinesin family member 2A | 0.60179674 | 0.02033445 |
| SLC39A10 | 57181 | solute carrier family 39 member 10 | 0.60088873 | 0.02812874 |
| NACA2 | 342538 | nascent polypeptide associated complex alp | 0.60006384 | 0.02040829 |
| UBE2E1 | 7324 | ubiquitin conjugating enzyme E2 E1 | 0.59966196 | 0.00899302 |
| MR1 | 3140 | major histocompatibility complex, class I-re | 0.5985598 | 0.01189036 |
| RSL1D1 | 26156 | ribosomal L1 domain containing 1 | 0.59840031 | 0.00831486 |
| PYCR1 | 5831 | pyrroline-5-carboxylate reductase 1 | 0.59827781 | 0.01370992 |
| GNL2 | 29889 | G protein nucleolar 2 | 0.59759084 | 0.01732472 |
| SYBU | 55638 | syntabulin | 0.59605787 | 0.0217081 |
| H2AFY2 | 55506 | H2A histone family member Y2 | 0.59573259 | 0.01283756 |
| GRPEL2 | 134266 | GrpE like 2, mitochondrial | 0.59509236 | 0.01288751 |
| ZNF415 | 55786 | zinc finger protein 415 | 0.5946732 | 0.04093753 |
| BTC | 685 | betacellulin | 0.59453504 | 0.02428763 |
| ARHGEF38 | 54848 | Rho guanine nucleotide exchange factor 38 | 0.59450186 | 0.04255404 |
| CHD4 | 1108 | chromodomain helicase DNA binding protei | 0.59379384 | 0.00662572 |
| LHX2 | 9355 | LIM homeobox 2 | 0.59203594 | 0.03492026 |
| ARSD | 414 | arylsulfatase D | 0.58976256 | 0.02319498 |
| EIF2S2 | 8894 | eukaryotic translation initiation factor 2 sub | 0.58942656 | 0.00903722 |
| LRIF1 | 55791 | ligand dependent nuclear receptor interacti | 0.58842301 | 0.02407515 |
| PDLIM1 | 9124 | PDZ and LIM domain 1 | 0.58795965 | 0.00299916 |
| DDX42 | 11325 | DEAD-box helicase 42 | 0.58768535 | 0.00313687 |
| TBL1Y | 90665 | transducin beta like 1 Y-linked | 0.58698921 | 0.01133118 |
| PPP1R12B | 4660 | protein phosphatase 1 regulatory subunit 12 | 0.58610993 | 0.04355394 |
| DMTN | 2039 | dematin actin binding protein | 0.58586219 | 0.0093092 |
| SLC22A3 | 6581 | solute carrier family 22 member 3 | 0.58525023 | 0.01898482 |
| IDE | 3416 | insulin degrading enzyme | 0.5846654 | 0.0225104 |
| LRIG2 | 9860 | leucine rich repeats and immunoglobulin lik | 0.58441499 | 0.02297833 |
| HINT2 | 84681 | histidine triad nucleotide binding protein 2 | 0.58369403 | 0.01575344 |
| RABL2A | 11159 | RAB, member of RAS oncogene family like 2 | 0.58346648 | 0.00944718 |
| ILF3 | 3609 | interleukin enhancer binding factor 3 | 0.58340098 | 0.00567259 |

| | | | | |
|---|---|---|---|---|
| PDAP1 | 11333 | PDGFA associated protein 1 | 0.60301658 | 0.01561296 |
| SNORD92 | 692209 | small nucleolar RNA, C/D box 92 | 0.60269355 | 0.03582173 |
| ZBTB25 | 7597 | zinc finger and BTB domain cont | 0.60184661 | 0.04910168 |
| CYFIP2 | 26999 | cytoplasmic FMR1 interacting pr | 0.60176134 | 0.01818588 |
| PLCL2 | 23228 | phospholipase C like 2 | 0.60125094 | 0.00297808 |
| RSL24D1 | 51187 | ribosomal L24 domain containing | 0.60112201 | 0.00774445 |
| CCDC59 | 29080 | coiled-coil domain containing 59 | 0.59974647 | 0.02196219 |
| MRPL15 | 29088 | mitochondrial ribosomal protein | 0.59922398 | 0.03363763 |
| ACTA2 | 59 | actin, alpha 2, smooth muscle, a | 0.59913576 | 0.04216373 |
| POFUT1 | 23509 | protein O-fucosyltransferase 1 | 0.59900997 | 0.03739957 |
| PRKAB1 | 5564 | protein kinase AMP-activated no | 0.59800574 | 0.0089459 |
| SMARCA2 | 6595 | SWI/SNF related, matrix associa | 0.5978697 | 0.04293624 |
| CSGALNACT2 | 55454 | chondroitin sulfate N-acetylgalac | 0.59681608 | 0.01749727 |
| MAPKAPK3 | 7867 | mitogen-activated protein kinase | 0.59618777 | 0.02287353 |
| ABL1 | 25 | ABL proto-oncogene 1, non-recep | 0.59589932 | 0.03202817 |
| TRRAP | 8295 | transformation/transcription dor | 0.59503536 | 0.03986046 |
| COPG2 | 26958 | coatomer protein complex subun | 0.59483942 | 0.0480692 |
| ZFP62 | 643836 | ZFP62 zinc finger protein | 0.59370658 | 0.01992218 |
| SIAH1 | 6477 | siah E3 ubiquitin protein ligase 1 | 0.59338727 | 0.0270379 |
| DESI1 | 27351 | desumoylating isopeptidase 1 | 0.59216904 | 0.01416068 |
| WDR12 | 55759 | WD repeat domain 12 | 0.59201397 | 0.04950029 |
| MAK16 | 84549 | MAK16 homolog | 0.59153519 | 0.01262385 |
| RPF2 | 84154 | ribosome production factor 2 hor | 0.59083568 | 0.0095712 |
| WDR45 | 11152 | WD repeat domain 45 | 0.58916952 | 0.0089459 |
| BEND3 | 57673 | BEN domain containing 3 | 0.58891616 | 0.04471799 |
| ITGB4 | 3691 | integrin subunit beta 4 | 0.58865184 | 0.0229306 |
| NSMAF | 8439 | neutral sphingomyelinase activat | 0.58827835 | 0.02306633 |
| APC | 324 | APC, WNT signaling pathway reg | 0.58809721 | 0.02906968 |
| FAM171A1 | 221061 | family with sequence similarity 1 | 0.58781373 | 0.02393189 |
| GFPT1 | 2673 | glutamine--fructose-6-phosphate | 0.58777012 | 0.01787727 |
| SEPSECS | 51091 | Sep (O-phosphoserine) tRNA:Sec | 0.58776098 | 0.02912298 |
| WDR74 | 54663 | WD repeat domain 74 | 0.58753365 | 0.02210353 |
| CERK | 64781 | ceramide kinase | 0.5872 | 0.03604565 |
| EIF6 | 3692 | eukaryotic translation initiation f | 0.58713103 | 0.00861386 |
| TBC1D16 | 125058 | TBC1 domain family member 16 | 0.58709802 | 0.00719306 |
| ELP3 | 55140 | elongator acetyltransferase com | 0.58701338 | 0.00792319 |
| SEMA4C | 54910 | semaphorin 4C | 0.58636638 | 0.0182953 |
| MRPS22 | 56945 | mitochondrial ribosomal protein | 0.58619146 | 0.00591603 |
| AKAP2 | 11217 | A-kinase anchoring protein 2 | 0.58481514 | 0.03330983 |
| PGPEP1 | 54858 | pyroglutamyl-peptidase I | 0.58475803 | 0.03703786 |
| PHB2 | 11331 | prohibitin 2 | 0.58401543 | 0.02824866 |
| ZFR | 51663 | zinc finger RNA binding protein | 0.5835498 | 0.00766691 |
| PLEKHG3 | 26030 | pleckstrin homology and RhoGEF | 0.58329655 | 0.03294259 |
| CENPBD1 | 92806 | CENPB DNA-binding domain cont | 0.58278418 | 0.04880324 |
| F2RL2 | 2151 | coagulation factor II thrombin re | 0.58167797 | 0.04496852 |
| NAA10 | 8260 | N(alpha)-acetyltransferase 10, N | 0.58163032 | 0.03707738 |
| FBRSL1 | 57666 | fibrosin like 1 | 0.58133793 | 0.00991595 |
| CLUH | 23277 | clustered mitochondria homolog | 0.581311 | 0.01212382 |
| CAD | 790 | carbamoyl-phosphate synthetase | 0.5812858 | 0.01003914 |
| HSPG2 | 3339 | heparan sulfate proteoglycan 2 | 0.5805595 | 0.03507443 |
| TBC1D24 | 57465 | TBC1 domain family member 24 | 0.58022283 | 0.03110207 |
| GLUL | 2752 | glutamate-ammonia ligase | -0.5801971 | 0.00962575 |
| MIR5589 | 100847093 | microRNA 5589 | -0.5817586 | 0.03728968 |
| STAG3L5P-PV | 101752399 | STAG3L5P-PVRIG2P-PILRB readt | -0.5825124 | 0.04589502 |
| CIDEB | 27141 | cell death-inducing DFFA-like eff | -0.5851913 | 0.04446091 |
| ANXA5 | 308 | annexin A5 | -0.5916817 | 0.02402969 |
| SAMD9 | 54809 | sterile alpha motif domain conta | -0.5955318 | 0.01143723 |
| FDPS | 2224 | farnesyl diphosphate synthase | -0.5988722 | 0.00526073 |
| ACSS2 | 55902 | acyl-CoA synthetase short chain f | -0.5996211 | 0.01428227 |
| ADI1 | 55256 | acireductone dioxygenase 1 | -0.6041289 | 0.02176796 |
| VILL | 50853 | villin like | -0.6057576 | 0.01196688 |
| ACSL1 | 2180 | acyl-CoA synthetase long chain fa | -0.609059 | 0.01609673 |
| LINC01363 | 101928484 | long intergenic non-protein codir | -0.6131725 | 0.04251199 |
| TEX30 | 93081 | testis expressed 30 | -0.6189085 | 0.03024505 |
| ADIRF | 10974 | adipogenesis regulatory factor | -0.6198006 | 0.02855361 |
| ACVR1C | 130399 | activin A receptor type 1C | -0.6214899 | 0.0078673 |
| SQLE | 6713 | squalene epoxidase | -0.6261472 | 0.0160321 |
| MSMO1 | 6307 | methylsterol monooxygenase 1 | -0.6383592 | 0.00870677 |
| ATP8B2 | 57198 | ATPase phospholipid transporting | -0.6393485 | 0.02567596 |
| VSIG1 | 340547 | V-set and immunoglobulin doma | -0.6433137 | 0.04797213 |
| ARMCX1 | 51309 | armadillo repeat containing, X-lir | -0.6482035 | 0.00538163 |
| PNMA1 | 9240 | PNMA family member 1 | -0.6485127 | 0.01250152 |
| GNAZ | 2781 | G protein subunit alpha z | -0.6488811 | 0.04078367 |
| LSS | 4047 | lanosterol synthase | -0.6498611 | 0.02144448 |

| | | | | |
|---|---|---|---|---|
| CDC16 | 8881 | cell division cycle 16 | 0.58332445 | 0.00565361 |
| ZNF181 | 339318 | zinc finger protein 181 | 0.58298474 | 0.023201 |
| EMILIN1 | 11117 | elastin microfibril interfacer 1 | 0.58259285 | 0.01767177 |
| WNK2 | 65268 | WNK lysine deficient protein kinase 2 | 0.58253934 | 0.02006959 |
| EIF2S3 | 1968 | eukaryotic translation initiation factor 2 sub | 0.58159865 | 0.01059843 |
| HNF1B | 6928 | HNF1 homeobox B | 0.58135565 | 0.04552835 |
| PCCA | 5095 | propionyl-CoA carboxylase alpha subunit | 0.58096384 | 0.02818064 |
| ACSL1 | 2180 | acyl-CoA synthetase long chain family mem | -0.5804313 | 0.01488624 |
| IL32 | 9235 | interleukin 32 | -0.5817584 | 0.01584898 |
| ENTPD7 | 57089 | ectonucleoside triphosphate diphosphohydr | -0.5823508 | 0.02043838 |
| PROSER2-AS | 219731 | PROSER2 antisense RNA 1 | -0.5823538 | 0.03697124 |
| RAP1A | 5906 | RAP1A, member of RAS oncogene family | -0.5824551 | 0.03098601 |
| AKR1B1 | 231 | aldo-keto reductase family 1 member B | -0.5838794 | 0.03628016 |
| APOC2 | 344 | apolipoprotein C2 | -0.5850224 | 0.04169343 |
| ABHD5 | 51099 | abhydrolase domain containing 5 | -0.5852376 | 0.02296854 |
| SEC23A | 10484 | Sec23 homolog A, coat complex II compone | -0.5875633 | 0.03586163 |
| RAB8B | 51762 | RAB8B, member RAS oncogene family | -0.5877758 | 0.01189036 |
| ABHD6 | 57406 | abhydrolase domain containing 6 | -0.5886202 | 0.03886631 |
| TNNT1 | 7138 | troponin T1, slow skeletal type | -0.589705 | 0.0299653 |
| CEACAM6 | 4680 | carcinoembryonic antigen related cell adhes | -0.5901478 | 0.00248918 |
| PDLIM2 | 64236 | PDZ and LIM domain 2 | -0.5907717 | 0.02158936 |
| AKR7A2 | 8574 | aldo-keto reductase family 7 member A2 | -0.5910367 | 0.00398531 |
| AGPAT2 | 10555 | 1-acylglycerol-3-phosphate O-acyltransferas | -0.5933914 | 0.01138615 |
| RHBDL2 | 54933 | rhomboid like 2 | -0.5937258 | 0.01538918 |
| LRRC66 | 339977 | leucine rich repeat containing 66 | -0.5944464 | 0.01980077 |
| LRRC3 | 81543 | leucine rich repeat containing 3 | -0.5953838 | 0.0139131 |
| SS18L2 | 51188 | SS18 like 2 | -0.5953935 | 0.02589349 |
| MVD | 4597 | mevalonate diphosphate decarboxylase | -0.5954029 | 0.00501884 |
| AGR3 | 155465 | anterior gradient 3, protein disulphide isom | -0.5955665 | 0.01764373 |
| SLX1A | 548593 | SLX1 homolog A, structure-specific endonuc | -0.5966239 | 0.01577201 |
| RASA4B | 100271927 | RAS p21 protein activator 4B | -0.5966753 | 0.01387043 |
| NIPAL1 | 152519 | NIPA like domain containing 1 | -0.5972662 | 0.00686509 |
| SLC2A14 | 144195 | solute carrier family 2 member 14 | -0.5975018 | 0.03700889 |
| ST3GAL1 | 6482 | ST3 beta-galactoside alpha-2,3-sialyltransfe | -0.5995305 | 0.01613569 |
| ABCA5 | 23461 | ATP binding cassette subfamily A member 5 | -0.6050585 | 0.02756235 |
| MTM1 | 4534 | myotubularin 1 | -0.6062328 | 0.02158015 |
| FUCA1 | 2517 | alpha-L-fucosidase 1 | -0.6062671 | 0.00648967 |
| GDPD1 | 284161 | glycerophosphodiester phosphodiesterase d | -0.6064186 | 0.02190605 |
| ACOX2 | 8309 | acyl-CoA oxidase 2 | -0.6065277 | 0.01042379 |
| RAB27A | 5873 | RAB27A, member RAS oncogene family | -0.6071672 | 0.0139131 |
| CD68 | 968 | CD68 molecule | -0.6077817 | 0.01611112 |
| SLC6A4 | 6532 | solute carrier family 6 member 4 | -0.6103971 | 0.03372039 |
| IGFBP4 | 3487 | insulin like growth factor binding protein 4 | -0.6106046 | 0.02232583 |
| BMP2 | 650 | bone morphogenetic protein 2 | -0.6111782 | 0.02786643 |
| STARD4 | 134429 | StAR related lipid transfer domain containir | -0.6112738 | 0.00261623 |
| CCL23 | 6368 | C-C motif chemokine ligand 23 | -0.6113243 | 0.01142306 |
| RELL1 | 768211 | RELT like 1 | -0.6161077 | 0.01165856 |
| SNRNP25 | 79622 | small nuclear ribonucleoprotein U11/U12 su | -0.6161318 | 0.017129 |
| STPG2-AS1 | 101410545 | STPG2 antisense RNA 1 | -0.6162133 | 0.0231401 |
| HAS3 | 3038 | hyaluronan synthase 3 | -0.6171559 | 0.00969052 |
| BDKRB2 | 624 | bradykinin receptor B2 | -0.6182663 | 0.04199827 |
| SMPD1 | 6609 | sphingomyelin phosphodiesterase 1 | -0.6194305 | 0.01152369 |
| FAM3C | 10447 | family with sequence similarity 3 member C | -0.6201536 | 0.04093753 |
| FDPS | 2224 | farnesyl diphosphate synthase | -0.620463 | 0.00248918 |
| ADGRL3 | 23284 | adhesion G protein-coupled receptor L3 | -0.6208929 | 0.03826803 |
| LINC01133 | 100505633 | long intergenic non-protein coding RNA 113 | -0.6218831 | 0.00771631 |
| SLC40A1 | 30061 | solute carrier family 40 member 1 | -0.6227631 | 0.02604542 |
| SLC20A1 | 6574 | solute carrier family 20 member 1 | -0.623791 | 0.0146251 |
| OR10W1 | 81341 | olfactory receptor family 10 subfamily W m | -0.6240327 | 0.01611112 |
| CNIH4 | 29097 | cornichon family AMPA receptor auxiliary pr | -0.6262114 | 0.00343595 |
| PROCR | 10544 | protein C receptor | -0.6266214 | 0.02470951 |
| PNP | 4860 | purine nucleoside phosphorylase | -0.6284363 | 0.00319103 |
| MIR3909 | 100500826 | microRNA 3909 | -0.6290651 | 0.01993306 |
| DQX1 | 165545 | DEAQ-box RNA dependent ATPase 1 | -0.633405 | 0.01284766 |
| DDX60 | 55601 | DExD/H-box helicase 60 | -0.6334345 | 0.0146251 |
| HTATIP2 | 10553 | HIV-1 Tat interactive protein 2 | -0.6359975 | 0.01079591 |
| EPHX2 | 2053 | epoxide hydrolase 2 | -0.6402871 | 0.0062458 |
| PCSK5 | 5125 | proprotein convertase subtilisin/kexin type 5 | -0.6404324 | 0.03546965 |
| HLA-F-AS1 | 285830 | HLA-F antisense RNA 1 | -0.641197 | 0.00722518 |
| SOX21 | 11166 | SRY-box 21 | -0.6412693 | 0.00282706 |
| DGAT2 | 84649 | diacylglycerol O-acyltransferase 2 | -0.6414055 | 0.01143299 |
| REEP3 | 221035 | receptor accessory protein 3 | -0.6423374 | 0.00899302 |
| AKR7A3 | 22977 | aldo-keto reductase family 7 member A3 | -0.6427413 | 0.00141158 |
| BMP8B | 656 | bone morphogenetic protein 8b | -0.6439645 | 0.04515191 |

| | | | | |
|---|---|---|---|---|
| KCNS3 | 3790 | potassium voltage-gated channe | -0.6570666 | 0.00624999 |
| INSIG1 | 3638 | insulin induced gene 1 | -0.659974 | 0.00480276 |
| SLC6A12 | 6539 | solute carrier family 6 member 1 | -0.661491 | 0.01724339 |
| ACER2 | 340485 | alkaline ceramidase 2 | -0.662819 | 0.02439953 |
| APOC1 | 341 | apolipoprotein C1 | -0.6651442 | 0.00334678 |
| IFNE | 338376 | interferon epsilon | -0.6743123 | 0.01123185 |
| PTPN22 | 26191 | protein tyrosine phosphatase, no | -0.6756279 | 0.03669796 |
| MIR580 | 693165 | microRNA 580 | -0.6826722 | 0.03873331 |
| SC5D | 6309 | sterol-C5-desaturase | -0.6843449 | 0.01301089 |
| RNU4ATAC | 100151683 | RNA, U4atac small nuclear (U12 | -0.6878323 | 0.00605733 |
| LINC00458 | 100507428 | long intergenic non-protein codir | -0.6927216 | 0.04532626 |
| APOBEC1 | 339 | apolipoprotein B mRNA editing e | -0.6976593 | 0.02762297 |
| SCD | 6319 | stearoyl-CoA desaturase | -0.7076779 | 0.01787727 |
| SFRP5 | 6425 | secreted frizzled related protein | -0.7172559 | 0.04446091 |
| HMGCS1 | 3157 | 3-hydroxy-3-methylglutaryl-CoA s | -0.7303128 | 0.00173406 |
| MFGE8 | 4240 | milk fat globule-EGF factor 8 pro | -0.7363919 | 0.01340919 |
| ID3 | 3399 | inhibitor of DNA binding 3, HLH p | -0.7412768 | 0.01474453 |
| RARB | 5915 | retinoic acid receptor beta | -0.7425089 | 0.00526073 |
| CHAD | 1101 | chondroadherin | -0.7565808 | 0.02185715 |
| MIR181A1HG | 100131234 | MIR181A1 host gene | -0.7594504 | 0.03992953 |
| MIR4521 | 100616406 | microRNA 4521 | -0.7698613 | 0.02619168 |
| RAB3B | 5865 | RAB3B, member RAS oncogene f | -0.7705992 | 0.00696825 |
| P3H2 | 55214 | prolyl 3-hydroxylase 2 | -0.7760751 | 0.03806126 |
| GATM | 2628 | glycine amidinotransferase | -0.7792662 | 0.00060938 |
| LINC00431 | 104355135 | long intergenic non-protein codir | -0.7815977 | 0.02595326 |
| IFI27 | 3429 | interferon alpha inducible protei | -0.786995 | 0.00194046 |
| MXRA5 | 25878 | matrix remodeling associated 5 | -0.7877554 | 0.01146655 |
| GCNT4 | 51301 | glucosaminyl (N-acetyl) transfera | -0.7924747 | 0.03206724 |
| SLC16A3 | 9123 | solute carrier family 16 member | -0.7978019 | 0.01868408 |
| MIR548Z | 100500856 | microRNA 548z | -0.8081247 | 0.03091411 |
| PLAGL1 | 5325 | PLAG1 like zinc finger 1 | -0.8108121 | 0.02432219 |
| BIRC6-AS2 | 103752586 | BIRC6 antisense RNA 2 | -0.8354996 | 0.01176105 |
| MMP1 | 4312 | matrix metallopeptidase 1 | -0.8368963 | 0.02210353 |
| TFF1 | 7031 | trefoil factor 1 | -0.8397346 | 0.00184513 |
| MIR3662 | 100500880 | microRNA 3662 | -0.8432212 | 0.02385708 |
| SLC9A4 | 389015 | solute carrier family 9 member A | -0.8918541 | 0.04914832 |
| HHLA2 | 11148 | HERV-H LTR-associating 2 | -0.9062485 | 0.00578861 |
| LINC01372 | 101929736 | long intergenic non-protein codir | -0.9109344 | 0.00474492 |
| VSIG2 | 23584 | V-set and immunoglobulin doma | -0.9136296 | 0.00114511 |
| GABBR1 | 2550 | gamma-aminobutyric acid type E | -0.9157281 | 0.01477118 |
| GUCA2B | 2981 | guanylate cyclase activator 2B | -0.9183128 | 0.0182953 |
| ACAT2 | 39 | acetyl-CoA acetyltransferase 2 | -0.9271565 | 0.00512142 |
| SEMA6C | 10500 | semaphorin 6C | -0.9280841 | 0.01902554 |
| LRP4 | 4038 | LDL receptor related protein 4 | -0.9288951 | 0.03440088 |
| LEAP2 | 116842 | liver enriched antimicrobial pept | -0.9377786 | 0.03407614 |
| LIFR | 3977 | LIF receptor alpha | -0.9415657 | 0.02588412 |
| CLDN18 | 51208 | claudin 18 | -0.9481757 | 0.02013321 |
| UGT2B28 | 54490 | UDP glucuronosyltransferase fan | -0.959152 | 0.03643259 |
| SULT1B1 | 27284 | sulfotransferase family 1B mem | -0.9796613 | 0.03044696 |
| RBPMS2 | 348093 | RNA binding protein, mRNA proc | -0.9987614 | 0.01288736 |
| CDA | 978 | cytidine deaminase | -1.0120136 | 0.0140705 |
| HABP2 | 3026 | hyaluronan binding protein 2 | -1.0306047 | 0.0297708 |
| DKK1 | 22943 | dickkopf WNT signaling pathway | -1.0428697 | 0.03396418 |
| BAAT | 570 | bile acid-CoA:amino acid N-acylt | -1.0498188 | 0.00334678 |
| PNCK | 139728 | pregnancy up-regulated nonubiqu | -1.0546125 | 0.02595326 |
| CCDC144A | 9720 | coiled-coil domain containing 14 | -1.0703907 | 0.0338229 |
| MIR186 | 406962 | microRNA 186 | -1.0833155 | 0.01152192 |
| KLK7 | 5650 | kallikrein related peptidase 7 | -1.0978353 | 0.0002599 |
| PDZD3 | 79849 | PDZ domain containing 3 | -1.1019319 | 0.01934555 |
| BHLHE40 | 8553 | basic helix-loop-helix family mer | -1.1094305 | 0.01480105 |
| ACE2 | 59272 | angiotensin I converting enzyme | -1.12023 | 0.03380218 |
| MIR215 | 406997 | microRNA 215 | -1.1342186 | 0.04385956 |
| GKN2 | 200504 | gastrokine 2 | -1.1745722 | 0.00429197 |
| MIR548U | 100422884 | microRNA 548u | -1.1921643 | 0.0264929 |
| FAXDC2 | 10826 | fatty acid hydroxylase domain co | -1.2102276 | 0.00063807 |
| SLC28A1 | 9154 | solute carrier family 28 member | -1.2306428 | 0.0046388 |
| FADS2 | 9415 | fatty acid desaturase 2 | -1.231235 | 0.04360947 |
| P4HA1 | 5033 | prolyl 4-hydroxylase subunit alph | -1.2608865 | 0.00290433 |
| PLOD2 | 5352 | procollagen-lysine,2-oxoglutarate | -1.2758716 | 0.04432339 |
| RAB31 | 11031 | RAB31, member RAS oncogene f | -1.3107342 | 0.00475156 |
| TCN1 | 6947 | transcobalamin 1 | -1.3237344 | 0.01430078 |
| SLC26A9 | 115019 | solute carrier family 26 member | -1.3905118 | 0.02514264 |
| MIR8066 | 102465868 | microRNA 8066 | -1.4157102 | 0.03363763 |
| SEMA3E | 9723 | semaphorin 3E | -1.4161063 | 0.00269422 |

| | | | | |
|---|---|---|---|---|
| TDRD7 | 23424 | tudor domain containing 7 | -0.6451241 | 0.00787021 |
| SAMD8 | 142891 | sterile alpha motif domain containing 8 | -0.6489521 | 0.04559136 |
| FAM96B | 51647 | family with sequence similarity 96 member | -0.6500158 | 0.00200456 |
| MIRLET7E | 406887 | microRNA let-7e | -0.6503202 | 0.00771631 |
| CDHR5 | 53841 | cadherin related family member 5 | -0.6515906 | 0.01867046 |
| ZDBF2 | 57683 | zinc finger DBF-type containing 2 | -0.6521431 | 0.03864404 |
| TMEM117 | 84216 | transmembrane protein 117 | -0.6523436 | 0.03093138 |
| RAB6B | 51560 | RAB6B, member RAS oncogene family | -0.6532701 | 0.00879027 |
| COL17A1 | 1308 | collagen type XVII alpha 1 chain | -0.6549097 | 0.00528698 |
| TMEM65 | 157378 | transmembrane protein 65 | -0.6549892 | 0.02174638 |
| ISM1-AS1 | 100505536 | ISM1 antisense RNA 1 | -0.6579055 | 0.02818064 |
| EGR1 | 1958 | early growth response 1 | -0.6580278 | 0.01724591 |
| FAM3B | 54097 | family with sequence similarity 3 member I | -0.658516 | 0.02922881 |
| CPQ | 10404 | carboxypeptidase Q | -0.6595065 | 0.02819253 |
| ANG | 283 | angiogenin | -0.6600634 | 0.00565361 |
| CYP2C18 | 1562 | cytochrome P450 family 2 subfamily C mem | -0.6609367 | 0.04556628 |
| NAB1 | 4664 | NGFI-A binding protein 1 | -0.6614929 | 0.01391501 |
| HMOX1 | 3162 | heme oxygenase 1 | -0.6619124 | 0.02448664 |
| PMM1 | 5372 | phosphomannomutase 1 | -0.6625603 | 0.02223383 |
| SLC35D1 | 23169 | solute carrier family 35 member D1 | -0.6642758 | 0.00859772 |
| HPRT1 | 3251 | hypoxanthine phosphoribosyltransferase 1 | -0.6668391 | 0.03713595 |
| SLC7A8 | 23428 | solute carrier family 7 member 8 | -0.6694096 | 0.00553008 |
| STYK1 | 55359 | serine/threonine/tyrosine kinase 1 | -0.6699088 | 0.00947194 |
| KLK1 | 3816 | kallikrein 1 | -0.6700858 | 0.01319979 |
| SLC10A5 | 347051 | solute carrier family 10 member 5 | -0.6713328 | 0.01509482 |
| SOX21-AS1 | 100507533 | SOX21 antisense divergent transcript 1 | -0.6721567 | 0.01424128 |
| DISP1 | 84976 | dispatched RND transporter family member | -0.6737457 | 0.00879027 |
| ACVR1C | 130399 | activin A receptor type 1C | -0.6777047 | 0.00278167 |
| BPNT1 | 10380 | 3'(2'), 5'-bisphosphate nucleotidase 1 | -0.6779598 | 0.00370042 |
| XKR9 | 389668 | XK related 9 | -0.6822073 | 0.02900739 |
| CA2 | 760 | carbonic anhydrase 2 | -0.6867282 | 0.00549755 |
| CYP4F2 | 8529 | cytochrome P450 family 4 subfamily F mem | -0.6895518 | 0.02402397 |
| ARHGAP42 | 143872 | Rho GTPase activating protein 42 | -0.6900975 | 0.0163018 |
| TMEM120A | 83862 | transmembrane protein 120A | -0.6905078 | 0.00638187 |
| BPIFB1 | 92747 | BPI fold containing family B member 1 | -0.691058 | 0.01856524 |
| PCDHB10 | 56126 | protocadherin beta 10 | -0.6921165 | 0.01598383 |
| SLC13A2 | 9058 | solute carrier family 13 member 2 | -0.6973153 | 0.02190605 |
| FAM47E | 100129583 | family with sequence similarity 47 member | -0.6973804 | 0.01764373 |
| RFC2 | 5982 | replication factor C subunit 2 | -0.698904 | 0.0139131 |
| CASR | 846 | calcium sensing receptor | -0.7014162 | 0.03392666 |
| ZNF488 | 118738 | zinc finger protein 488 | -0.703095 | 0.0042616 |
| FGFBP1 | 9982 | fibroblast growth factor binding protein 1 | -0.7034695 | 0.01370992 |
| P2RY1 | 5028 | purinergic receptor P2Y1 | -0.7035747 | 0.04222458 |
| HIST1H1C | 3006 | histone cluster 1 H1 family member c | -0.7048761 | 0.00222082 |
| IPMK | 253430 | inositol polyphosphate multikinase | -0.7054062 | 0.00628842 |
| B4GALT1-AS1 | 101929639 | B4GALT1 antisense RNA 1 | -0.7055598 | 0.02283909 |
| STRIP2 | 57464 | striatin interacting protein 2 | -0.707595 | 0.02474602 |
| MORN5 | 254956 | MORN repeat containing 5 | -0.7076546 | 0.03735059 |
| ENPP1 | 5167 | ectonucleotide pyrophosphatase/phosphodi | -0.7100244 | 0.02818064 |
| OSTM1 | 28962 | osteopetrosis associated transmembrane p | -0.7100988 | 0.024336 |
| TSPAN13 | 27075 | tetraspanin 13 | -0.7134782 | 0.00386342 |
| LINC00881 | 100498859 | long intergenic non-protein coding RNA 881 | -0.7142659 | 0.03700889 |
| CDCA3 | 83461 | cell division cycle associated 3 | -0.7169029 | 0.01065413 |
| CENPJ | 55835 | centromere protein J | -0.7210018 | 0.02264169 |
| RHOF | 54509 | ras homolog family member F, filopodia as | -0.722918 | 0.00515273 |
| ACSF2 | 80221 | acyl-CoA synthetase family member 2 | -0.7230414 | 0.03772217 |
| SRGAP2C | 653464 | SLIT-ROBO Rho GTPase activating protein 2 | -0.7247232 | 0.00238386 |
| HACL1 | 26061 | 2-hydroxyacyl-CoA lyase 1 | -0.7263716 | 0.02672931 |
| ARRDC3 | 57561 | arrestin domain containing 3 | -0.7296554 | 0.03596635 |
| EXOC6B | 23233 | exocyst complex component 6B | -0.7319491 | 0.00845467 |
| TAS2R50 | 259296 | taste 2 receptor member 50 | -0.7340089 | 0.01936399 |
| TRANK1 | 9881 | tetratricopeptide repeat and ankyrin repeat | -0.7341088 | 0.00969196 |
| SLC16A3 | 9123 | solute carrier family 16 member 3 | -0.7349109 | 0.02068385 |
| FADS6 | 283985 | fatty acid desaturase 6 | -0.7361721 | 0.01094839 |
| LINC01127 | 100506328 | long intergenic non-protein coding RNA 112 | -0.7366179 | 0.01370992 |
| HACD3 | 51495 | 3-hydroxyacyl-CoA dehydratase 3 | -0.7383097 | 0.00731198 |
| AHCYL2 | 23382 | adenosylhomocysteinase like 2 | -0.7384391 | 0.00299034 |
| VIPR1 | 7433 | vasoactive intestinal peptide receptor 1 | -0.7389579 | 0.02091509 |
| DEPDC1B | 55789 | DEP domain containing 1B | -0.7390328 | 0.017129 |
| RHPN2 | 85415 | rhophilin Rho GTPase binding protein 2 | -0.7390954 | 0.00161934 |
| CPM | 1368 | carboxypeptidase M | -0.739398 | 0.00428765 |
| PLAGL1 | 5325 | PLAG1 like zinc finger 1 | -0.7398609 | 0.02816682 |
| CCNG2 | 901 | cyclin G2 | -0.7406131 | 0.00776591 |
| DHRS11 | 79154 | dehydrogenase/reductase 11 | -0.742994 | 0.0140763 |

| | | | | |
|---|---|---|---|---|
| BPIFB1 | 92747 | BPI fold containing family B men | -1.4171227 | 0.00031413 |
| MUC5AC | 4586 | mucin 5AC, oligomeric mucus/ge | -1.4382486 | 0.00620708 |
| REP15 | 387849 | RAB15 effector protein | -1.4519645 | 0.01594405 |
| AQP10 | 89872 | aquaporin 10 | -1.486324 | 0.01194033 |
| CD36 | 948 | CD36 molecule | -1.5012939 | 0.00553376 |
| PCK1 | 5105 | phosphoenolpyruvate carboxykina | -1.5127964 | 0.00386458 |
| GAS2L3 | 283431 | growth arrest specific 2 like 3 | -1.5145683 | 0.03024505 |
| LINC01559 | 283422 | long intergenic non-protein codir | -1.5311984 | 8.27E-05 |
| DHRS9 | 10170 | dehydrogenase/reductase 9 | -1.5905765 | 0.00655304 |
| MIR548AU | 100847045 | microRNA 548au | -1.5979714 | 0.0430881 |
| ANKFN1 | 162282 | ankyrin repeat and fibronectin ty | -1.6019069 | 0.02009829 |
| SLC11A2 | 4891 | solute carrier family 11 member | -1.6173094 | 0.00272165 |
| MIR5687 | 100847019 | microRNA 5687 | -1.627064 | 0.02266863 |
| RNASE1 | 6035 | ribonuclease A family member 1 | -1.768953 | 6.37E-05 |
| CXCR4 | 7852 | C-X-C motif chemokine receptor | -1.8182634 | 0.03227903 |
| MSMB | 4477 | microseminoprotein beta | -1.8196348 | 0.00184513 |
| UCA1 | 652995 | urothelial cancer associated 1 (n | -1.9458365 | 0.00080315 |
| PSCA | 8000 | prostate stem cell antigen | -2.0662814 | 2.70E-05 |
| TFF2 | 7032 | trefoil factor 2 | -2.0689684 | 2.17E-05 |
| EGLN3 | 112399 | egl-9 family hypoxia inducible fa | -2.1425182 | 0.00755328 |
| ALDOC | 230 | aldolase, fructose-bisphosphate | -2.2449962 | 0.00012826 |
| UGT2B11 | 10720 | UDP glucuronosyltransferase fam | -2.3117279 | 6.58E-05 |
| ST8SIA6 | 338596 | ST8 alpha-N-acetyl-neuraminide | -2.3256702 | 0.01152192 |
| CAPN9 | 10753 | calpain 9 | -2.351904 | 7.55E-05 |
| GPX3 | 2878 | glutathione peroxidase 3 | -2.511567 | 1.79E-05 |
| FAM177B | 400823 | family with sequence similarity 1 | -2.519225 | 2.73E-05 |
| LINC00443 | 100874173 | long intergenic non-protein codir | -3.0792113 | 9.91E-05 |

| | | | | |
|---|---|---|---|---|
| REG4 | 83998 | regenerating family member 4 | -0.7460323 | 0.006096 |
| SULT1A1 | 6817 | sulfotransferase family 1A member 1 | -0.747668 | 0.00148994 |
| TPK1 | 27010 | thiamin pyrophosphokinase 1 | -0.7479265 | 0.02287513 |
| FABP7 | 2173 | fatty acid binding protein 7 | -0.7484043 | 0.03087028 |
| PER3 | 8863 | period circadian regulator 3 | -0.7484953 | 0.0009314 |
| IL22RA1 | 58985 | interleukin 22 receptor subunit alpha 1 | -0.7498003 | 0.00879027 |
| TMEM141 | 85014 | transmembrane protein 141 | -0.7500946 | 0.01221887 |
| TMEM14A | 28978 | transmembrane protein 14A | -0.7516944 | 0.00131114 |
| HIST1H4L | 8368 | histone cluster 1 H4 family member l | -0.7516984 | 0.01989426 |
| LINC01220 | 731223 | long intergenic non-protein coding RNA 122 | -0.752822 | 0.01640753 |
| ACOT11 | 26027 | acyl-CoA thioesterase 11 | -0.753878 | 0.00282706 |
| CBR1 | 873 | carbonyl reductase 1 | -0.7561766 | 0.02483811 |
| LINC01336 | 104326191 | long intergenic non-protein coding RNA 133 | -0.7571321 | 0.02811033 |
| TSPAN5 | 10098 | tetraspanin 5 | -0.7571382 | 0.00147482 |
| RARB | 5915 | retinoic acid receptor beta | -0.7581345 | 0.00263885 |
| RDH5 | 5959 | retinol dehydrogenase 5 | -0.758238 | 0.00328578 |
| HSD17B11 | 51170 | hydroxysteroid 17-beta dehydrogenase 11 | -0.759447 | 0.00243073 |
| MYO1A | 4640 | myosin IA | -0.7596239 | 0.00798763 |
| RNFT1 | 51136 | ring finger protein, transmembrane 1 | -0.7620638 | 0.00409179 |
| AKR1B15 | 441282 | aldo-keto reductase family 1 member B15 | -0.7626563 | 0.00511987 |
| CTNNAL1 | 8727 | catenin alpha like 1 | -0.7643036 | 0.04733568 |
| SLC28A1 | 9154 | solute carrier family 28 member 1 | -0.7710875 | 0.0329782 |
| AQP12A | 375318 | aquaporin 12A | -0.7764631 | 0.0163018 |
| SLC22A18AS | 5003 | solute carrier family 22 member 18 antisen | -0.7775214 | 0.00282706 |
| MIER3 | 166968 | MIER family member 3 | -0.7800665 | 0.03065001 |
| HIST1H3H | 8357 | histone cluster 1 H3 family member h | -0.7826372 | 0.00648512 |
| DDX60L | 91351 | DExD/H-box 60 like | -0.7847162 | 0.01424128 |
| NCKAP5 | 344148 | NCK associated protein 5 | -0.7862091 | 0.01834964 |
| SAMD9 | 54809 | sterile alpha motif domain containing 9 | -0.7885337 | 0.00127435 |
| FER1L6 | 654463 | fer-1 like family member 6 | -0.7889792 | 0.00644906 |
| EBP | 10682 | emopamil binding protein (sterol isomerase | -0.7894468 | 0.00234724 |
| DSN1 | 79980 | DSN1 homolog, MIS12 kinetochore complex | -0.7926685 | 0.04278584 |
| APOBEC3B | 9582 | apolipoprotein B mRNA editing enzyme cat | -0.7952417 | 0.03056874 |
| HIST1H4D | 8360 | histone cluster 1 H4 family member d | -0.7976704 | 0.01094839 |
| SLC27A2 | 11001 | solute carrier family 27 member 2 | -0.7978247 | 0.00953026 |
| P4HA1 | 5033 | prolyl 4-hydroxylase subunit alpha 1 | -0.7984479 | 0.02264169 |
| STK17B | 9262 | serine/threonine kinase 17b | -0.802565 | 0.01416295 |
| IDI1 | 3422 | isopentenyl-diphosphate delta isomerase 1 | -0.8039539 | 0.00327644 |
| PEPD | 5184 | peptidase D | -0.8039925 | 0.00278167 |
| HK2 | 3099 | hexokinase 2 | -0.8044718 | 0.04496043 |
| COL6A1 | 1291 | collagen type VI alpha 1 chain | -0.8048629 | 0.00321611 |
| USP2 | 9099 | ubiquitin specific peptidase 2 | -0.8049361 | 0.0355943 |
| FAM13A | 10144 | family with sequence similarity 13 member | -0.8053688 | 0.0493943 |
| ANGPTL4 | 51129 | angiopoietin like 4 | -0.8057056 | 0.01178738 |
| GSTO1 | 9446 | glutathione S-transferase omega 1 | -0.8061134 | 0.00063291 |
| CYP3A5 | 1577 | cytochrome P450 family 3 subfamily A mem | -0.8067042 | 0.0049046 |
| RNA5S9 | 100169760 | RNA, 5S ribosomal 9 | -0.8079332 | 0.04260995 |
| PSD3 | 23362 | pleckstrin and Sec7 domain containing 3 | -0.8169738 | 0.00055821 |
| STRADB | 55437 | STE20-related kinase adaptor beta | -0.8175957 | 0.00562609 |
| P3H2 | 55214 | prolyl 3-hydroxylase 2 | -0.8190142 | 0.02470951 |
| RNA5S1 | 100169751 | RNA, 5S ribosomal 1 | -0.8210084 | 0.04143086 |
| RNA5S10 | 100169761 | RNA, 5S ribosomal 10 | -0.8210084 | 0.04143086 |
| RNA5S11 | 100169762 | RNA, 5S ribosomal 11 | -0.8210084 | 0.04143086 |
| RNA5S2 | 100169753 | RNA, 5S ribosomal 2 | -0.8210084 | 0.04143086 |
| RNA5S3 | 100169754 | RNA, 5S ribosomal 3 | -0.8210084 | 0.04143086 |
| RNA5S4 | 100169755 | RNA, 5S ribosomal 4 | -0.8210084 | 0.04143086 |
| RNA5S6 | 100169757 | RNA, 5S ribosomal 6 | -0.8210084 | 0.04143086 |
| MT1X | 4501 | metallothionein 1X | -0.8227502 | 0.00278167 |
| MIR642A | 693227 | microRNA 642a | -0.8234696 | 0.00407527 |
| FABP12 | 646486 | fatty acid binding protein 12 | -0.8251107 | 0.00683915 |
| FUT11 | 170384 | fucosyltransferase 11 | -0.8252834 | 0.02590335 |
| TMEM139 | 135932 | transmembrane protein 139 | -0.8275346 | 0.0014344 |
| PLAC8 | 51316 | placenta specific 8 | -0.8316391 | 0.00108344 |
| PADI2 | 11240 | peptidyl arginine deiminase 2 | -0.8338049 | 0.0262345 |
| GRAMD1C | 54762 | GRAM domain containing 1C | -0.8363632 | 0.02566474 |
| NEK2 | 4751 | NIMA related kinase 2 | -0.8401618 | 0.02303169 |
| VSIG2 | 23584 | V-set and immunoglobulin domain containi | -0.8402944 | 0.00112841 |
| SLC16A5 | 9121 | solute carrier family 16 member 5 | -0.8415316 | 0.00266571 |
| FAM69A | 388650 | family with sequence similarity 69 member | -0.842988 | 0.02768862 |
| SECTM1 | 6398 | secreted and transmembrane 1 | -0.8433404 | 0.01330393 |
| AKR1B10 | 57016 | aldo-keto reductase family 1 member B10 | -0.8442504 | 0.00406387 |
| BTNL8 | 79908 | butyrophilin like 8 | -0.8473038 | 0.01066023 |
| RNU4ATAC | 100151683 | RNA, U4atac small nuclear (U12-dependent | -0.8480131 | 0.00096118 |
| DPYD-AS1 | 100873932 | DPYD antisense RNA 1 | -0.852464 | 0.03848784 |

| | | | | |
|---|---|---|---|---|
| SLC23A3 | 151295 | solute carrier family 23 member 3 | -0.8528341 | 0.00511987 |
| PEX5L-AS2 | 101928790 | PEX5L antisense RNA 2 | -0.8545092 | 0.03593442 |
| SULT1A4 | 445329 | sulfotransferase family 1A member 4 | -0.8550653 | 0.00035662 |
| A1CF | 29974 | APOBEC1 complementation factor | -0.8574754 | 0.01163007 |
| OAT | 4942 | ornithine aminotransferase | -0.8581186 | 0.00169506 |
| ABCG1 | 9619 | ATP binding cassette subfamily G member | -0.8646008 | 0.02658288 |
| SLC9A4 | 389015 | solute carrier family 9 member A4 | -0.8647051 | 0.04752529 |
| SLC26A4 | 5172 | solute carrier family 26 member 4 | -0.8670076 | 0.04424537 |
| ESPL1 | 9700 | extra spindle pole bodies like 1, separase | -0.8680981 | 0.03324571 |
| ANO3 | 63982 | anoctamin 3 | -0.8704523 | 0.01856524 |
| MBOAT2 | 129642 | membrane bound O-acyltransferase domai | -0.8704667 | 0.01138615 |
| ACSS2 | 55902 | acyl-CoA synthetase short chain family men | -0.87366 | 0.00099227 |
| VTN | 7448 | vitronectin | -0.8750941 | 0.00584288 |
| ADH1A | 124 | alcohol dehydrogenase 1A (class I), alpha po | -0.8755534 | 0.01131409 |
| BCO1 | 53630 | beta-carotene oxygenase 1 | -0.8767489 | 0.00667557 |
| ACAT2 | 39 | acetyl-CoA acetyltransferase 2 | -0.8779406 | 0.004053 |
| FLVCR1 | 28982 | feline leukemia virus subgroup C cellular rec | -0.8795894 | 0.00263885 |
| NDRG1 | 10397 | N-myc downstream regulated 1 | -0.8823633 | 0.02287513 |
| ERP27 | 121506 | endoplasmic reticulum protein 27 | -0.8839685 | 0.00347904 |
| MIA2 | 4253 | melanoma inhibitory activity 2 | -0.8843688 | 0.01364154 |
| MEP1A | 4224 | meprin A subunit alpha | -0.8872281 | 0.02252994 |
| RBP4 | 5950 | retinol binding protein 4 | -0.8883298 | 0.04214547 |
| KNG1 | 3827 | kininogen 1 | -0.8895543 | 0.01650885 |
| LIN7A | 8825 | lin-7 homolog A, crumbs cell polarity compl | -0.8904977 | 0.02341232 |
| TEX15 | 56154 | testis expressed 15, meiosis and synapsis a | -0.898878 | 0.04238229 |
| UGT1A1 | 54658 | UDP glucuronosyltransferase family 1 mem | -0.9070346 | 0.01286059 |
| MXD1 | 4084 | MAX dimerization protein 1 | -0.9076513 | 0.00134588 |
| SMPDL3A | 10924 | sphingomyelin phosphodiesterase acid like | -0.9136995 | 0.00194195 |
| MMP28 | 79148 | matrix metallopeptidase 28 | -0.9144466 | 0.00536248 |
| GAL3ST1 | 9514 | galactose-3-O-sulfotransferase 1 | -0.9178459 | 0.00276248 |
| UGT2B10 | 7365 | UDP glucuronosyltransferase family 2 mem | -0.9201517 | 0.02422437 |
| OASL | 8638 | 2'-5'-oligoadenylate synthetase like | -0.9223273 | 0.00081817 |
| FMO5 | 2330 | flavin containing monooxygenase 5 | -0.9235607 | 0.00141512 |
| UGCG | 7357 | UDP-glucose ceramide glucosyltransferase | -0.9259768 | 0.00206586 |
| PAQR7 | 164091 | progestin and adipoQ receptor family mem | -0.9265551 | 0.01172913 |
| GPAT3 | 84803 | glycerol-3-phosphate acyltransferase 3 | -0.9316497 | 0.00511987 |
| INSIG1 | 3638 | insulin induced gene 1 | -0.9339832 | 0.00025438 |
| FA2H | 79152 | fatty acid 2-hydroxylase | -0.935695 | 0.00027929 |
| AQP3 | 360 | aquaporin 3 (Gill blood group) | -0.9360864 | 0.0134066 |
| BTNL3 | 10917 | butyrophilin like 3 | -0.9382694 | 0.00702089 |
| UGT2B28 | 54490 | UDP glucuronosyltransferase family 2 mem | -0.9383227 | 0.03269393 |
| DUSP5 | 1847 | dual specificity phosphatase 5 | -0.9400783 | 0.01013816 |
| NAT2 | 10 | N-acetyltransferase 2 | -0.9410522 | 0.0018525 |
| IL7 | 3574 | interleukin 7 | -0.9429795 | 0.01544184 |
| KCNS3 | 3790 | potassium voltage-gated channel modifier s | -0.944616 | 0.00033511 |
| PAG1 | 55824 | phosphoprotein membrane anchor with glyc | -0.9474112 | 0.01416566 |
| APOL1 | 8542 | apolipoprotein L1 | -0.947494 | 0.00882845 |
| IFNE | 338376 | interferon epsilon | -0.9507048 | 0.00081817 |
| EFNA1 | 1942 | ephrin A1 | -0.9567663 | 0.024336 |
| GKN2 | 200504 | gastrokine 2 | -0.9623655 | 0.00798763 |
| SLC2A1 | 6513 | solute carrier family 2 member 1 | -0.9641374 | 0.01530409 |
| DNASE1 | 1773 | deoxyribonuclease 1 | -0.9681435 | 0.01343959 |
| IL6R | 3570 | interleukin 6 receptor | -0.9714101 | 0.002804 |
| EMP1 | 2012 | epithelial membrane protein 1 | -0.9745016 | 0.00287567 |
| GREB1L | 80000 | GREB1 like retinoic acid receptor coactivato | -0.9750596 | 0.00188893 |
| IFI27 | 3429 | interferon alpha inducible protein 27 | -0.9762412 | 0.00022484 |
| DUSP1 | 1843 | dual specificity phosphatase 1 | -0.9788056 | 0.00033511 |
| BMP8A | 353500 | bone morphogenetic protein 8a | -0.9788715 | 0.0009314 |
| NT5E | 4907 | 5'-nucleotidase ecto | -0.9832963 | 0.00261623 |
| CSRP2 | 1466 | cysteine and glycine rich protein 2 | -0.9864828 | 0.00512916 |
| SYTL5 | 94122 | synaptotagmin like 5 | -0.9870918 | 0.01189036 |
| MALL | 7851 | mal, T cell differentiation protein like | -0.9875747 | 0.00192692 |
| CEACAM20 | 125931 | carcinoembryonic antigen related cell adhes | -0.9885693 | 0.02350878 |
| ADIRF | 10974 | adipogenesis regulatory factor | -0.9905468 | 0.00151599 |
| TEX30 | 93081 | testis expressed 30 | -0.9912676 | 0.00165149 |
| GPCPD1 | 56261 | glycerophosphocholine phosphodiesterase 1 | -0.9947784 | 0.00769698 |
| ADI1 | 55256 | acireductone dioxygenase 1 | -0.994919 | 0.00079302 |
| AQP11 | 282679 | aquaporin 11 | -0.9991814 | 0.00176029 |
| CLDN23 | 137075 | claudin 23 | -0.9992382 | 0.00112027 |
| TRIM40 | 135644 | tripartite motif containing 40 | -1.0012043 | 0.00248918 |
| PRSS55 | 203074 | serine protease 55 | -1.0021658 | 0.02296462 |
| THRB | 7068 | thyroid hormone receptor beta | -1.0073977 | 0.00165453 |
| OPN3 | 23596 | opsin 3 | -1.0093554 | 0.00565361 |
| MIR4753 | 100616224 | microRNA 4753 | -1.0130682 | 0.00580209 |

| | | | | |
|---|---|---|---|---|
| VAMP8 | 8673 | vesicle associated membrane protein 8 | -1.0189035 | 0.00026637 |
| MOGAT3 | 346606 | monoacylglycerol O-acyltransferase 3 | -1.0227633 | 0.0009314 |
| MS4A8 | 83661 | membrane spanning 4-domains A8 | -1.0231496 | 0.00750314 |
| CTSV | 1515 | cathepsin V | -1.0261953 | 0.00032768 |
| RBPMS2 | 348093 | RNA binding protein, mRNA processing fact | -1.0325766 | 0.0070941 |
| CLDN18 | 51208 | claudin 18 | -1.0337077 | 0.009321 |
| NEK10 | 152110 | NIMA related kinase 10 | -1.0367565 | 0.00222082 |
| TTR | 7276 | transthyretin | -1.0380556 | 0.00200456 |
| IL17RB | 55540 | interleukin 17 receptor B | -1.0506343 | 0.00029253 |
| ARL14 | 80117 | ADP ribosylation factor like GTPase 14 | -1.056874 | 0.00080724 |
| FAXDC2 | 10826 | fatty acid hydroxylase domain containing 2 | -1.0586275 | 0.00093536 |
| TMEM37 | 140738 | transmembrane protein 37 | -1.0592315 | 0.01138615 |
| SERPINI1 | 5274 | serpin family I member 1 | -1.0613839 | 0.02287513 |
| OTC | 5009 | ornithine carbamoyltransferase | -1.0639388 | 0.00165149 |
| MXRA5 | 25878 | matrix remodeling associated 5 | -1.0655238 | 0.00111346 |
| ITLN1 | 55600 | intelectin 1 | -1.0725136 | 0.00453723 |
| AADAC | 13 | arylacetamide deacetylase | -1.0875703 | 0.0070524 |
| HPGD | 3248 | 15-hydroxyprostaglandin dehydrogenase | -1.0939933 | 0.00278167 |
| TMOD1 | 7111 | tropomodulin 1 | -1.0963607 | 0.00803621 |
| APOL4 | 80832 | apolipoprotein L4 | -1.1003129 | 0.00248918 |
| VILL | 50853 | villin like | -1.1016737 | 0.0001383 |
| SLC31A2 | 1318 | solute carrier family 31 member 2 | -1.1054881 | 0.00014546 |
| PHGR1 | 644844 | proline, histidine and glycine rich 1 | -1.108437 | 0.0010823 |
| FRRS1 | 391059 | ferric chelate reductase 1 | -1.1095995 | 0.00053711 |
| ALDOB | 229 | aldolase, fructose-bisphosphate B | -1.1098354 | 0.00470812 |
| SLC26A3 | 1811 | solute carrier family 26 member 3 | -1.1112476 | 0.00028908 |
| FUOM | 282969 | fucose mutarotase | -1.1167611 | 0.00047726 |
| SNORA11 | 677799 | small nucleolar RNA, H/ACA box 11 | -1.1174494 | 0.00671844 |
| RNASE1 | 6035 | ribonuclease A family member 1, pancreati | -1.125867 | 0.00111445 |
| SEMA6C | 10500 | semaphorin 6C | -1.126777 | 0.004607 |
| SDR16C5 | 195814 | short chain dehydrogenase/reductase famil | -1.1307721 | 0.00258266 |
| ASAH2 | 56624 | N-acylsphingosine amidohydrolase 2 | -1.1325081 | 0.01776104 |
| CRIP1 | 1396 | cysteine rich protein 1 | -1.1341487 | 0.00036503 |
| SLC46A1 | 113235 | solute carrier family 46 member 1 | -1.1369326 | 0.00371794 |
| SLC3A1 | 6519 | solute carrier family 3 member 1 | -1.1371875 | 0.00112841 |
| CHST5 | 23563 | carbohydrate sulfotransferase 5 | -1.1376343 | 0.00151599 |
| BHLHE40 | 8553 | basic helix-loop-helix family member e40 | -1.1393616 | 0.00906551 |
| PKIB | 5570 | cAMP-dependent protein kinase inhibitor be | -1.1530106 | 0.00118153 |
| SULT1C3 | 442038 | sulfotransferase family 1C member 3 | -1.1555307 | 0.02609339 |
| PFKFB4 | 5210 | 6-phosphofructo-2-kinase/fructose-2,6-biph | -1.1662332 | 0.02420714 |
| GUCA2B | 2981 | guanylate cyclase activator 2B | -1.1670557 | 0.00326692 |
| SYTL4 | 94121 | synaptotagmin like 4 | -1.185164 | 0.00059228 |
| IL1B | 3553 | interleukin 1 beta | -1.1911654 | 0.02699596 |
| TFF1 | 7031 | trefoil factor 1 | -1.1998417 | 7.19E-05 |
| FABP1 | 2168 | fatty acid binding protein 1 | -1.2133754 | 0.00700744 |
| GPA33 | 10223 | glycoprotein A33 | -1.2168482 | 0.00713505 |
| CEACAM18 | 729767 | carcinoembryonic antigen related cell adhes | -1.2244097 | 0.00969196 |
| LRP4 | 4038 | LDL receptor related protein 4 | -1.2244296 | 0.00655449 |
| SLC36A1 | 206358 | solute carrier family 36 member 1 | -1.2249292 | 6.84E-05 |
| MIR194-1 | 406969 | microRNA 194-1 | -1.2274491 | 0.01159099 |
| TMEM253 | 643382 | transmembrane protein 253 | -1.2349202 | 0.0001261 |
| SULT1E1 | 6783 | sulfotransferase family 1E member 1 | -1.2382573 | 0.03412984 |
| TM6SF2 | 53345 | transmembrane 6 superfamily member 2 | -1.2383286 | 0.00068172 |
| TUBAL3 | 79861 | tubulin alpha like 3 | -1.2441656 | 0.00413244 |
| APOA1 | 335 | apolipoprotein A1 | -1.2469922 | 0.02768614 |
| PRAP1 | 118471 | proline rich acidic protein 1 | -1.2551883 | 6.84E-05 |
| CA1 | 759 | carbonic anhydrase 1 | -1.2599139 | 0.01867046 |
| RAB3B | 5865 | RAB3B, member RAS oncogene family | -1.2641236 | 0.0001383 |
| CLIC5 | 53405 | chloride intracellular channel 5 | -1.264631 | 0.01530409 |
| ADM | 133 | adrenomedullin | -1.2760577 | 0.00916209 |
| CAPN9 | 10753 | calpain 9 | -1.2941042 | 0.00346267 |
| TMCC3 | 57458 | transmembrane and coiled-coil domain fam | -1.2959534 | 0.00230315 |
| ID1 | 3397 | inhibitor of DNA binding 1, HLH protein | -1.3024589 | 6.84E-05 |
| LIFR | 3977 | LIF receptor alpha | -1.3100796 | 0.00309341 |
| RNF186 | 54546 | ring finger protein 186 | -1.3223056 | 0.00494949 |
| ANPEP | 290 | alanyl aminopeptidase, membrane | -1.337648 | 6.84E-05 |
| AQP10 | 89872 | aquaporin 10 | -1.3394818 | 0.0140763 |
| SLC4A7 | 9497 | solute carrier family 4 member 7 | -1.3490884 | 0.00248926 |
| SLC11A2 | 4891 | solute carrier family 11 member 2 | -1.3512252 | 0.0045439 |
| AQP7 | 364 | aquaporin 7 | -1.3581606 | 0.00015167 |
| IL1R2 | 7850 | interleukin 1 receptor type 2 | -1.3642705 | 0.00110253 |
| SFRP5 | 6425 | secreted frizzled related protein 5 | -1.374308 | 0.00097696 |
| NAAA | 27163 | N-acylethanolamine acid amidase | -1.3832842 | 0.00215429 |
| CYP3A7 | 1551 | cytochrome P450 family 3 subfamily A mem | -1.398136 | 0.00060101 |

| | | | | |
|---|---|---|---|---|
| PHLPP2 | 23035 | PH domain and leucine rich repeat protein p | -1.4078813 | 0.00081817 |
| SLC5A4 | 6527 | solute carrier family 5 member 4 | -1.4192629 | 0.01749623 |
| MMP1 | 4312 | matrix metallopeptidase 1 | -1.4339071 | 0.00061469 |
| PDZD3 | 79849 | PDZ domain containing 3 | -1.4401242 | 0.00294818 |
| SCIN | 85477 | scinderin | -1.4649594 | 0.00248926 |
| FLVCR2 | 55640 | feline leukemia virus subgroup C cellular rec | -1.4753531 | 0.00248926 |
| SI | 6476 | sucrase-isomaltase | -1.4881972 | 0.01138615 |
| TFF2 | 7032 | trefoil factor 2 | -1.4887125 | 0.00013484 |
| ACER2 | 340485 | alkaline ceramidase 2 | -1.4927516 | 9.88E-05 |
| SMIM24 | 284422 | small integral membrane protein 24 | -1.4995706 | 6.84E-05 |
| APOBEC1 | 339 | apolipoprotein B mRNA editing enzyme cata | -1.5096989 | 0.00015689 |
| PON3 | 5446 | paraoxonase 3 | -1.5236547 | 0.00127435 |
| TM4SF5 | 9032 | transmembrane 4 L six family member 5 | -1.5269889 | 3.82E-05 |
| FRMD3 | 257019 | FERM domain containing 3 | -1.5332872 | 0.00112841 |
| PNMA1 | 9240 | PNMA family member 1 | -1.5336077 | 2.94E-05 |
| MOGAT2 | 80168 | monoacylglycerol O-acyltransferase 2 | -1.534212 | 0.00219155 |
| EPB41L3 | 23136 | erythrocyte membrane protein band 4.1 like | -1.5354756 | 0.01725508 |
| SLC6A8 | 6535 | solute carrier family 6 member 8 | -1.5596516 | 0.00715236 |
| MIR215 | 406997 | microRNA 215 | -1.5856105 | 0.00674013 |
| SLC7A9 | 11136 | solute carrier family 7 member 9 | -1.5861536 | 0.01506465 |
| PLA2G12B | 84647 | phospholipase A2 group XIIB | -1.6039618 | 0.00127703 |
| KLK7 | 5650 | kallikrein related peptidase 7 | -1.6424803 | 1.26E-05 |
| GDPD2 | 54857 | glycerophosphodiester phosphodiesterase d | -1.6453046 | 0.00146775 |
| GAS2L3 | 283431 | growth arrest specific 2 like 3 | -1.6461806 | 0.01598383 |
| SUSD2 | 56241 | sushi domain containing 2 | -1.6575229 | 0.00648512 |
| CYP27A1 | 1593 | cytochrome P450 family 27 subfamily A me | -1.6846705 | 0.01103778 |
| HRASLS2 | 54979 | HRAS like suppressor 2 | -1.6891799 | 1.26E-05 |
| PTPRR | 5801 | protein tyrosine phosphatase, receptor type | -1.6896346 | 0.00120315 |
| ID3 | 3399 | inhibitor of DNA binding 3, HLH protein | -1.7208887 | 4.25E-05 |
| XPNPEP2 | 7512 | X-prolyl aminopeptidase 2 | -1.7227813 | 0.00363774 |
| RBP2 | 5948 | retinol binding protein 2 | -1.7323876 | 0.02229176 |
| MMP12 | 4321 | matrix metallopeptidase 12 | -1.760809 | 0.004053 |
| MSMB | 4477 | microseminoprotein beta | -1.7865021 | 0.00111346 |
| KCNJ13 | 3769 | potassium voltage-gated channel subfamily | -1.8207626 | 0.00263885 |
| PLOD2 | 5352 | procollagen-lysine,2-oxoglutarate 5-dioxyge | -1.8235087 | 0.0061054 |
| HHLA2 | 11148 | HERV-H LTR-associating 2 | -1.836576 | 2.94E-05 |
| SEMA5A | 9037 | semaphorin 5A | -1.8422784 | 3.82E-05 |
| CIDEB | 27141 | cell death-inducing DFFA-like effector b | -1.8488771 | 3.14E-05 |
| CDHR2 | 54825 | cadherin related family member 2 | -1.8507356 | 0.00101363 |
| SMLR1 | 100507203 | small leucine rich protein 1 | -1.8589573 | 0.00206586 |
| REP15 | 387849 | RAB15 effector protein | -1.8668178 | 0.00255404 |
| SLC2A5 | 6518 | solute carrier family 2 member 5 | -1.8741218 | 0.00831486 |
| EGLN3 | 112399 | egl-9 family hypoxia inducible factor 3 | -1.8778047 | 0.00986325 |
| MGAM | 8972 | maltase-glucoamylase | -1.8794622 | 0.00247061 |
| CDA | 978 | cytidine deaminase | -1.8990185 | 0.00014422 |
| HNF4A-AS1 | 101927219 | HNF4A antisense RNA 1 | -1.901531 | 0.00937922 |
| UCA1 | 652995 | urothelial cancer associated 1 (non-protein | -1.9052292 | 0.00052419 |
| IL2RG | 3561 | interleukin 2 receptor subunit gamma | -1.9382627 | 1.26E-05 |
| DKK1 | 22943 | dickkopf WNT signaling pathway inhibitor 1 | -1.9397167 | 0.0007324 |
| OTOP3 | 347741 | otopetrin 3 | -1.9705092 | 7.19E-05 |
| GCNT4 | 51301 | glucosaminyl (N-acetyl) transferase 4, core | -2.0031371 | 6.89E-05 |
| LINC01559 | 283422 | long intergenic non-protein coding RNA 155 | -2.0254448 | 1.26E-05 |
| MAL | 4118 | mal, T cell differentiation protein | -2.0644384 | 0.00054241 |
| ALDOC | 230 | aldolase, fructose-bisphosphate C | -2.0959803 | 0.0001383 |
| TMEM236 | 653567 | transmembrane protein 236 | -2.1014113 | 0.0176175 |
| SEMA3E | 9723 | semaphorin 3E | -2.130714 | 7.47E-05 |
| SLC2A2 | 6514 | solute carrier family 2 member 2 | -2.1361501 | 0.00740245 |
| RAB31 | 11031 | RAB31, member RAS oncogene family | -2.1866118 | 7.19E-05 |
| LEAP2 | 116842 | liver enriched antimicrobial peptide 2 | -2.2174814 | 0.00013046 |
| CIDEC | 63924 | cell death inducing DFFA like effector c | -2.2443713 | 6.49E-05 |
| AGMO | 392636 | alkylglycerol monooxygenase | -2.2490558 | 0.00130361 |
| SULT1B1 | 27284 | sulfotransferase family 1B member 1 | -2.2598291 | 0.0001261 |
| ABCG2 | 9429 | ATP binding cassette subfamily G member | -2.2765791 | 0.00013796 |
| CREB3L3 | 84699 | cAMP responsive element binding protein 3 | -2.289806 | 0.00020534 |
| ADH4 | 127 | alcohol dehydrogenase 4 (class II), pi polype | -2.3041243 | 0.00022645 |
| DHRS9 | 10170 | dehydrogenase/reductase 9 | -2.3385905 | 0.00029253 |
| ST8SIA6 | 338596 | ST8 alpha-N-acetyl-neuraminide alpha-2,8- | -2.3762813 | 0.00648512 |
| PSCA | 8000 | prostate stem cell antigen | -2.3997936 | 1.26E-05 |
| SLC15A1 | 6564 | solute carrier family 15 member 1 | -2.4207621 | 0.00022484 |
| LINC00443 | 100874173 | long intergenic non-protein coding RNA 443 | -2.4689989 | 0.00036503 |
| MS4A10 | 341116 | membrane spanning 4-domains A10 | -2.5478198 | 0.00107341 |
| CYP3A4 | 1576 | cytochrome P450 family 3 subfamily A mem | -2.6070429 | 0.0001261 |
| MEP1B | 4225 | meprin A subunit beta | -2.6383332 | 0.00027691 |
| ACE2 | 59272 | angiotensin I converting enzyme 2 | -2.731119 | 9.88E-05 |

| | | | | |
|---|---|---|---|---|
| MTTP | 4547 | microsomal triglyceride transfer protein | -2.7670114 | 6.49E-05 |
| UGT2B11 | 10720 | UDP glucuronosyltransferase family 2 mem | -2.8216312 | 1.45E-05 |
| PCK1 | 5105 | phosphoenolpyruvate carboxykinase 1 | -2.9382003 | 2.46E-05 |
| APOC3 | 345 | apolipoprotein C3 | -2.9407613 | 0.0001667 |
| G6PC | 2538 | glucose-6-phosphatase catalytic subunit | -2.9641097 | 7.47E-05 |
| FAM177B | 400823 | family with sequence similarity 177 membe | -2.977892 | 1.26E-05 |
| ABCC2 | 1244 | ATP binding cassette subfamily C member 2 | -3.0440141 | 6.89E-05 |
| CD36 | 948 | CD36 molecule | -3.3249848 | 1.45E-05 |
| APOB | 338 | apolipoprotein B | -3.3548687 | 6.84E-05 |
| GPX3 | 2878 | glutathione peroxidase 3 | -3.5421529 | 8.47E-07 |
